# Associations between forensic loci and neighboring gene expression levels may compromise medical privacy

**DOI:** 10.1101/2021.07.20.21260897

**Authors:** Mayra M. Bañuelos, Jhony A. Zavaleta, Alennie Roldan, Rochelle-Jan Reyes, Miguel Guardado, Berenice Chavez Rojas, Thet Nyein, Ana Rodriguez Vega, Maribel Santos, Emilia Huerta Sanchez, Rori Rohlfs

## Abstract

A set of 20 short tandem repeats (STRs) is used by the United States criminal justice system to identify suspects, and to maintain a database of genetic profiles for individuals who have been previously convicted or arrested. Some of these STRs were identified in the 1990s, with a preference for markers in putative gene deserts to avoid forensic profiles revealing protected medical information. We revisit that assumption, investigating whether forensic genetic profiles reveal information about gene expression variation, or potential medical information. We find six significant correlations (FDR = 0.23) between the forensic STRs and the expression levels of neighboring genes in lymphoblastoid cell lines. We explore possible mechanisms for these associations, with evidence compatible with forensic STRs causing expression variation, or being in LD with a causal locus in three cases, and weaker or potentially spurious associations in the other three cases. Together, these results suggest that forensic genetic loci may reveal expression level and, perhaps, medical information.

## INTRODUCTION

Forensic genetic identification in the United States is typically performed using genotype data from 20 short tandem repeats (STRs), known as the Combined DNA Index System (CODIS) core loci. Because these markers are highly polymorphic, even just 20 loci provide an immense amount of identifying information regarding a specific individual (Evett and Weir 1998). Thirteen of these CODIS core loci were established by the Federal Bureau of Investigation (FBI) in 1998. These loci were selected for efficient PCR multiplexing, while maximizing identifying information, and minimizing ancestry-based population differences and medically relevant information (J. M. Butler 2006). In 2017, seven additional STRs were added to the CODIS core loci, selected for similar criteria, particularly no known associations with medical conditions (Hares 2012).

It is important from a legal standpoint that CODIS genotypes do not reveal medical information. Laws authorizing the compulsory collection of DNA from certain persons may come into conflict with state privacy statutes or the U.S. Constitution if medical information is embedded (Murphy 2015). In fact, hundreds of court cases rely on the premise that the CODIS variants are uninformative, often citing this quote relating to the DNA Analysis Backlog Elimination Act of 2000, which states that the CODIS loci “were purposefully selected because they are not associated with any known physical or medical characteristics” (Letter from Robert Raben, Assistant Attorney General, to Judiciary Committee Chairman Henry Hyde). Yet, some of the CODIS loci, particularly those selected before the human genome was sequenced, are very close to genes. In fact, 11 of the CODIS loci are intronic (Katsanis and Wagner 2013). Any trait information conveyed by CODIS genotypes would raise questions regarding the medical privacy of individuals whose CODIS profiles are compelled by the government (including non-suspects, arrestees, and convicted individuals), as well as their genetic relatives. In particular, the historical and current treatment of arrested and convicted individuals is rife with rights unjustly curtailed, raising even more concern about a potentially lax approach to medical privacy for this population (Roth and Ainsworth 2015; Bauer 2016; Chesney-Lind and Mauer 2003). In this study, we re-examine the assumption that CODIS genotypes have no functional or medical impact.

It has long been known that variation in STR repeat number can alter gene function and regulation, sometimes resulting in dramatic phenotypes. A classic example is the coding STR expansion in the *HD* gene, which causes increasingly severe Huntington’s Disease (Mirkin 2007). Non-coding STRs have also been found to impact gene expression, resulting in trait variation. For instance, large numbers of repeats in an STR in the 5’ UTR of *FRAXA* impacts local methylation and gene regulation, causing Fragile X syndrome (Mirkin 2007).

More recent studies genome-wide surveys have found thousands of replicable associations between STR length and gene expression level (Gymrek et al. 2016; Quilez et al. 2016; Fotsing et al. 2019). STR length variation can impact methylation as well as histone modifications, causing evolutionarily conserved changes in gene expression (Gymrek et al. 2016; Quilez et al. 2016). Some of these STR-associated expression changes were associated with clinical traits (Gymrek et al. 2016). Somatic STR mutations have been implicated in the development of cancer (Fujimoto et al. 2020). One recent analysis showed that individuals with Autism have significantly more de novo STR mutations (particularly in introns) as compared with their neurotypical siblings (Mitra et al. 2021). This growing body of evidence suggests that STR length variation is causally responsible for a range of complex trait variation, including pathogenic conditions (Hannan 2018).

These results raise questions about whether the CODIS loci could impact medically relevant traits. Based on data available in 2011, a review of phenotypic associations with genetic loci concluded that there were no significant associations with the CODIS STRs (Katsanis and Wagner 2013). However, the study did report that some CODIS loci fall within predicted sites for genomic regulation, and all CODIS loci are within 1kb of at least one genetic variant associated with a phenotype (Katsanis and Wagner 2013). Because the LD surrounding the CODIS loci is strong enough to infer the genotypes of surrounding SNPs (Edge et al. 2017; Kim et al. 2018), phenotype information may be inferable through the CODIS genotypes. A more recent review of literature has identified 84 significant published associations between traits and STRs for 18 of the 20 CODIS loci (Wyner, Barash, and McNevin 2020).

Here we investigate whether genotypes at the CODIS loci could directly reveal information about a fundamental trait: the expression levels of neighboring genes. We identify CODIS loci significantly correlated with the expression of nearby genes (CODISeSTRs). We shed light on the mechanisms underlying these associations. First, we consider the possibility that the associations are caused by population structure as a confounding factor by testing for expression-genotype associations within subpopulations. With population stratification ruled out, we explore the possibility of CODISeSTRs causing expression variation by both comparing their genomic features to a panel of STRs with strong evidence of expression impact (Fotsing et al. 2019), and using a fine-mapping framework (CAVIAR) to identify putative causal loci (Hormozdiari et al. 2014). Finally, we investigate the hypothesis that CODISeSTRs may be in LD with a causal variant by examining their LD with putative causal sites identified by CAVIAR, as well as DHS sites.

## RESULTS

### Gene expression and the CODIS loci in the 1000 Genomes data set

We turned to a subset of the 1000 Genomes Project to investigate the relationship between gene expression levels and CODIS loci genotypes. STR length variation was not directly genotyped in the 1000 Genomes Project because this dataset used short-read sequencing. Thus, additional measures were taken to genotype these loci. Saini et al. (2018) imputed STR genotypes for the 1000 Genomes data by leveraging linkage disequilibrium between STRs and surrounding SNPs to create a publicly available haplotype reference panel. This haplotype reference panel includes 18 of the 20 core CODIS STRs currently in use (FBI n.d.). Genotypes for STRs D16S539 and D21S11 were unavailable because their unusually long alleles are challenging to impute from short read data.

While these imputed STR genotypes provide a tremendous resource, their accuracy is limited. The CODIS STRs in particular have a lower genotype imputation accuracy because of their extensive length and variation. Further, the accuracy of imputed STR genotypes is lower for individuals with non-European ancestry because the imputation training data consisted only of individuals with European ancestry, (Saini et al. 2018). The concordance between imputation and direct genotyping at a subset of the CODIS loci varied from 48% to 94%, even when benchmarking against a different European ancestry cohort (Saini et al. 2018). These limitations of the data erode power to detect signal related to CODIS loci genotypes, thus; we use a summary statistic, β, to describe the expected sum of alleles at a locus, given the probability assigned to each possible allele (see Methods).

We considered gene expression values based on transcriptome data from lymphoblastoid cell lines from 421 individuals in the 1000 Genomes Project. The populations represented in this set are: CEPH (CEU), Finns (FIN), British (GBR), Toscani (TSI) and Yoruba (YRI), each population with a sample size ranging from 89 to 95 individuals (Lappalainen, Sammeth, Friedländer, ‘t Hoen, et al. 2013). We investigated a model of CODIS STRs causing or being in LD with cis eQTLs by considering expression level variation of genes within 100kb of the CODIS loci. Out of the 18 CODIS STRs included in the haplotype reference panel, only 14 CODIS STRs are within 100kb of at least one gene that is expressed in the lymphoblastoid data (Supplemental Table 1). We considered a total of 39 CODIS STR-gene pairs, as the number of expressed genes within 100kb varied for each CODIS loci. For each CODIS STR-gene pair, we tested for correlation between CODIS loci genotypes and the expression levels of neighboring genes. Note that in this analysis we did not correct for population structure because we are not querying the molecular causality of an STR. Instead, we are investigating informative STR-expression associations, regardless of their cause.

**Table 1:** Genomic features of CODISeSTRs

| CODISeSTR | Location relative to genes | Distance to nearest TSS (genomic percentile) | Distance to TSS of associated gene | Distance to nearest DHS site (genomic percentile) | Distance to nearest lymph DHS site (genomic percentile) | Length (genomic percentile) | Repeating unit |
| --- | --- | --- | --- | --- | --- | --- | --- |
| D3S1358 | Intronic to LARS2 | 31,194 bp (52.6%) | 152,157 bp to LARS2 TSS | 1,916 bp (72.3%) | 4651 bp (69.1%) | 63 bp (96.7%) | [AGAT]<sub>n</sub> |
| D2S441 | Intergenic | 41,143 bp (45.8%) | 41,143 bp to C1D TSS. | 14,064 bp (28.1%) | 19,514 bp (39.3%) | 47 bp (93.6%) | [TGCC]<sub>m</sub>[TTCC]<sub>n</sub> |
| CSF1PO | Intronic to CSF1R | 4,649 bp (88.6%) | 4,649 bp to CSF1R TSS.<br>75,157 bp to TIGD6 TSS | 0 bp (100.0%) | 0 bp (100.0%) | 51 bp (94.8%) | [AGAT]<sub>n</sub> |
| D18S51 | Intronic to BCL2 | 36,928 bp (48.4%) | 85,535 bp to KDSR TSS | 13,230 bp (29.3%) | 3714 bp (73.2%) | 71 bp (97.3%) | [AGAA]<sub>n</sub> |
| FGA | Intronic to FGA | 2,922 bp (92.7%) | 2,922 bp to FGA TSS<br>37,303 bp to PLRG1 TSS | 8,065 bp (40%) | 27,933 (27.9%) | 87 bp (98.1%) | [TTTC]<sub>m</sub>TTTTTCT[C<br>TTT]<sub>n</sub>CTCC[TTCC]<sub>o</sub> |

Of the 39 CODIS STR-gene pairs tested, six showed significant correlations with *p-*values below 0.05 and a false discovery rate of 0.23 (so the expected number of false positives is 1.4 (Figure 1, Supplemental Table 1, Supplemental Figure 1). The strongest signal was between D3S1358 and *LARS2* (*p* = 1.1e-6, *r*^2^ = 0.059), while we see less strong correlations, although still significant, between CSF1PO and *CSF1R* (*p*=0.03, *r*^2^ = 0.01), between CSF1PO and *TIGD6* (*p*=0.04, *r*^2^ = 0.009), between D2S441 and *C1D* (*p*=0.01, *r*^2^ = 0.014), between D18S51 and *KDSR* (*p*=0.02, *r*^2^ = 0.011) and between FGA and *PLRG1* (*p=*0.03, *r*^2^ = 0.011) (Supplemental Table 1). While the coefficients of determination (*r*^*2*^) observed are weak, their statistical significance or marginal significance invites further investigation. The six CODIS STR-gene pairs with *p* < 0.05 represent an excess of correlation between CODIS loci and gene expression (*p*=2.9e-3, chi square test). We refer to the CODIS STRs associated with gene expression levels as CODISeSTRs.

**Figure 1:**
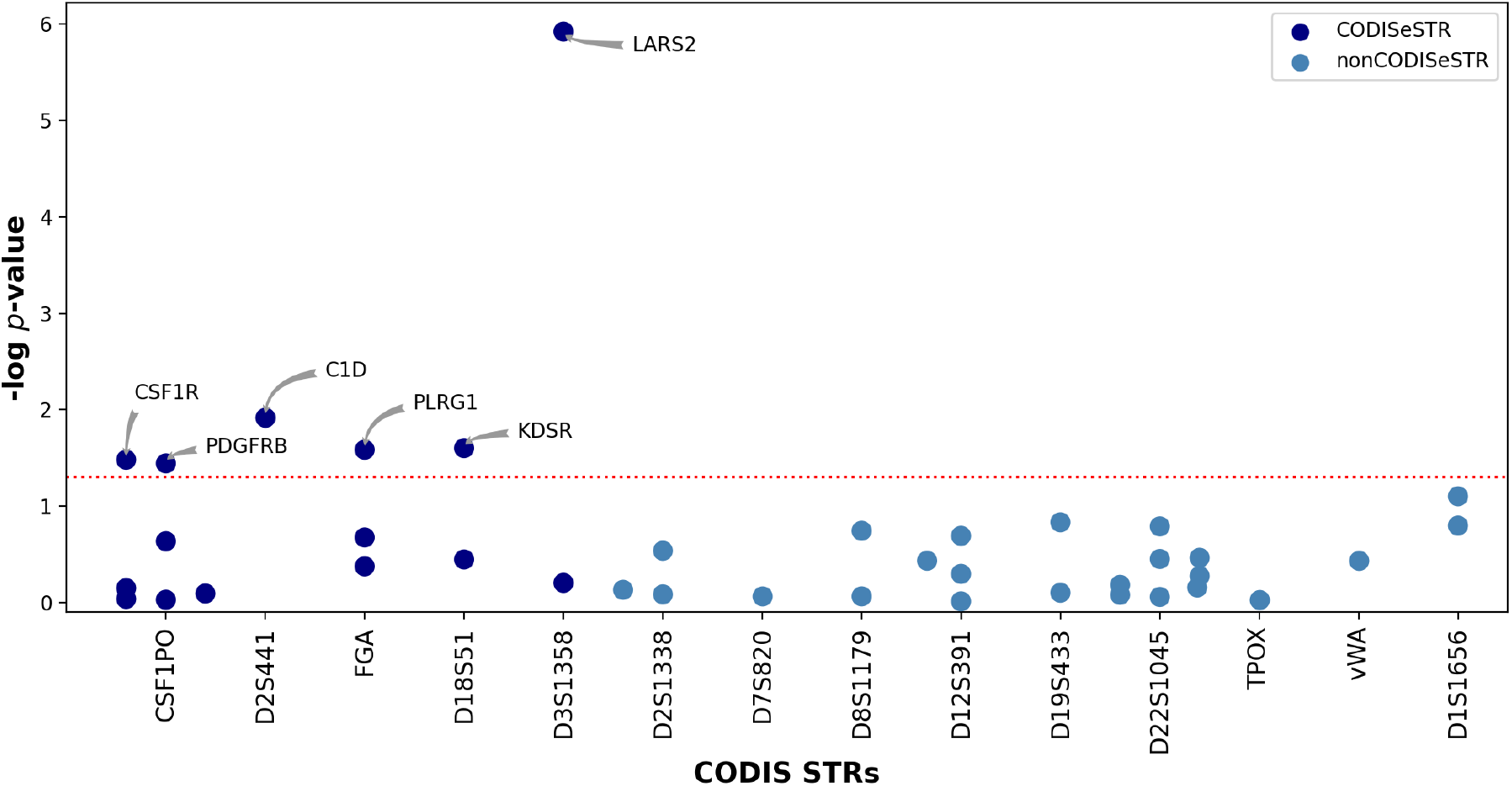
Correlations between CODIS loci and the expression of neighboring genes. Associations of CODIS STR-gene pairs are shown as negative log *p*-values. Red dotted line denotes the significant *p*-value threshold. CODISeSTRs are shown in dark blue and non-CODISeSTRs are shown in light blue.

The correlations that we observe could be explained by 1) CODISeSTRs causally impacting the expression of a neighboring gene, 2) LD between the CODISeSTR and a different causal locus that impacts expression, 3) a confounding factor like population structure and/or environmental variables in both CODISeSTR genotypes and expression levels, or 4) a spurious association in this particular dataset. If the correlation has any non-spurious basis, then the critical observation is that expression information may be inferred from the CODIS genotype, regardless of the precise mechanism for the association.

However, establishing a putative mechanism for each of the observed associations may inform us about its stability and generalizability. We explore these hypotheses in the following analyses.

### Exploring the role of population sub-structure in observed CODISeSTR correlations

First, we investigate if the observed CODISeSTR-expression level associations observed across the whole data set (including CEU, GBR, FIN, TSI, and YRI) are be caused by population structure as a confounding factor. We investigate this possibility by adding population membership as a covariate in the linear models of gene expression and CODISeSTR variation. For most CODISeSTR-gene pairs (D3S1358-*LARS2*, D18S51-*KDSR*, D2S441-*C1D*, and FGA-*PLRG1)* the associations remain significant with population as a covariate. Thus the associations observed are unlikely to be caused by population structure (Supplemental Table 2). For CSF1PO-*CSF1R* and CSF1PO-*TIGD6*, the associations are somewhat less pronounced with population as a covariate (CSF1PO-*CSF1R p-*value goes from 0.03 to 0.06 and CSF1PO-*TIGD6* 0.04 to 0.06). This suggests that the CSF1PO associations may be caused in part by population stratification.

**TABLE 2:**
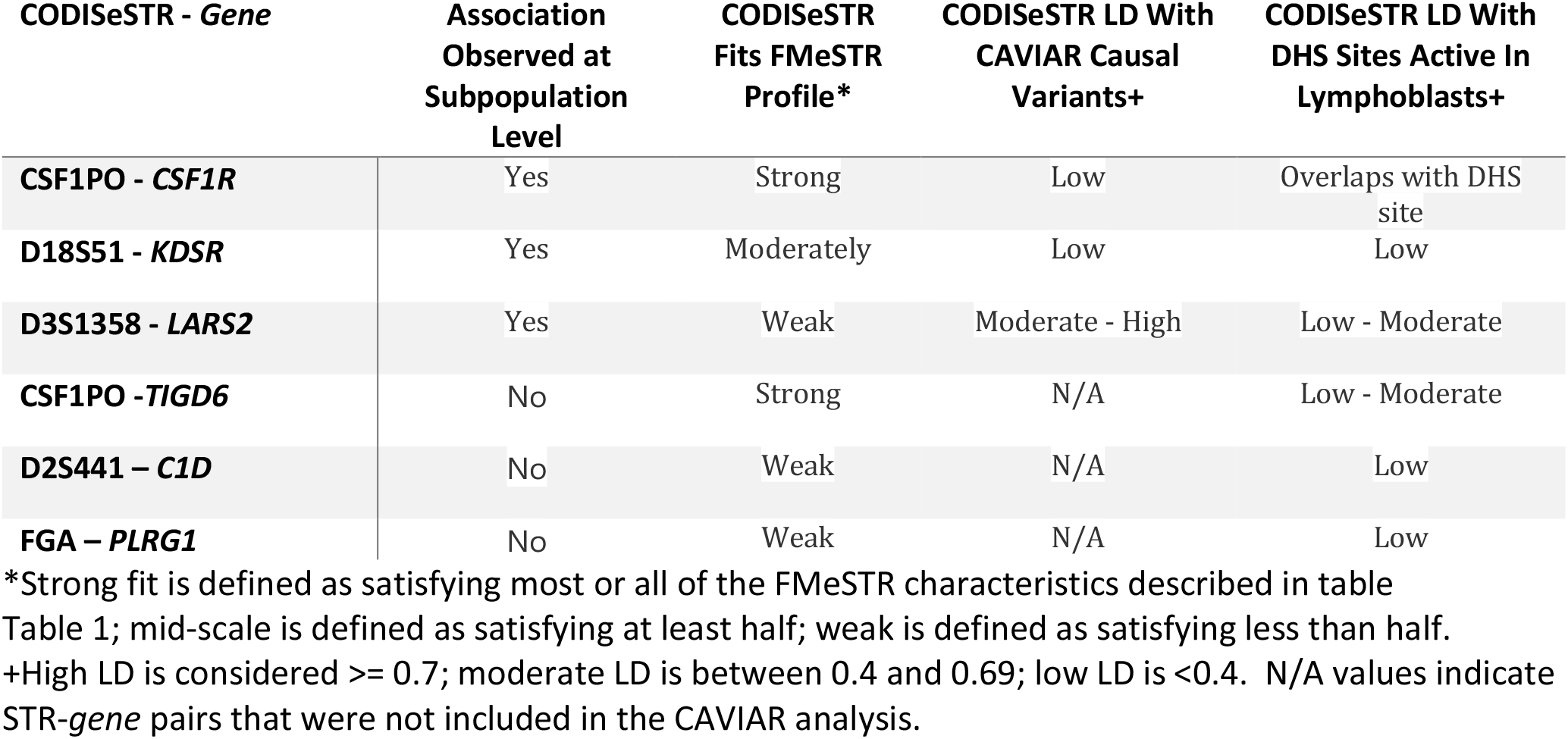
Putative mechanisms for observed CODISeSTR-expression associations

If a cumulative association is caused entirely by structure at the level of the specified subpopulations, then there would be no associations within subpopulations, and there would be differences in β and expression level distributions across subpopulations. We further investigate population-specific associations by testing for CODISeSTR-expression level associations within subpopulations (Supplemental Figure 2, Supplemental Table 3). We found 6 significant associations under a *p*-value threshold of 0.05, with false discovery rate of .21 (expected number of false positives is 1.3) (Supplemental Table 2, Supplemental Table 3).

**Figure 2:**
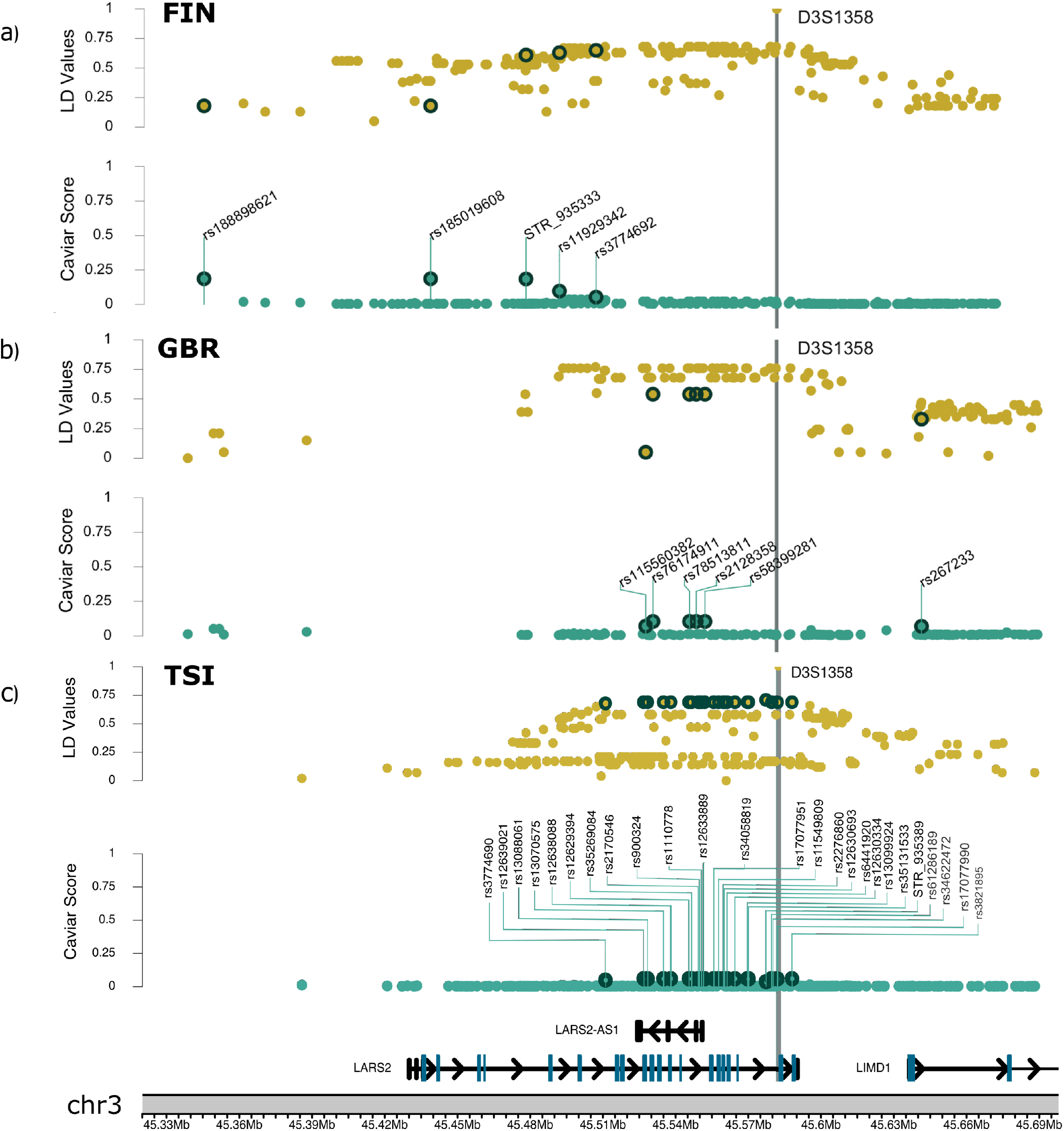
*LARS2*-D3S1358 CAVIAR and local LD landscape Local LD and CAVIAR score landscapes in a 100kb window centered on *LARS2* gene for a) FIN subpopulation, b) GBR subpopulation, and c) TSI subpopulation. For each plot, the top panel shows LD between the CODISeSTR D3S1358 versus each variant in the ρ causal set. Bottom panel shows CAVIAR scores for variants in the ρ causal set. Dark green circles enclose putative causal variants in both CAVIAR and LD panels.

For D3S1358-*LARS2*, D18S51-*KDSR*, and CSF1PO-*CSF1R*, we observe CODISeSTR-expression level associations within subpopulations. D3S1358 β is significantly correlated with *LARS2* expression in the FIN group (*p*=0.0013, *r*^2^ = 0.11) as well as showing a significant correlation in the TSI (*p*=0.005, *r*^2^ = 0.08) and the GBR (*p*=0.02, *r*^2^ = 0.06) group. This is consistent with the significant *p-*value in the cumulative population (*p*=1.12e-6, *r*^*2*^=0.06). Similarly, for D18S51 and *KDSR* we see a significant association in YRI (*p*=0.003, *r*^*2*^=0.11), with non-significant results for all other sub-populations. We again see consistent correlations in some subpopulations for the CSF1PO-*CSF1R* association in FIN (*p*=0.04, *r*^*2*^=0.04) and GBR (*p*=0.04, *r*^*2*^=0.04). These results suggest that for those CODISeSTR-gene pairs, the cumulative signal is not a product of population structure, but it may be driven by stronger associations in some subpopulation groups.

By contrast, while the associations for D2S441-*C1D*, CSF1PO-*TIGD6*, and FGA-*PLRG1* are significant in the cumulative dataset, within subpopulations we observe no significant or nominally significant correlations (Supplemental Table 3). To determine subpopulation structure is causing both the cumulative association and lack of associations within populations are due to, we tested for differences in the β and expression level distributions between subpopulations (Supplemental Table 4, Supplemental Figure 3). We do observe some significant differences in β distributions for FGA (YRI-CEU) and CSF1PO (YRI-FIN, -GBR, -TSI), but there are no corresponding significant differences in the expression distributions of *C1D, TIGD6*, or *PLRG1*. Thus, the significant associations for these CODISeSTR-gene pairs are not caused by this level of population structure as a confounding factor. Instead, it suggests either that the association is too weak to detect within subpopulations with decreased sample size and statistical power, or that the cumulative correlation is spurious.

### Comparing genomic features of CODISeSTRs and FMeSTRs

We go on to investigate if the CODISeSTRs resemble expression-associated STRs (eSTRs). A previous genomic analysis of a total of 1,620,030 STRs in humans identified 20,609 eSTRs with evidence of impacting gene expression (Fotsing et al. 2019). These eSTRs were then fine-mapped and ranked by their probability of causality using the statistical framework, CAVIAR (Hormozdiari et al. 2014). The eSTRs with the top 5% of probabilities of causality (1380 unique STRs) were then characterized as fine-mapped expression-associated STRs (FMeSTRs) to express the additional evidence for their impact on gene expression (Fotsing et al. 2019). Of note, three of the 20 CODIS STRs were previously identified as eSTRs by Fotsing *et al*. (2019), more than expected by chance (one-tailed binomial test, *p* = 0.002).

Specifically, expression associations were found for TPOX in tibial nerve tissue, for D2S1338 in heart left ventricle tissue and for THO1 in both visceral adipose and esophagus mucosa tissues (Fotsing et al. 2019). It is unsurprising that this study in lymphoblast cell lines did not reproduce those associations. However, these potential associations raise questions about expression associations of other CODIS loci across tissues.

The genomic features of FMeSTRs were characterized, showing that they are more likely to be long, intronic, located near transcription start sites (TSSs), located near DNAse I hypersensitivity (DHS) sites, and to contain particular repeating units. We examine how the CODISeSTRs fit the FMeSTR profile in order to investigate the hypothesis that the CODISeSTR genotypes are directly causing changes in the expression of neighboring genes (Table 1).

In general, the CODIS loci are similar to FMeSTRs in their extreme length — the CODISeSTRs, in particular, are all in at least the 93^rd^ percentile of lengths compared to all genomic STRs (Supplemental Figure 4). The CODISeSTRs further resemble FMeSTRs in that four of the five are intronic (CSF1PO:CSF1R; D18S51:BCL2; D3S1358:LARS2; FGA:FGA). Like FMeSTRs, two CODISeSTRs are unusually near to a TSS: FGA is 2.92 kilobases from the TSS of the gene FGA (92.7^d^ genomic percentile) and CSF1PO is 4.65 kilobases from the TSS of CSF1R (88.6^th^ genomic percentile) (Supplemental Figure 5). Similar to FMeSTRs which are disproportionately found near DHS sites, one CODISeSTR in particular overlaps with a DHS site observed in lymphoblasts or lymphoblast derivatives: CSF1PO (100^th^ genomic percentile) (Supplemental Figure 6, Supplemental Figure 7). Finally, the repeating units of four of the five CODISeSTRs have been found to be significantly enriched among eSTRs (D3S1358, CSF1PO, D2S441, D18S51) (Fotsing et al. 2019) (Table 1).

Altogether, CODISeSTRs, most particularly CSF1PO, fit the genomic profile of a FMeSTR that putatively impacts gene expression levels. These results are consistent with a hypothesis that CSF1PO has a causal impact on *CSF1R* expression levels, without being conclusive evidence.

### Identifying local genetic variants to explain observed variation in gene expression

Knowing that CODISeSTRs resemble STRs that impact expression, we attempted to determine whether the expression variation is being driven by each CODISeSTR itself, or if it is due to nearby causal genetic variants (other STRs or SNPs) in LD with the CODISeSTR. To identify causal cis variants, we use CAVIAR, a Bayesian fine-mapping framework that leverages pairwise LD and z-scores to identify a set of putatively causal variants (Hormozdiari et al. 2014). CAVIAR assigns a posterior probability (which we will refer to as “CAVIAR score”) to each marker in the ρ-causal set (Willems et al. 2017; Fotsing et al. 2019). With a ρ=0.95, the ρ-causal set is a subset of markers that with 95% confidence contains all causal variants.

Fine-mapping was performed in each subpopulation and CODISeSTR-gene combination for which we found a significant or nominally significant association (see section below “Exploring the role of population sub-structure in observed CODISeSTR correlations”). Thus, this causality analysis includes *LARS2* in the FIN, GBR, and TSI populations; *CSF1R* in the FIN and GBR populations, and *KDSR* in YRI.

These CAVIAR analyses produced scores between 0.04 and 0.60 for the putative causal variants (Supplemental Table 5). While the relatively small sample sizes (65-83 individuals) mean that power may be limited for some of these analyses, scores as high as 0.60 are noteworthy.

We also used CAVIAR to estimate the most likely *n* causal variants contributing to the phenotype with a max of *n*=4. The “putative causal set” is comprised of *n* variants in the ρ-causal set with the highest CAVIAR scores. While the CODISeSTRs were not tagged as putative causal variants, they do appear in most of the ρ-causal sets. For example, the highest CAVIAR score in a CODISeSTR is for D18S51 in the YRI subpopulation at 0.10.

### Investigating LD between CODISeSTRs and putative regulatory elements

The observed correlation between CODISeSTR genotypes and expression levels of neighboring genes could be caused by LD between a CODISeSTR and a regulatory variant that impacts gene expression. To investigate this possibility, we consider LD between CODISeSTRs and both CAVIAR-identified putative causal variants and DHS sites, indicating accessible DNA likely to contain regulatory elements.

In addition to the identification of CODISeSTR D18S51 in the ρ-causal set for *KDSR* expression in YRI, we observe high LD between CODISeSTR D3S1358 and the putative causal variants impacting *LARS2* expression levels (Supplemental Figure 7, Figure 2). Two of the putative causal variants for *LARS2* expression in FIN have LD of at least 0.61 with CODISeSTR D3S1358, while four of the putative causal variants for *LARS2* expression in GBR have LD of 0.54 with D3S1358, and the putative causal variants with top CAVIAR scores for LARS2 expression in TSI have LD of at least 0.68 with D3S1358. While the CAVIAR scores associated with these ρ-causal sets are modest, the convergent high LD values across subpopulations support the hypothesis that D3S1358 may be in LD with a causal locus contributing to LARS2 expression variation.

We also examined the LD between CODIS STRs and DHS sites within 100kb (Supplemental Figure 8). We measured STR-DHS site LD as the maximum correlation between the STR and a SNP located in the DHS site (Supplemental Figure 9, Supplemental Table 6). We observe high DHS site LD for some CODIS STRs (D1S1656, THO1, TPOX, vWA), and a large number of DHS sites surrounding others (CSF1PO, D22S1045, THO1). Note that of these, one is a CODISeSTR, while TPOX and THO1 were previously identified as eSTRs (Fotsing et al. 2019). We do not observe a general excess of LD with DHS sites for CODISeSTRs, as compared to other CODIS STRs (*p*=0.52, two-tailed Kolmogorov-Smirnov test).

Additionally, the CODISeSTR CSF1PO overlaps with a DHS, suggesting that variation in CSF1PO length may directly impact the action of that DHS. D3S1358 is in LD (*r*^*2*^>0.45) with SNPs in four DHSs detected in lymphoblasts, while D18S51 has LD of *r*^*2*^ = 0.31 with SNPs in one DHS (Supplemental Table 6).

### Putative mechanisms for observed CODISeSTR-expression associations

For each CODISeSTR-gene pair, we weighed the results supporting different mechanisms for the observed STR-gene expression association. (Table 2).

#### Association between D3S1358 and LARS2 expression

We observe concordant significant negative correlations between D3S1358 allele length and the *LARS2* expression levels in our cumulative 1000 genomes analysis (Supplemental Table 11). The strength of this correlation is demonstrated by the maintenance of the significant association within the smaller FIN subpopulation, as well as significant associations within GBR and TSI (Supplemental Table 2). As to the mechanism for this correlation, there is weak evidence suggesting that D3S1358 resembles a causal FMeSTR (Table 2). However, there is stronger evidence that D3S1358 is in LD with both a variant that putatively impacts LARS2 expression (Supplemental Table 5), and DHS sites active in lymphoblasts (Supplemental Table 6). Together, these results are consistent with the hypothesis that D3S1358 may be in LD with a locus which impacts *LARS2* expression.

#### Association between CSF1PO and CSF1R expression

*CSF1R* expression has a significant positive correlation with the genotype of intronic CODIS locus CSF1PO (Supplemental Table 1). The subpopulations FIN and GBR show consistent significant positive correlations (Supplemental Table 2), and there is only weak evidence that the association is partially due to population stratification (Supplemental Table 2). CSF1PO bears a remarkable resemblance to FMeSTRs with its close proximity to *CSF1R*’s TSS and particularly with its overlap with a DHS site found in lymph cell lines, as well as its length, and repeating unit (Table 1, Figure 9). These results are consistent with the hypothesis that CSF1PO could causally impact *CSF1R* expression, or be in LD with a different causal locus.

#### Association between D18S51 and KDSR expression

D18S51 β values are significantly correlated with *KDSR* expression across all samples, as well as within the YRI subpopulation (Supplemental Table 1, Supplemental Table 2). We note that the distribution of β values is significantly different in YRI compared to the other subpopulations (Supplemental Table 4, Supplemental Figure 3). Further, in the YRI subpopulation, the D18S51 was identified as the second most probable locus to cause *KDSR* expression variation with a CAVIAR score of 0.10 (Supplemental Table 5). Even if D18S51 itself is not causal, we note that its LD with a DHS site is 0.31 (Supplemental Table 6). Together, these results suggest that a correlation within the YRI subpopulation could be driving the correlation at the cumulative level, and that the correlation likely has a molecular basis (whether causal, or in LD with a causal locus).

#### Associations between CSF1PO and TIGD6 expression; D2S441 and C1D expression; and FGA and PLRG1 expression

For the remaining CODISeSTR-gene pairs, we see significant correlations at the cumulative population level, with no significant associations within subpopulations (Supplemental Table 1, Supplemental Table 2). While this might suggest the associations are due to population structure, two factors tell a different story: 1) the maintenance of a significant association with population as a covariate (Supplemental Table 2) and 2) the lack of consistent subpopulation differences in both β frequencies and expression levels tell a different story (Supplemental Table 4, Supplemental Figure 3). These results may be explained by either the smaller sub-population group sample sizes reducing power to detect weak correlations, or a spurious association in the cumulative analysis. This is consistent with the other analyses that show that D2S441 and FGA more weakly fit the pattern of FMeSTRs (Table 1) and relatively low LD to local DHS sites (Supplemental Table 6).

## DISCUSSION

CODIS loci were chosen because, at the time, researchers believed that no medical information would be revealed. However, in this study, we identified CODISeSTRs whose genotypes are correlated to the expression of neighboring genes in lymphoblast cell lines. Our results build on previous work finding expression associations with CODIS loci TPOX, THO1, and D2S1338 in other tissues (Fotsing et al. 2019). Specifically, we observed six significant correlations: between D3S1358 and *LARS2*, between D18S51 and *KDSR*, CSF1PO and both *CSF1R* and *TIGD6*, D2S441 and *C1D*, and FGA and *PLRG1*. We go on to investigate the putative mechanism for these correlations, finding that the associations between D3S1358-*LARS*, D18S51-*KDSR*, and CSF1PO-*CSF1R* are likely due to a causal relationship or LD with at least one causal locus, while the other associations are weaker or possibly spurious. These results provide evidence that contravenes the assumption that CODIS genotypes convey no trait information.

### Medical relevance of CODISeSTR-associated gene expression variance

Our analysis shows that a CODIS genotype profile can be used to infer the expression of some genes. This raises the question: would inferring those expression levels reveal medical information? We consulted the medical genetics literature to begin to address this question (Supplemental Table 7, Supplemental Text 1-6). We discuss some of the most striking cases: *CSF1R, LARS2*, and *KDSR*

#### CSF1R expression variation and neural and psychiatric conditions

CSF1R, which is intronic to CSF1PO, encodes a cytokine receptor that plays a key role in microglial regulation (Konno et al. 2014). Disruptive sequence mutations in CSF1R lead to a variety of brain conditions including leukoencephalopathy (Guo et al. 2019; Nicholson et al. 2013; Eichler et al. 2016; Rademakers et al. 2012; Oosterhof et al. 2019; Konno et al. 2014), while inhibition of CSf1R protein function seems to ameliorate some neural conditions like epilepsy (Srivastava et al. 2018), Alzheimer’s disease (Mancuso et al. 2019; Olmos-Alonso et al. 2016; Sosna et al. 2018), and spinal cord injury recovery (Bellver-Landete et al. 2019) (Supplemental Text 3). Further, and most relevantly for this study, variation in the expression and splicing CSF1R are associated with psychiatric conditions including depression and schizophrenia in humans (Zhang et al. 2020; Shimamoto-Mitsuyama et al. 2021; Gandal et al. 2018). Since CSF1R expression is correlated with CSF1PO, the CODIS genotype may be informative about those psychiatric conditions.

#### LARS2 and KDSR associations with medical conditions

LARS2 and KDSR function reduction or elimination have also been associated with medical conditions. LARS2, which contains D3S1538 in an intron, encodes a mitochondrial leucyl-tRNA synthetase gene (Bullard, Cai, and Spremulli 2000). LARS2 is well-established as an essential gene, as mutations that reduce or knock out its function have been associated with Perrault syndrome (Pierce et al. 2013; Willems et al. 2014; Soldà et al. 2016; Demain et al. 2017), MELAS syndrome (Li, Chomyn, and Guan 2010), and other conditions (Supplemental Text 1). KDSR, which is near to D18S51, encodes an enzyme involved in synthesis of the lipid ceramide. Mutations in KDSR that eliminate or decrease enzyme function have been associated with a number of severe skin and platelet conditions (Boyden et al. 2017; Bariana et al. 2019; Takeichi et al. 2017) (Supplemental Text 5). These medical genetic studies provide strong evidence that LARS2 and KDSR expression variation impact a number of medical conditions. The fact that dramatically reduced function leads to severe phenotypes raises the question whether marginally lowered expression may lead to intermediate conditions. The association between CODISeSTRs and those genes’ expression means that the CODIS genotype may be informative about risk of those conditions, or other intermediate phenotypes.

### Limitations

The associations reported here were observed in the subset of the 1000 Genomes Project where expression data was also available. These data are limited in a few important ways. First, expression data was only available in lymphoblastoid cell lines. This single cell type means that our analysis will miss genes which are not highly expressed, or whose expression isn’t regulated by cis-elements, in lymphoblastoid cell lines specifically. Second, data was only available from CEU, FIN, GBR, TSI, and YRI subpopulations. As four of five of these populations are European, they do not reflect the genetic diversity of the general population of the United States, notably with regards to their lack of admixture. Correlations due to population structure as a confounding factor may be underestimated in our analysis. Further, this analysis is unable to identify correlations that are specific to subpopulations not represented here. Third, errors in the imputation of the CODIS genotypes may erode power to identify associations, particularly in non-European subpopulations where imputation has higher error rates. In addition, our approach to detecting associations is specifically testing for a linear relationship between STR allele length and expression levels. While this type of linear relationship is generally expected (see Methods), there could be other non-linear relationships present that were not detected here.

Altogether, while our analysis produced significant correlations, it is limited in scope and underpowered. This raises the question of whether stronger correlations would be identified in an analysis on a larger more representative sample, with direct STR genotyping, using more expression data from more varied tissues.

### Conclusion

Within the limitations of the publicly available data examined here, our results suggest that information on gene expression levels may be revealed by CODIS profiles. Further, some of those gene expression levels have been connected to medical phenotypes. These results join a growing body of work showing that CODIS genotypes may contain more information than purely identity. CODIS profiles have been found to provide information about the surrounding haplotype (Edge et al. 2017; Kim et al. 2018), as well as genetic ancestry (Algee-Hewitt et al. 2016). Together, these findings raise concerns about the medical privacy of individuals whose CODIS profiles are seized, databased, and accessed, as well as the genetic relatives of those persons.

## MATERIALS AND METHODS

### 1000 Genomes Project CODIS Genotype Data

Phase 3 of the 1000 Genomes Project sampled 2,504 individuals from 26 different populations with ancestry from Africa, East Asia, Europe, South Asia and the Americas (Auton et al. 2015). The short-read sequencing approach used for this dataset presents a challenge for genotyping the CODIS loci, which are highly polymorphic, often with very long alleles. We used imputed CODIS loci genotype data that was made publicly available as a haplotype reference panel (see Online Resources for url) (Saini et al. 2018; Browning, Zhou, and Browning 2018). Because of the limits of this approach with particularly long alleles, genotypes for only 18 of the 20 CODIS STRs were successfully imputed (Saini et al. 2018). These 18 loci are D22S1045, TPOX, D2S441, D2S1338, vWA, D12S391, D5S818, CSF1PO, D1S1656, D10S1248, TH01, D13S317, D18S51, D19S433, D3S1358, FGA, D7S820, and D8S1179. The two CODIS STRs not included in our study are D16S539 and D21S11. We note that the very factors that make these loci difficult to impute (length and polymorphism) may make them particularly relevant for studies of phenotypic impact (Fotsing et al. 2019).

For our analysis of correlation between CODIS genotypes and expression levels, we created a summary statistic based on estimated allelic dosages generated by Beagle during imputation. For each individual, STR estimated allele dosages are the sum of the posterior allele probabilities for both haplotypes (Browning and Browning 2016). Hence, their values range from 0 to 2 (Yun et al. 2009).

We used the imputed STR allelic dosages to compute a normalized linear weighted genotype for each CODIS STR. We refer to this weighted average genotype as β (beta). We computed β for each individual for each CODIS STR using the following:

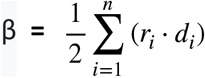

where *n* is the number of distinct alleles on record at the locus, *r*_*i*_ is the number of repeats in allele *i*, and *d*_*i*_ is its estimated allelic dosage (genotype probability). For non-CODIS STRs, β genotypes were computed by substituting *r*_*i*_ with the allele nucleotide length, instead of the repeat count.

### 1000 Genomes Project Gene Expression Data

Transcriptomes were typed from lymphoblastoid cell lines of 462 unrelated individuals from the 1000 Genomes Project (Lappalainen, Sammeth, Friedländer, ‘T Hoen, et al. 2013). The samples in this set correspond to five populations: the CEPH (CEU), Finns (FIN), British (GBR), Toscani (TSI) and Yoruba (YRI). Transcriptomic levels were quantified with Reads Per Kilobase of transcription per Million mapped reads (RPKM). Transcripts with 0 counts in more than half the number of samples were removed (Lappalainen, Sammeth, Friedländer, ‘T Hoen, et al. 2013). Full data is available at the EBI ArrayExpress portal, under accession E-GEUV-1 (see Online Resources for url).

Of the 462 individuals with gene expression data, 90 were filtered out because either β genotypes or gene expression values were missing. Our study was performed with 372 individuals for which we have CODIS genotype data and where at least one known gene within a 100kb window was expressed in the lymphoblastoid cell lines data. Within population analysis contained between 65 and 83 individuals.

### Testing associations between STR length and gene expression

Using data from the UCSC Genome Browser, we identified genes within 100 kilobases of the CODIS markers, measuring 100 kb from the start and end of each CODIS STR genomic location. We summarized the genotypes with β values, as detailed in section “1000 Genomes Project CODIS Genotype Data”. We next fit linear regression models to test for Pearson correlation between the β genotypes for each CODIS STR versus the expression levels of nearby genes.

The approach implicitly assumed a linear relationship between STR alleles by length. This assumption is justified by findings that STR impact on expression level scales with allele length (Tirosh, Barkai, and Verstrepen 2009; Mirkin 2007; Hannan 2018). For an STR that is not causal, but is in LD with a causal locus, the step-wise STR mutational process (Levinson and Gutman 1987; Ellegren 2004) will lead to multiple similarly-lengthed STR alleles on the causal haplotype.

We controlled for false discovery rate (FDR) using *q* values (Storey and Tibshirani 2003). With 6 features under a *p*-value threshold of 0.05, we expect 1.4 of those to be a false positive (Supplemental Table 2).

### Characterizing genomic features of CODISeSTRs

We quantified several genomic features of the CODISeSTRs in order to examine how they compare to the characteristics of putatively causative expression-altering STRs (referred to as FMeSTRs) (Fotsing et al. 2019). Genomic STR coordinates, including those of CODIS STRs and CODISeSTRs, were gathered from a genome-wide survey of STRs (Willems et al. 2017) (see Online Resources).

For context, we used the survey data to compute CODISeSTR lengths and their length percentiles as the proportion of genome-wide STRs that are at least as long. Distance, in bp, between STRs to the nearest gene and the nearest TSS was determined by additionally using genetic coordinates from the UCSC Genes track in the UCSC Genome Browser (Haeussler et al. 2019). We also used the UCSC Genes track to determine the distance of each STR relative to its associated gene(s) and the TSS(s) thereof. The repeating units for each CODISeSTR were gathered from STRbase (J. Butler n.d.) (see Online Resources).

Like in analysis of FMeSTRs (Willems et al. 2017; Fotsing et al. 2019), for each CODISeSTR, we computed the distance between each STR and the nearest DHS site. DHS site cluster locations were taken from the ENCODE Regulation ‘DNase Clusters’ track via the UCSC Genome Browser (Rosenbloom et al. 2013) (see Online Resources). All distances between STRs and nearby genomic elements, except for TSSs which are represented by the starting coordinate of the protein coding region, were calculated to reflect the distance between the closest endpoints of the elements in question.

For the general analysis, we considered DHS site clusters annotated in at least 20 sources. We performed additional analyses focusing on DHS site clusters observed in lymphoblasts or lymphoblast derivatives. We identified 20 cell lines that are lymphoblasts or lymphoblast derivatives, specifically Adult_CD4_Th0*, CD20+, CLL, CMK, GM06990, GM12864, GM12865, GM12878, GM12891, GM12892, GM18507, GM19238, GM19239, GM19240, HL-60, Jurkat, K562, NB4, Th1, and Th2. For lymphoblast specific analyses, we consider DHS sites that were observed in at least 5 of the 20 lymphoblasts or lymphoblast derivatives in the dataset.

### Evaluating the potential causality of cis variants

We performed a fine-mapping analysis with CAVIAR (Willems et al. 2017; Fotsing et al. 2019) to identify specific local genetic variants (either CODISeSTRs, other STRs or SNPs) that are putatively causal of the variation in expression levels. CAVIAR employs the variants’ LD structure as well as association statistics to predict a subset of variants, the ρ causal set, in which all causal markers are said to be included with a certain probability ρ, ρ = 95% in our case. Each variant in the CAVIAR ρ credible set is then assigned a probability of being causal. We refer to this posterior probability as CAVIAR score.

We considered SNPs and STRs within 100 kilobases up and downstream of genes with significant or marginally significant CODIS locus associations at the subpopulation level: *LARS2, CSF1R*, and *KDSR*. SNPs that did not exhibit variation within each subpopulation group were removed. We followed the CAVIAR protocol established by Fotsing *et al*. (Fotsing et al. 2019). Next, we filtered for SNPs and STRs that hold a significant association with gene expression level. Specifically, we tested for correlation between gene expression and either SNP or STR genotypes. For non-CODIS STRs, genotypes were considered as the nucleotide length of each allele and β values were computed, while for CODIS STRs we consider the number of repeats. Variants with a *p*-value > 0.05 were excluded from further analysis. Since it is unlikely for a phenotype to be caused by one variant alone, we allowed for CAVIAR to consider up to four independent causal variants per locus by including the parameters -f1 -c 4. We define the putative causal variants as the *n* number of variants with the highest CAVIAR scores, where *n* is CAVIAR’s predicted number of putative causal variants, ranging from 1 to 4. In the cases where a set of variants are in perfect LD with one another (and therefore have identical CAVIAR scores), the set is considered as a single prediction.

### Quantifying linkage disequilibrium

LD between STRs and SNPs was quantified as correlation between CODIS STR β values versus the SNP genotypes (sum of alternative alleles), implicitly testing a linear relationship. This measure of LD between STRs and SNPs is similar to a haplotype-based method shown to reliably follow the expected patterns of variation when applied to phased X-chromosome haplotypes (Willems et al. 2014). LD between two STRs was quantified as the correlation between β values. For CODISeSTRs, β was based on the number of repeats, while for non-CODIS STRs β was based on the nucleotide length. We used genotypic LD, rather than haplotypic LD, because the imputed STR estimated allele dosages lack phase information.

For analyses of LD between STRs and DHS sites, we calculated LD between CODIS loci and all SNPs within a DHS site. We considered DHS sites found within at least 5 of the 20 available lymphoblast cell lines or derivatives, as well as DHS sites found within at least 20 cell line sources (Supplemental Figure 8). As a summary, we considered the highest LD per DHS site (Supplemental Table 6). For DHS sites without SNPs in the region, LD was not computed and therefore not included in the summary.

### Online resources

CODIS STR genotypes imputed in 1000 Genomes Dataset: http://gymreklab.com/2018/03/05/snpstr_imputation.html

Transcriptome data from cell lines derived from individuals participating in 1000 Genomes Dataset: https://www.ebi.ac.uk/arrayexpress/experiments/E-GEUV-1/

Genome-wide STR survey:

https://github.com/HipSTR-Tool/HipSTR-references/blob/master/human/hg19.hipstr_reference.bed.gz

Technical details on CODIS STRs: https://strbase.nist.gov/str_fact.htm

DHS site locations from ENCODE: http://hgdownload.cse.ucsc.edu/goldenpath/hg19/encodeDCC/wgEncodeRegDnaseClustered/wgEncodeRegDnaseClusteredV3.bed.gz

## Data Availability

All data analyzed in the study is already published. The specific sources are referenced and links to specific tables are included in the "Online Resources" section of the manuscript.

http://gymreklab.com/2018/03/05/snpstr_imputation.html

https://www.ebi.ac.uk/arrayexpress/experiments/E-GEUV-1/

https://github.com/HipSTR-Tool/HipSTR-references/blob/master/human/hg19.hipstr_reference.bed.gz

https://strbase.nist.gov/str_fact.htm

http://hgdownload.cse.ucsc.edu/goldenpath/hg19/encodeDCC/wgEncodeRegDnaseClustered/wgEncodeRegDnaseClusteredV3.bed.gz

## Acknowledgements

We first thank all of the research participants who shared their genomic data through the publicly available sources used for this project. This research is only possible with their generosity. We are grateful to Doc Edge and Erin Murphy for thoughtful conversations and manuscript review that informed and improved this project. We thank Camille Rey for writing suggestions that improved this manuscript. San Francisco State University students Mayra Bañuelos, Jhony Zavaleta, Alennie Roldan, and Miguel Guardado were supported by the SFSU MBRS-RISE Fellowships (R25-GM059298), SFSU MARC Fellowships (T34-GM008574), Bridges Fellowships (R25-GM048972), and Genentech Foundation Fellowships. Funds through NIH-R35GM128946-01 supported Emilia Huerta Sanchez, funds through Brown University Predoctoral Training Program in Biological Data Science (NIH T32 GM128596) supported Mayra Bañuelos. The joint sixth authors were supported through the Big Data Summer Program funded by NIH 5R25MD011714-03.

## References

Algee-Hewitt, Bridget F.B., Michael D. Edge, Jaehee Kim, Jun Z. Li, and Noah A. Rosenberg. 2016. “Individual Identifiability Predicts Population Identifiability in Forensic Microsatellite Markers.” Current Biology 26 (7): 935–42. https://doi.org/10.1016/J.CUB.2016.01.065.

Auton, Adam, Gonçalo R. Abecasis, David M. Altshuler, Richard M. Durbin, David R. Bentley, Aravinda Chakravarti, Andrew G. Clark, et al. 2015. “A Global Reference for Human Genetic Variation.” Nature 526 (7571): 68–74. https://doi.org/10.1038/nature15393.

Bariana, Tadbir K., Veerle Labarque, Jessica Heremans, Chantal Thys, Mara De Reys, Daniel Greene, Benjamin Jenkins, et al. 2019. “Sphingolipid Dysregulation Due to Lack of Functional KDSR Impairs Proplatelet Formation Causing Thrombocytopenia.” Haematologica 104 (5): 1036–45. https://doi.org/10.3324/haematol.2018.204784.

Bauer, Shane. 2016. “My Four Months as a Private Prison Guard.” Mother Jones, 2016. https://www.motherjones.com/politics/2016/06/cca-private-prisons-corrections-corporation-inmates-investigation-bauer/.

Bellver-Landete, Victor, Floriane Bretheau, Benoit Mailhot, Nicolas Vallières, Martine Lessard, Marie Eve Janelle, Nathalie Vernoux, et al. 2019. “Microglia Are an Essential Component of the Neuroprotective Scar That Forms after Spinal Cord Injury.” Nature Communications 10 (1): 1–18. https://doi.org/10.1038/s41467-019-08446-0.

Boyden, Lynn M., Nicholas G. Vincent, Jing Zhou, Ronghua Hu, Brittany G. Craiglow, Susan J. Bayliss, Ilana S. Rosman, et al. 2017. “Mutations in KDSR Cause Recessive Progressive Symmetric Erythrokeratoderma.” American Journal of Human Genetics 100 (6): 978–84. https://doi.org/10.1016/j.ajhg.2017.05.003.

Browning, Brian L., and Sharon R. Browning. 2016. “Genotype Imputation with Millions of Reference Samples.” American Journal of Human Genetics 98 (1): 116–26. https://doi.org/10.1016/j.ajhg.2015.11.020.

Browning, Brian L., Ying Zhou, and Sharon R. Browning. 2018. “A One-Penny Imputed Genome from Next-Generation Reference Panels.” American Journal of Human Genetics 103 (3): 338–48. https://doi.org/10.1016/j.ajhg.2018.07.015.

Bullard, James M., Ying Chun Cai, and Linda L. Spremulli. 2000. “Expression and Characterization of the Human Mitochondrial Leucyl-TRNA Synthetase.” Biochimica et Biophysica Acta -Gene Structure and Expression 1490 (3): 245–58. https://doi.org/10.1016/S0167-4781(99)00240-7.

Butler, John. n.d. “STRBase: Overview of STR Fact Sheets.” Accessed August 18, 2020. https://strbase.nist.gov/str_fact.htm.

Butler, John M. 2006. “Genetics and Genomics of Core Short Tandem Repeat Loci Used in Human Identity Testing.” Journal of Forensic Sciences 51 (2): 253–65. https://doi.org/10.1111/j.1556-4029.2006.00046.x.

Chesney-Lind, Meda, and Marc Mauer, eds. 2003. Invisible Punishment: The Collateral Consequences of Mass Imprisonment. The New Press. https://www.amazon.com/Invisible-Punishment-Collateral-Consequences-Imprisonment/dp/1565848489.

Demain, L.A.M., J.E. Urquhart, J. O’Sullivan, S.G. Williams, S.S. Bhaskar, E.M. Jenkinson, C.M. Lourenco, et al. 2017. “Expanding the Genotypic Spectrum of Perrault Syndrome.” Clinical Genetics 91 (2): 302–12. https://doi.org/10.1111/cge.12776.

Edge, Michael D, Bridget F B Algee-Hewitt, Trevor J Pemberton, Jun Z Li, and Noah A Rosenberg. 2017. “Linkage Disequilibrium Matches Forensic Genetic Records to Disjoint Genomic Marker Sets.” Proceedings of the National Academy of Sciences of the United States of America 114 (22): 5671–76. https://doi.org/10.1073/pnas.1619944114.

Eichler, Florian S., Jiankang Li, Yiran Guo, Paul A. Caruso, Andrew C. Bjonnes, Jessica Pan, Jessica K. Booker, et al. 2016. “CSF1R Mosaicism in a Family with Hereditary Diffuse Leukoencephalopathy with Spheroids.” Brain 139 (6): 1666–72. https://doi.org/10.1093/brain/aww066.

Ellegren, Hans. 2004. “Microsatellites: Simple Sequences with Complex Evolution.” Nature Reviews Genetics. Nature Publishing Group. https://doi.org/10.1038/nrg1348.

Evett, Ian W., and Bruce S. Weir. 1998. Interpreting DNA Evidence: Statistical Genetics for Forensic Scientists. Sinauer Associates. https://www.amazon.com/Interpreting-DNA-Evidence-Statistical-Scientists/dp/B01JXQ5IVW.

FBI. n.d. “CODIS and NDIS Fact Sheet.” Accessed August 6, 2020. https://www.fbi.gov/services/laboratory/biometric-analysis/codis/codis-and-ndis-fact-sheet.

Fotsing, Stephanie Feupe, Jonathan Margoliash, Catherine Wang, Shubham Saini, Richard Yanicky, Sharona Shleizer-Burko, Alon Goren, and Melissa Gymrek. 2019. “The Impact of Short Tandem Repeat Variation on Gene Expression.” Nature Genetics 51 (11): 1652–59. https://doi.org/10.1038/s41588-019-0521-9.

Fujimoto, Akihiro, Masashi Fujita, Takanori Hasegawa, Jing Hao Wong, Kazuhiro Maejima, Aya Oku-Sasaki, Kaoru Nakano, et al. 2020. “Comprehensive Analysis of Indels in Whole-Genome Microsatellite Regions and Microsatellite Instability across 21 Cancer Types.” Genome Research 30 (3): 334–46. https://doi.org/10.1101/gr.255026.119.

Gandal, Michael J., Pan Zhang, Evi Hadjimichael, Rebecca L. Walker, Chao Chen, Shuang Liu, Hyejung Won, et al. 2018. “Transcriptome-Wide Isoform-Level Dysregulation in ASD, Schizophrenia, and Bipolar Disorder.” Science 362 (6420). https://doi.org/10.1126/science.aat8127.

Guo, Long, Débora Romeo Bertola, Asako Takanohashi, Asuka Saito, Yuko Segawa, Takanori Yokota, Satoru Ishibashi, et al. 2019. “Bi-Allelic CSF1R Mutations Cause Skeletal Dysplasia of Dysosteosclerosis-Pyle Disease Spectrum and Degenerative Encephalopathy with Brain Malformation.” American Journal of Human Genetics 104 (5): 925–35. https://doi.org/10.1016/j.ajhg.2019.03.004.

Gymrek, Melissa, Thomas Willems, Audrey Guilmatre, Haoyang Zeng, Barak Markus, Stoyan Georgiev, Mark J Daly, et al. 2016. “Abundant Contribution of Short Tandem Repeats to Gene Expression Variation in Humans.” Nature Genetics 48 (1): 22–29. https://doi.org/10.1038/ng.3461.

Haeussler, Maximilian, Ann S. Zweig, Cath Tyner, Matthew L. Speir, Kate R. Rosenbloom, Brian J. Raney, Christopher M. Lee, et al. 2019. “The UCSC Genome Browser Database: 2019 Update.” Nucleic Acids Research 47 (D1): D853–58. https://doi.org/10.1093/nar/gky1095.

Hannan, Anthony J. 2018. “Tandem Repeats Mediating Genetic Plasticity in Health and Disease.” Nature Reviews Genetics 19 (5): 286–98. https://doi.org/10.1038/nrg.2017.115.

Hares, Douglas R. 2012. “Expanding the CODIS Core Loci in the United States.” Forensic Science International. Genetics 6 (1): e52–4. https://doi.org/10.1016/j.fsigen.2011.04.012.

Hormozdiari, Farhad, Emrah Kostem, Eun Yong Kang, Bogdan Pasaniuc, and Eleazar Eskin. 2014. “Identifying Causal Variants at Loci with Multiple Signals of Association.” Genetics 198 (2): 497– 508. https://doi.org/10.1534/genetics.114.167908.

Katsanis, Sara H., and Jennifer K. Wagner. 2013. “Characterization of the Standard and Recommended CODIS Markers*.” Journal of Forensic Sciences 58 (January): S169–72. https://doi.org/10.1111/j.1556-4029.2012.02253.x.

Kim, Jaehee, Michael D. Edge, Bridget F.B. Algee-Hewitt, Jun Z. Li, and Noah A. Rosenberg. 2018. “Statistical Detection of Relatives Typed with Disjoint Forensic and Biomedical Loci.” Cell 175 (3): 848-858.e6. https://doi.org/10.1016/j.cell.2018.09.008.

Konno, Takuya, Masayoshi Tada, Mari Tada, Akihide Koyama, Hiroaki Nozaki, Yasuo Harigaya, Jin Nishimiya, et al. 2014. “Haploinsufficiency of CSF-1R and Clinicopathologic Characterization in Patients with HDLS.” Neurology 82 (2): 139–48. https://doi.org/10.1212/WNL.0000000000000046.

Lappalainen, Tuuli, Michael Sammeth, Marc R. Friedländer, Peter A.C. ‘t Hoen, Jean Monlong, Manuel A. Rivas, Mar Gonzàlez-Porta, et al. 2013. “Transcriptome and Genome Sequencing Uncovers Functional Variation in Humans.” Nature 501 (7468): 506–11. https://doi.org/10.1038/nature12531.

Lappalainen, Tuuli, Michael Sammeth, Marc R. Friedländer, Peter A.C. ‘T Hoen, Jean Monlong, Manuel A. Rivas, Mar Gonzàlez-Porta, et al. 2013. “Transcriptome and Genome Sequencing Uncovers Functional Variation in Humans.” Nature 501 (7468): 506–11. https://doi.org/10.1038/nature12531.

Levinson, Gene, and George A. Gutman. 1987. “High Frequencies of Short Frameshifts in Poly-CA/TG Tandem Repeats Borne by Bacteriophage M13 in Escherichia Coli K-12.” Nucleic Acids Research 15 (13): 5323–38. https://doi.org/10.1093/nar/15.13.5323.

Li, Ronghua, Anne Chomyn, and Min-Xin Guan. 2010. “Human Mitochondrial Leucyl-TRNA Synthetase Corrected Mitochondrial Dysfunctions Due to the MELAS and Diabetes Associated TRNA Leu(UUR) A3243G Mutation Running Title: TRNA Synthetase Corrects Mitochondrial Dysfunction Downloaded From.” Mol. Cell. Biol. https://doi.org/10.1128/MCB.01614-09.

Mancuso, Renzo, Gemma Fryatt, Madeleine Cleal, Juliane Obst, Elena Pipi, Jimena Monzón-Sandoval, Elena Ribe, et al. 2019. “CSF1R Inhibitor JNJ-40346527 Attenuates Microglial Proliferation and Neurodegeneration in P301S Mice.” Brain 142 (10): 3243–64. https://doi.org/10.1093/brain/awz241.

Mirkin, Sergei M. 2007. “Expandable DNA Repeats and Human Disease.” Nature 447 (7147): 932–40. https://doi.org/10.1038/nature05977.

Mitra, Ileena, Bonnie Huang, Nima Mousavi, Nichole Ma, Michael Lamkin, Richard Yanicky, Sharona Shleizer-Burko, Kirk E. Lohmueller, and Melissa Gymrek. 2021. “Patterns of de Novo Tandem Repeat Mutations and Their Role in Autism.” Nature 589 (7841): 246–50. https://doi.org/10.1038/s41586-020-03078-7.

Murphy, Erin E. 2015. “Inside the Cell.” In, 228–32. Nation Books.

Nicholson, Alexandra M., Matt C. Baker, Ni Cole A. Finch, Nicola J. Rutherford, Christian Wider, Neill R. Graff-Radford, Peter T. Nelson, et al. 2013. “CSF1R Mutations Link POLD and HDLS as a Single Disease Entity.” Neurology 80 (11): 1033–40. https://doi.org/10.1212/WNL.0b013e31828726a7.

Olmos-Alonso, Adrian, Sjoerd T. T. Schetters, Sarmi Sri, Katharine Askew, Renzo Mancuso, Mariana Vargas-Caballero, Christian Holscher, V. Hugh Perry, and Diego Gomez-Nicola. 2016. “Pharmacological Targeting of CSF1R Inhibits Microglial Proliferation and Prevents the Progression of Alzheimer’s-like Pathology.” Brain 139 (3): 891–907. https://doi.org/10.1093/brain/awv379.

Oosterhof, Nynke, Irene J. Chang, Ehsan Ghayoor Karimiani, Laura E. Kuil, Dana M. Jensen, Ray Daza, Erica Young, et al. 2019. “Homozygous Mutations in CSF1R Cause a Pediatric-Onset Leukoencephalopathy and Can Result in Congenital Absence of Microglia.” American Journal of Human Genetics 104 (5): 936–47. https://doi.org/10.1016/j.ajhg.2019.03.010.

Pierce, Sarah B., Ksenija Gersak, Rachel Michaelson-Cohen, Tom Walsh, Ming K. Lee, Daniel Malach, Rachel E. Klevit, Mary Claire King, and Ephrat Levy-Lahad. 2013. “Mutations in LARS2, Encoding Mitochondrial Leucyl-TRNA Synthetase, Lead to Premature Ovarian Failure and Hearing Loss in Perrault Syndrome.” American Journal of Human Genetics 92 (4): 614–20. https://doi.org/10.1016/j.ajhg.2013.03.007.

Quilez, Javier, Audrey Guilmatre, Paras Garg, Gareth Highnam, Melissa Gymrek, Yaniv Erlich, Ricky S. Joshi, David Mittelman, and Andrew J. Sharp. 2016. “Polymorphic Tandem Repeats within Gene Promoters Act as Modifiers of Gene Expression and DNA Methylation in Humans.” Nucleic Acids Research 44 (8): 3750–62. https://doi.org/10.1093/nar/gkw219.

Rademakers, Rosa, Matt Baker, Alexandra M. Nicholson, Nicola J. Rutherford, Nicole Finch, Alexandra Soto-Ortolaza, Jennifer Lash, et al. 2012. “Mutations in the Colony Stimulating Factor 1 Receptor (CSF1R) Gene Cause Hereditary Diffuse Leukoencephalopathy with Spheroids.” Nature Genetics 44 (2): 200–205. https://doi.org/10.1038/ng.1027.

Rosenbloom, Kate R., Cricket A. Sloan, Venkat S. Malladi, Timothy R. Dreszer, Katrina Learned, Vanessa M. Kirkup, Matthew C. Wong, et al. 2013. “ENCODE Data in the UCSC Genome Browser: Year 5 Update.” Nucleic Acids Research 41 (D1): D56–63. https://doi.org/10.1093/nar/gks1172.

Roth, Rachel, and Sara L. Ainsworth. 2015. “‘If They Hand You a Paper, You Sign It’: A Call to End the Sterilization of Women in Prison.” Hastings Women’s Law Journal 26:7–50.

Saini, Shubham, Ileena Mitra, Nima Mousavi, Stephanie Feupe Fotsing, and Melissa Gymrek. 2018. “A Reference Haplotype Panel for Genome-Wide Imputation of Short Tandem Repeats.” Nature Communications 9 (1): 4397. https://doi.org/10.1038/s41467-018-06694-0.

Shimamoto-Mitsuyama, Chie, Akihiro Nakaya, Kayoko Esaki, Shabeesh Balan, Yoshimi Iwayama, Tetsuo Ohnishi, Motoko Maekawa, Tomoko Toyota, Brian Dean, and Takeo Yoshikawa. 2021. “Lipid Pathology of the Corpus Callosum in Schizophrenia and the Potential Role of Abnormal Gene Regulatory Networks with Reduced Microglial Marker Expression.” Cerebral Cortex 31 (1): 448–62. https://doi.org/10.1093/cercor/bhaa236.

Soldà, Giulia, Sonia Caccia, Michela Robusto, Chiara Chiereghin, Pierangela Castorina, Umberto Ambrosetti, Stefano Duga, and Rosanna Asselta. 2016. “First Independent Replication of the Involvement of LARS2 in Perrault Syndrome by Whole-Exome Sequencing of an Italian Family.” Journal of Human Genetics 61 (4): 295–300. https://doi.org/10.1038/jhg.2015.149.

Sosna, Justyna, Stephan Philipp, Ricardo Iii Albay, Jorge Mauricio Reyes-Ruiz, David Baglietto-Vargas, Frank M. LaFerla, and Charles G. Glabe. 2018. “Early Long-Term Administration of the CSF1R Inhibitor PLX3397 Ablates Microglia and Reduces Accumulation of Intraneuronal Amyloid, Neuritic Plaque Deposition and Pre-Fibrillar Oligomers in 5XFAD Mouse Model of Alzheimer’s Disease.” Molecular Neurodegeneration 13 (1): 1–11. https://doi.org/10.1186/s13024-018-0244-x.

Srivastava, Prashant K., Jonathan van Eyll, Patrice Godard, Manuela Mazzuferi, Andree Delahaye-Duriez, Juliette Van Steenwinckel, Pierre Gressens, et al. 2018. “A Systems-Level Framework for Drug Discovery Identifies Csf1R as an Anti-Epileptic Drug Target.” Nature Communications 9 (1): 1–15. https://doi.org/10.1038/s41467-018-06008-4.

Storey, John D., and Robert Tibshirani. 2003. “Statistical Significance for Genomewide Studies.” Proceedings of the National Academy of Sciences 100 (16): 9440–45. https://doi.org/10.1073/PNAS.1530509100.

Takeichi, Takuya, Antonio Torrelo, John Y.W. Lee, Yusuke Ohno, María Luisa Lozano, Akio Kihara, Lu Liu, et al. 2017. “Biallelic Mutations in KDSR Disrupt Ceramide Synthesis and Result in a Spectrum of Keratinization Disorders Associated with Thrombocytopenia.” Journal of Investigative Dermatology 137 (11): 2344–53. https://doi.org/10.1016/j.jid.2017.06.028.

Tirosh, Itay, Naama Barkai, and Kevin J. Verstrepen. 2009. “Promoter Architecture and the Evolvability of Gene Expression.” Journal of Biology. BioMed Central. https://doi.org/10.1186/jbiol204.

Willems, Thomas, Melissa Gymrek, Gareth Highnam, The 1000 Genomes Project 1000 Genomes Project Consortium, David Mittelman, and Yaniv Erlich. 2014. “The Landscape of Human STR Variation.” Genome Research 24 (11): 1894–1904. https://doi.org/10.1101/gr.177774.114.

Willems, Thomas, Dina Zielinski, Jie Yuan, Assaf Gordon, Melissa Gymrek, and Yaniv Erlich. 2017. “Genome-Wide Profiling of Heritable and de Novo STR Variations.” Nature Methods 14 (6): 590–92. https://doi.org/10.1038/nmeth.4267.

Wyner, Nicole, Mark Barash, and Dennis McNevin. 2020. “Forensic Autosomal Short Tandem Repeats and Their Potential Association With Phenotype.” Frontiers in Genetics. Frontiers Media S.A. https://doi.org/10.3389/fgene.2020.00884.

Yun, Li, Cristen Willer, Serena Sanna, and Gonçalo Abecasis. 2009. “Genotype Imputation.” Annual Review of Genomics and Human Genetics. NIH Public Access. https://doi.org/10.1146/annurev.genom.9.081307.164242.

Zhang, Jiancheng, Lijia Chang, Yaoyu Pu, and Kenji Hashimoto. 2020. “Abnormal Expression of Colony Stimulating Factor 1 Receptor (CSF1R) and Transcription Factor PU.1 (SPI1) in the Spleen from Patients with Major Psychiatric Disorders: A Role of Brain–Spleen Axis.” Journal of Affective Disorders 272 (July): 110–15. https://doi.org/10.1016/j.jad.2020.03.128.

