## Supplemental Information for "Associations between forensic loci and neighboring gene expression levels may compromise medical privacy"

**Supplementary Information**

**Supplemental Table 1: Correlations between CODIS loci β values and the expression of neighboring genes.**

| CODIS STR | Gene | $r^2$ | *p* value | |
|---|---|---|---|---|
| CSF1PO | CSF1R | 0.010 | 0.033 | * |
| | PDGFRB | -0.002 | 0.708 | |
| | TIGD6 | 0.009 | 0.036 | * |
| | RPS20P4 | -0.003 | 0.805 | |
| | RPL7P1 | -0.003 | 0.934 | |
| | HMGXB3 | -0.003 | 0.916 | |
| | SLC26A2 | 0.001 | 0.231 | |
| D1S1656 | C1orf198 | 0.006 | 0.079 | |
| | COG2 | 0.003 | 0.159 | |
| D2S441 | C1D | 0.014 | 0.012 | * |
| D2S1338 | CXCR2P1 | -0.002 | 0.737 | |
| | RUFY4 | -0.003 | 0.822 | |
| | TNS1 | 0.000 | 0.290 | |
| D3S1358 | LIMD1 | -0.002 | 0.625 | |
| | LARS2 | 0.059 | 0.0000012 | * |
| D7S820 | SEMA3A | -0.003 | 0.851 | |
| D8S1179 | ZNF572 | 0.002 | 0.180 | |
| | RP11-1082L8 | -0.003 | 0.862 | |
| D12S391 | LOH12CR2 | -0.001 | 0.368 | |
| | MANSC1 | 0.002 | 0.203 | |
| | BORCS5 | -0.001 | 0.504 | |
| | LRP6 | -0.003 | 0.971 | |
| D18S51 | BCL2 | -0.0004 | 0.357 | |
| | KDSR | 0.011 | 0.025 | * |
| D19S433 | RPL9P32 | -0.003 | 0.790 | |

| | | | | |
|---|---|---|---|---|
| | URI1 | 0.003 | 0.147 | |
| D22S1045 | SSTR3 | -0.003 | 0.831 | |
| | IL2RB | -0.003 | 0.874 | |
| | C1QTNF6 | 0.000 | 0.353 | |
| | KCTD17 | -0.002 | 0.530 | |
| | Z82188 | -0.002 | 0.697 | |
| | TMPRSS6 | 0.003 | 0.162 | |
| | RP1-151B14.6 | -0.002 | 0.655 | |
| | RAC2 | 0.000 | 0.343 | |
| FGA | DCHS2 | 0.002 | 0.211 | |
| | PLRG1 | 0.011 | 0.026 | * |
| | RP11-158C21.3 | -0.001 | 0.421 | |
| TPOX | TPO | -0.003 | 0.935 | |
| vWA | VWF | 0.000 | 0.366 | |

* significant

**Supplemental Table 2: 1KG $\beta$ Regression Summaries, Population as Covariate Results, FDR.** Table of linear regression summary statistics for the CODIS loci $\beta$ statistic. vs gene expression levels of neighboring genes. Summaries for cumulative and within population linear model statistics are provided. After correcting for multiple testing, the threshold of significance in the cumulative analysis is 0.0013. In the within population analysis, performed on the cumulative significant and marginally significant observations, the corrected *p* value is 0.0016.

**Supplemental Figure 1: 1000 Genomes correlation plots between CODISeSTR β values and neighboring gene expression levels.**

Linear regression models fitted to test β (beta) association with gene expression levels in the CODIS loci using cumulative 1000 Genomes data. The black dots show the STR β value per individual (x-axis) and their corresponding RPKM (y-axis). The blue line is the best fitted linear model. Shaded blue region illustrates a 95% confidence interval on the fitted values. Results are shown for (A) D1S1656, (B) D2S441, (C) vWA, (D) CSF1PO, (E) D2S1338, (F) D3S1358, (G) D7S820, (H) TPOX, (I) D8S1179, (J) D18S51, (K) D12S391, (L) D19S433, (M) D22S1045, and (N) FGA.

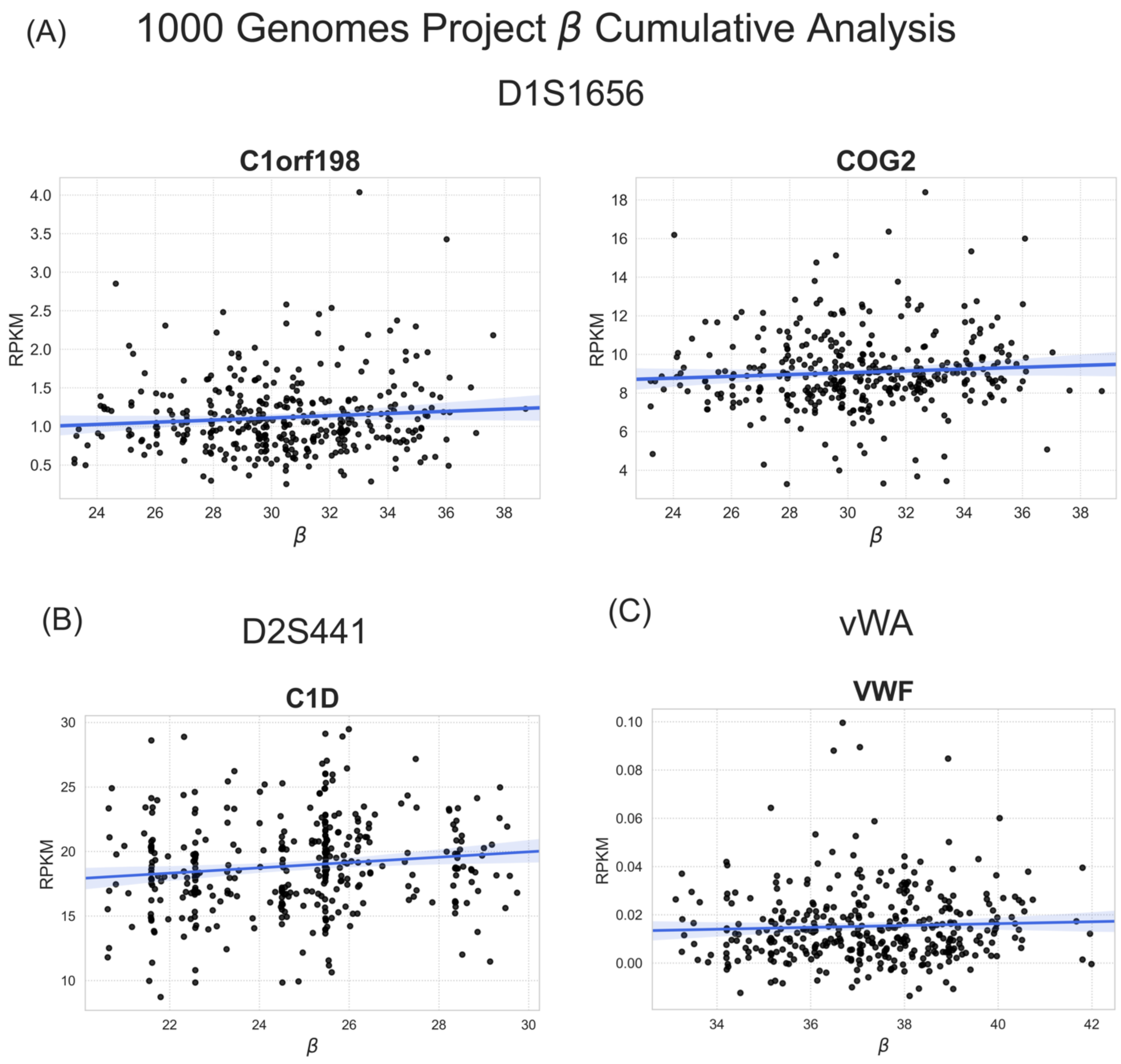

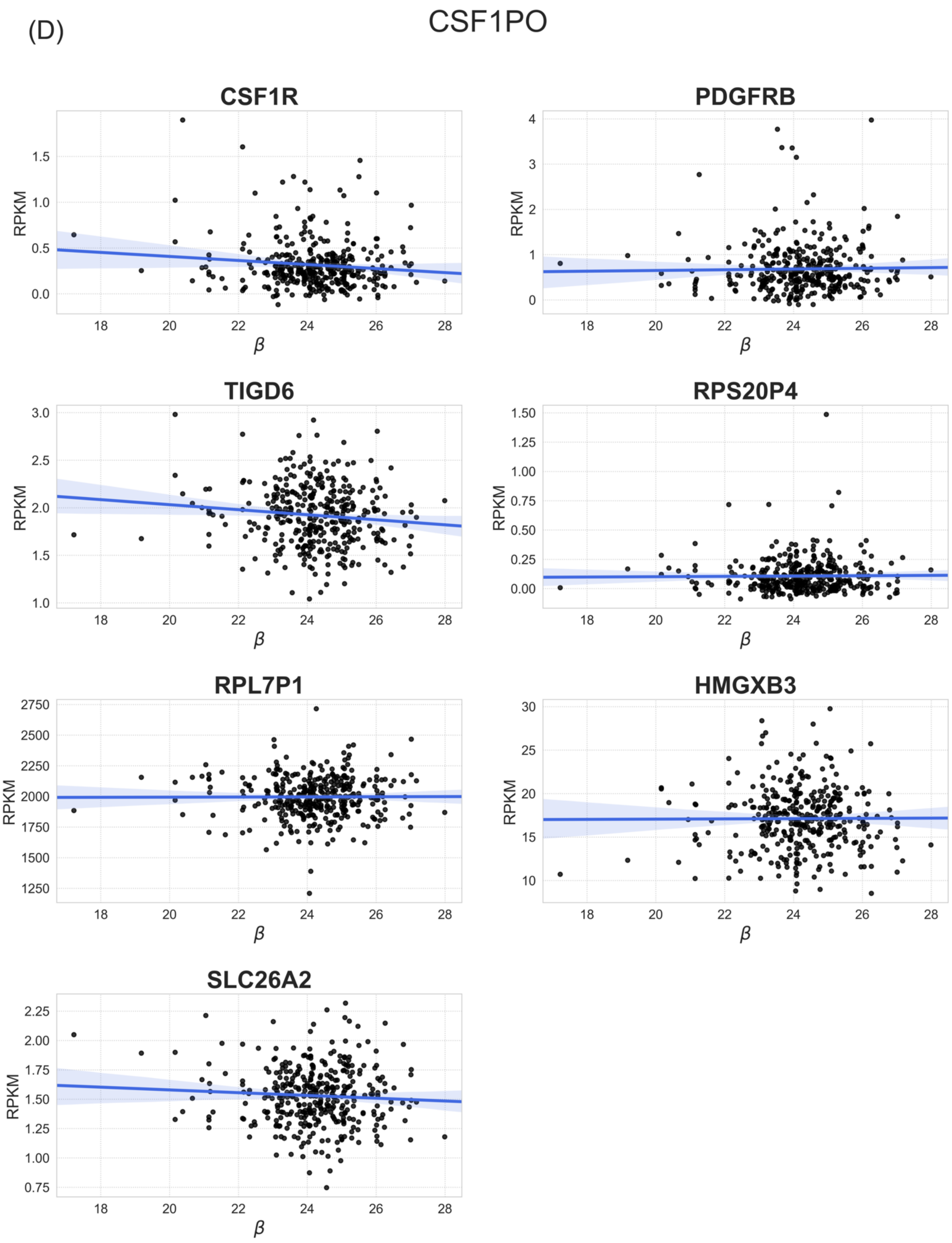

(D)
CSF1PO
CSF1R
PDGFRB
TIGD6
RPS20P4
RPL7P1
HMGXB3
SLC26A2
RPKM
β

(E)

D2S1338

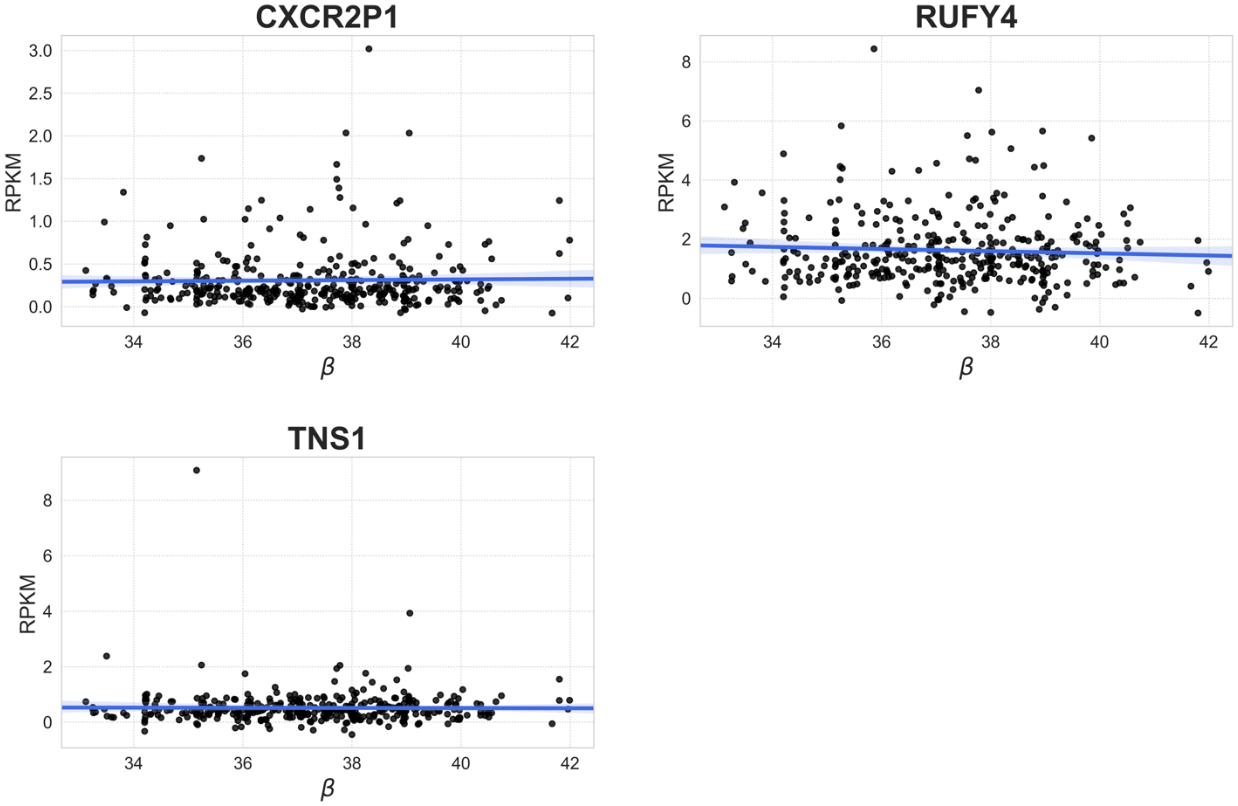

(F)

D3S1358

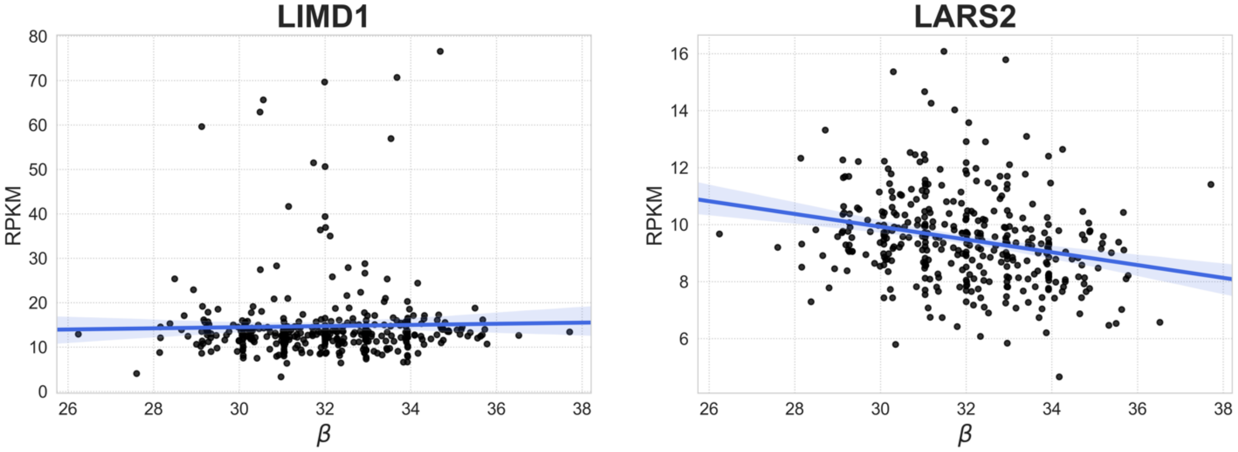

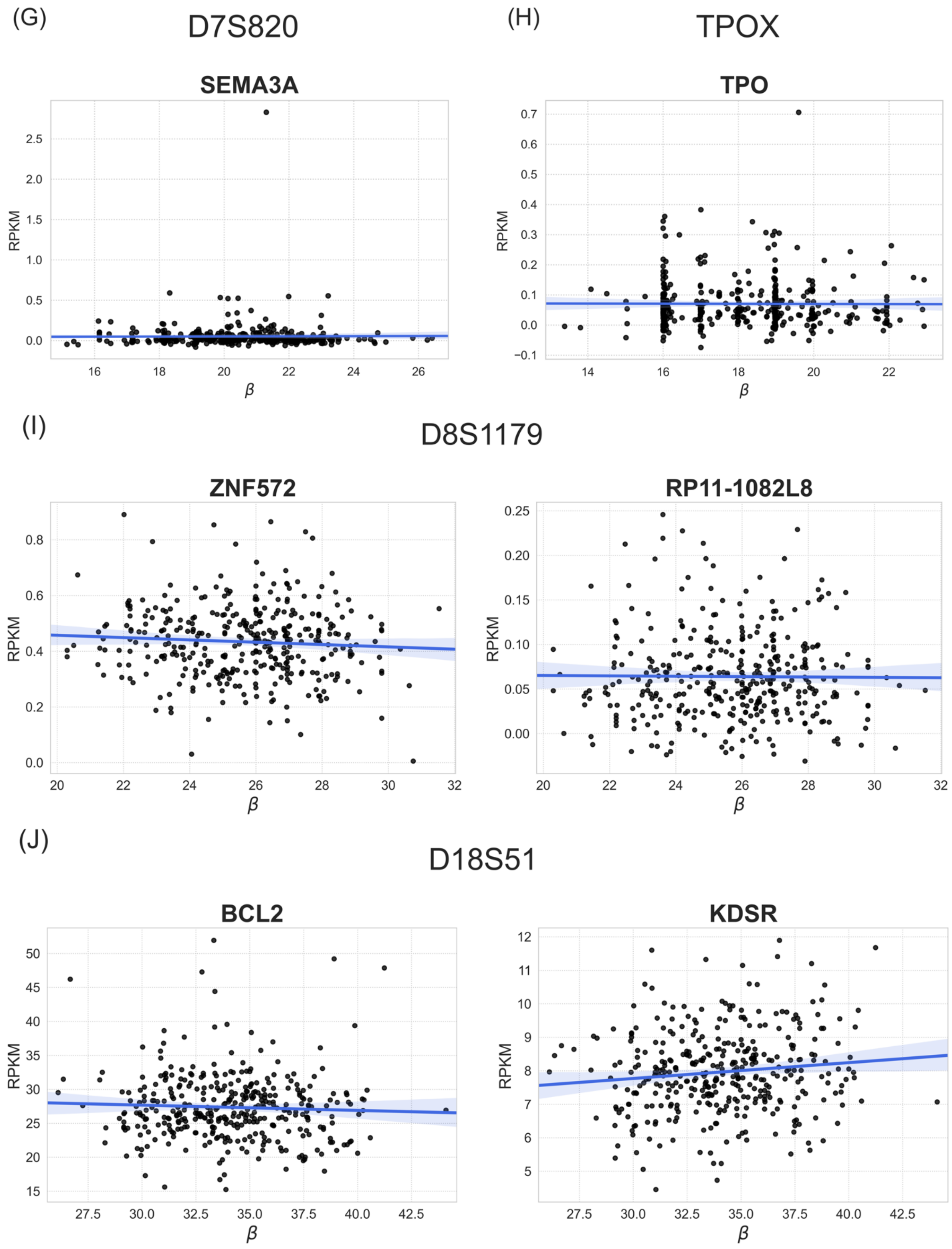
(G)
D7S820
SEMA3A
RPKM
β
(H)
TPOX
TPO
RPKM
β
(I)
D8S1179
ZNF572
RPKM
β
RP11-1082L8
RPKM
β
(J)
D18S51
BCL2
RPKM
β
KDSR
RPKM
β

(K)

D12S391

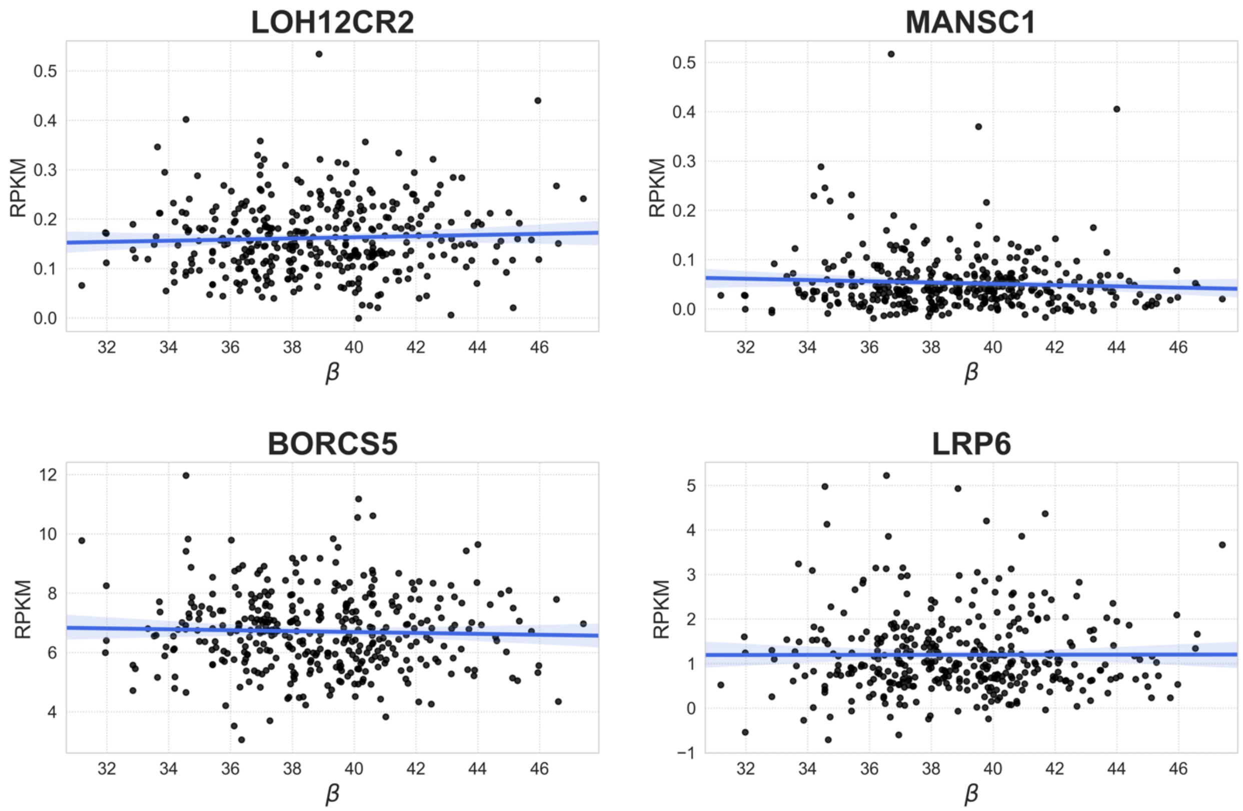

(L)

D19S433

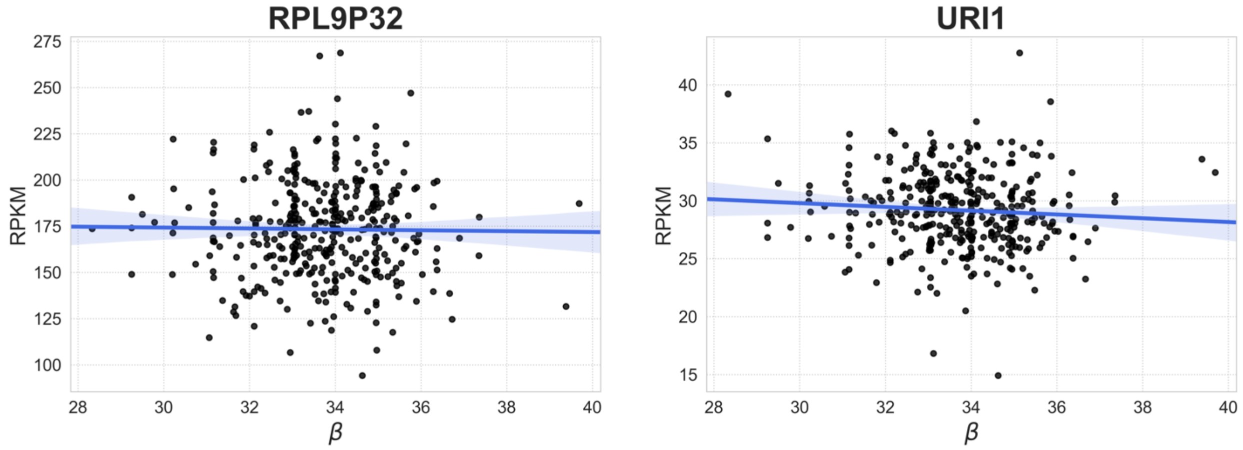

(M)

D22S1045

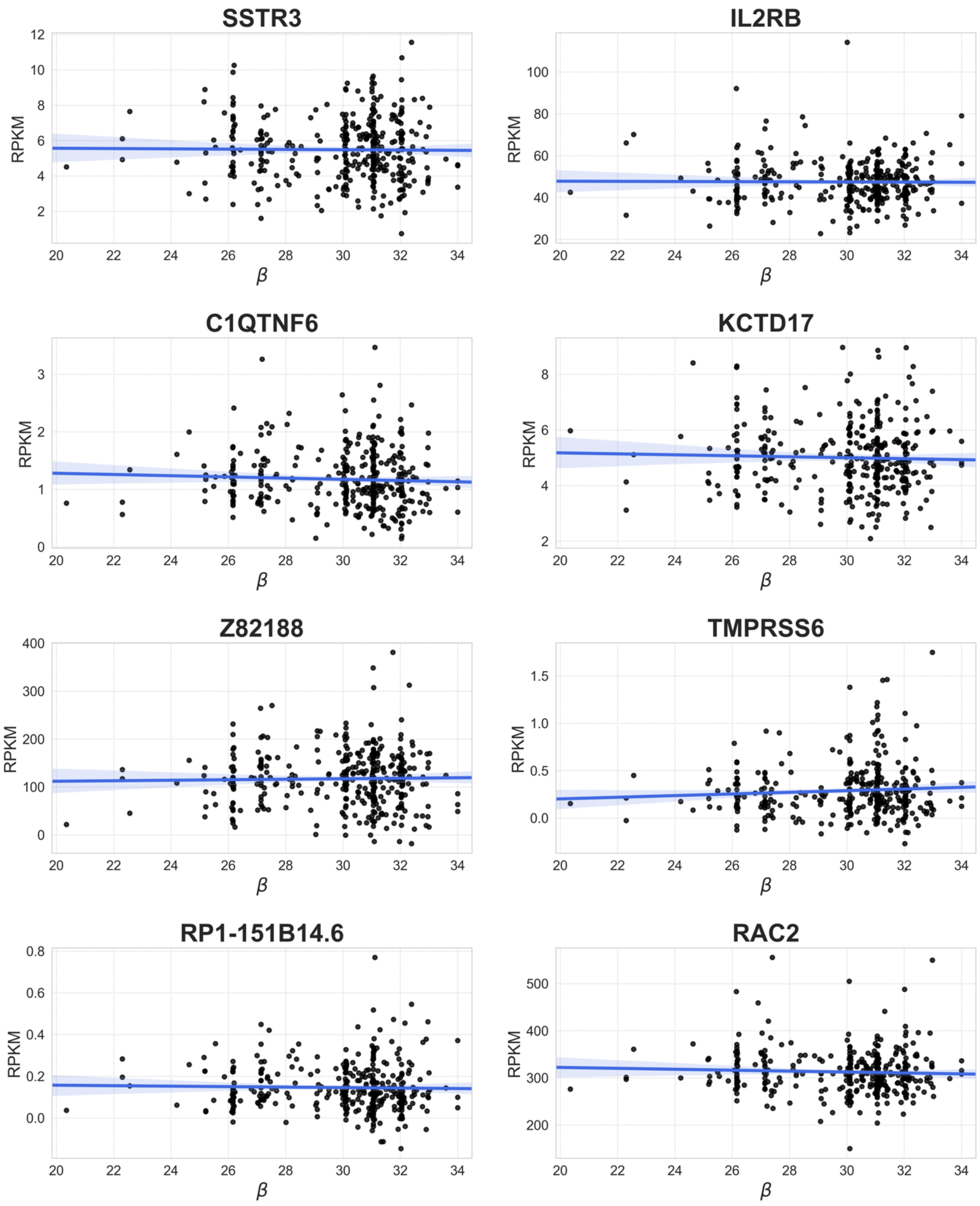

SSTR3
IL2RB
C1QTNF6
KCTD17
Z82188
TMPRSS6
RP1-151B14.6
RAC2
RPKM
β

(N)

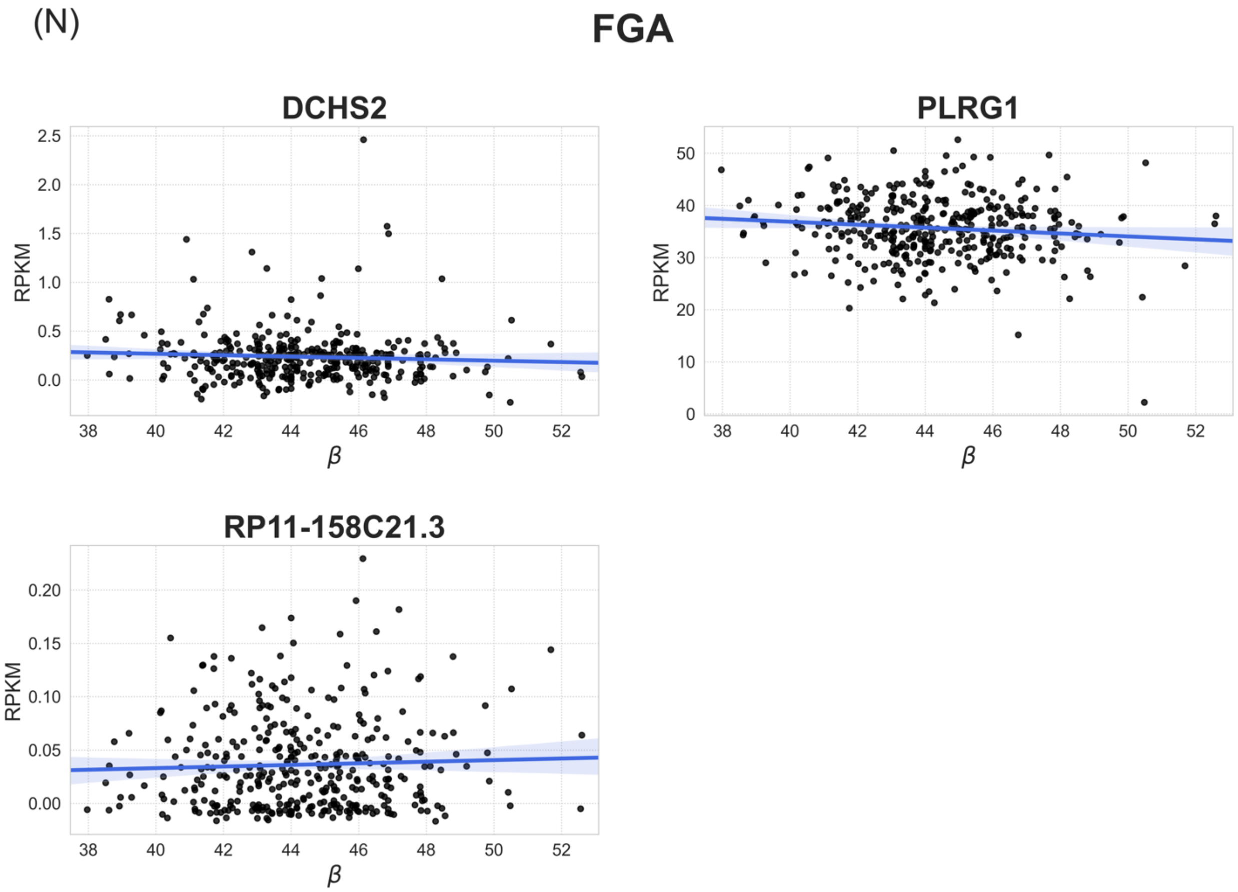

**Supplemental Figure 2: By population correlation plots fitted to b (beta) values vs RPKM levels in significant and marginally significant associations in STR-gene pairs.**

The black dots show the STR β (beta) genotype per individual (x-axis) and their corresponding RPKM (y-axis). The blue line demonstrates the best fitted linear model. Shaded blue region illustrates a 95% confidence interval on the fitted values. Results are shown for (A) CSF1PO-*CSF1R*, (B) CSF1PO-*TIGD6*, (C) D18S51-*KDSR*, (D) D2S441-*C1D*, (E) D3S1358-*LARS2* and (F) FGA- *PLGR1*.

(A)

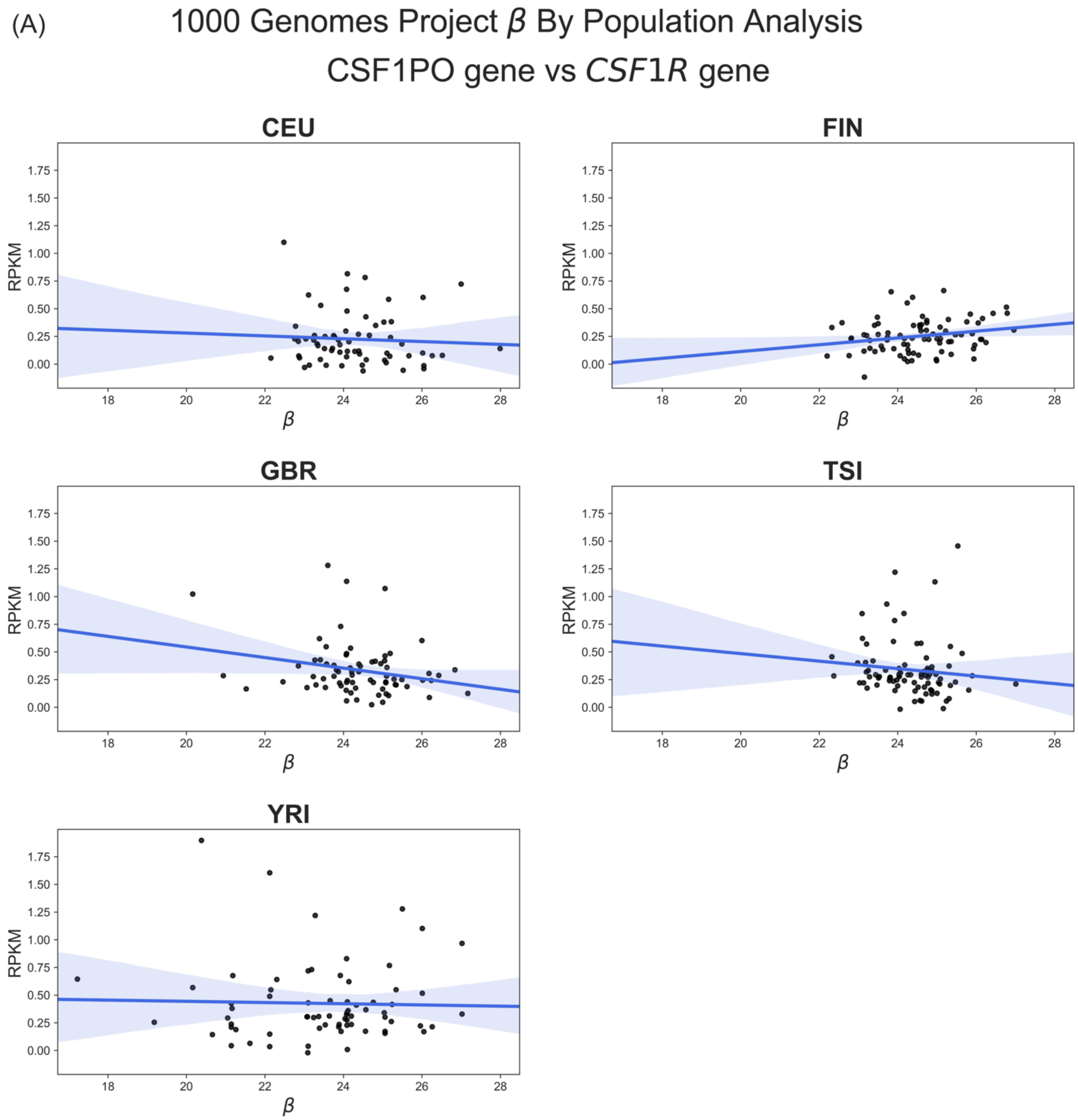

(B) CSF1PO locus vs *TIGD6* gene

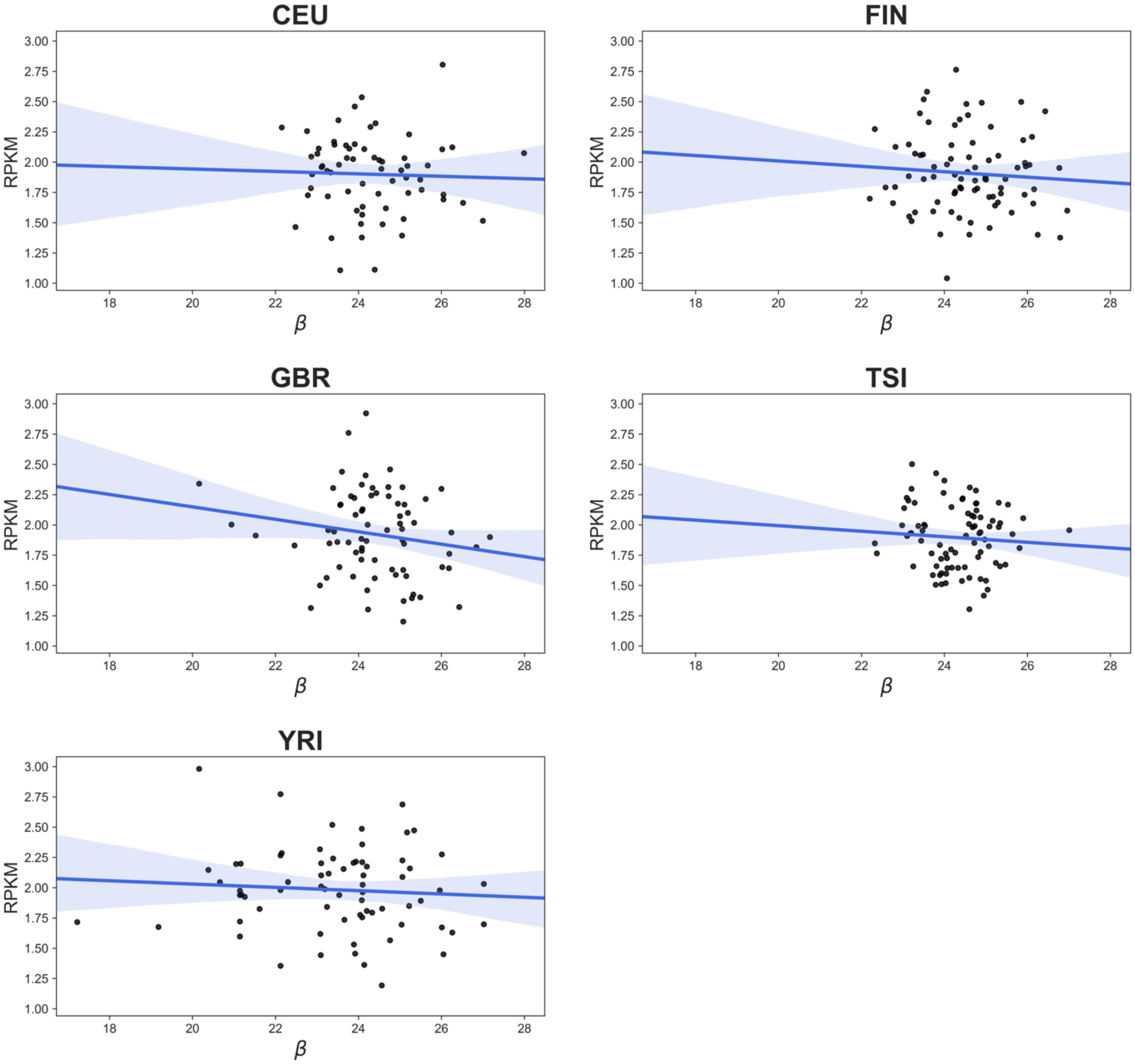

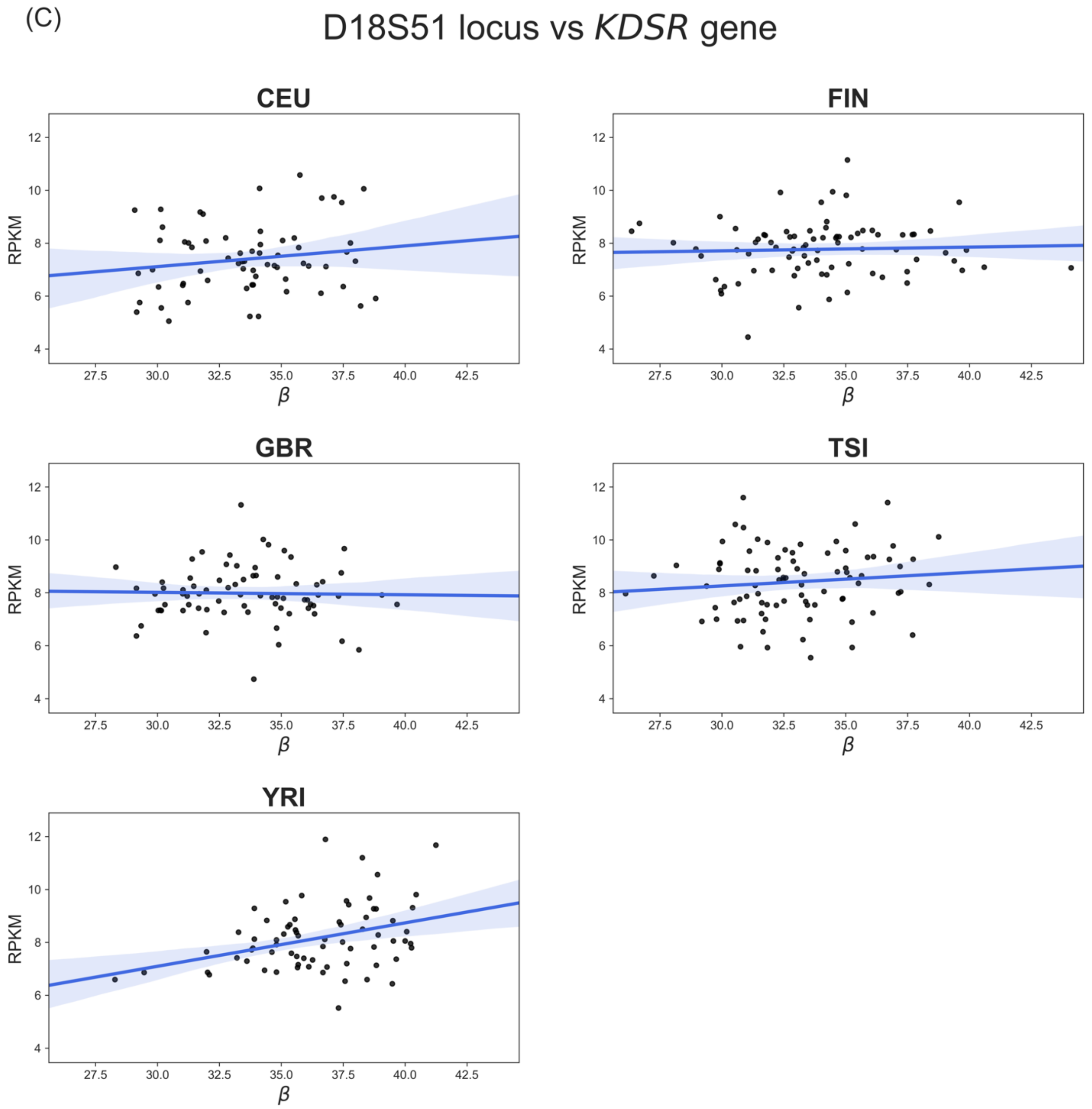
(C)
D18S51 locus vs KDSR gene
CEU
FIN
GBR
TSI
YRI
RPKM
β
12
10
8
6
4
27.5
30.0
32.5
35.0
37.5
40.0
42.5

(D) D2S441 locus vs *C1D* gene

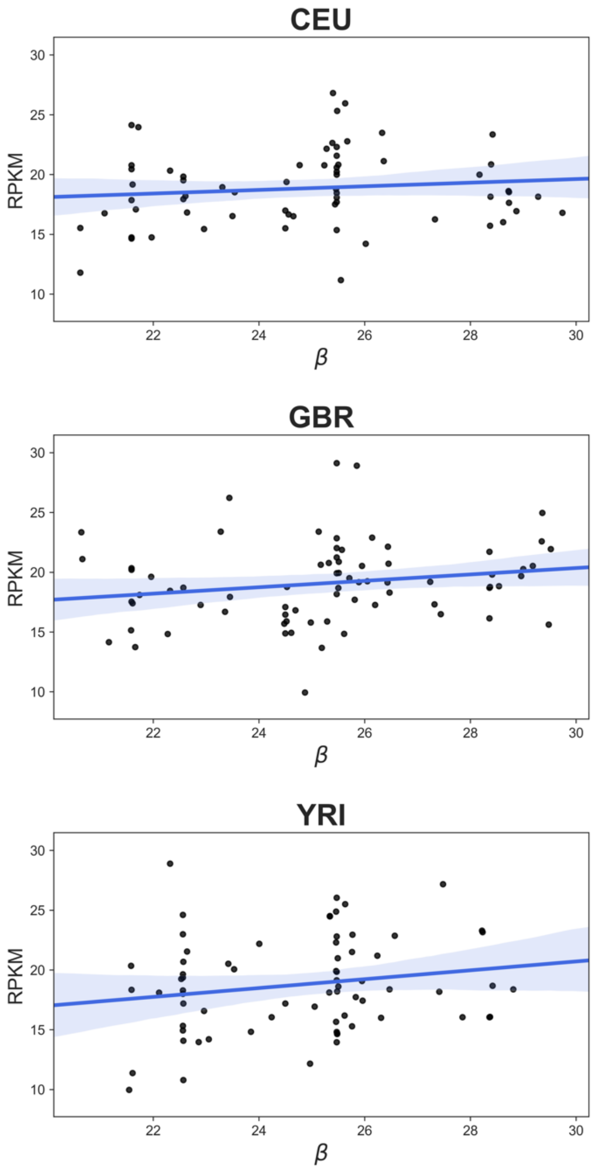

(E) D3S1358 locus vs *LARS2* gene

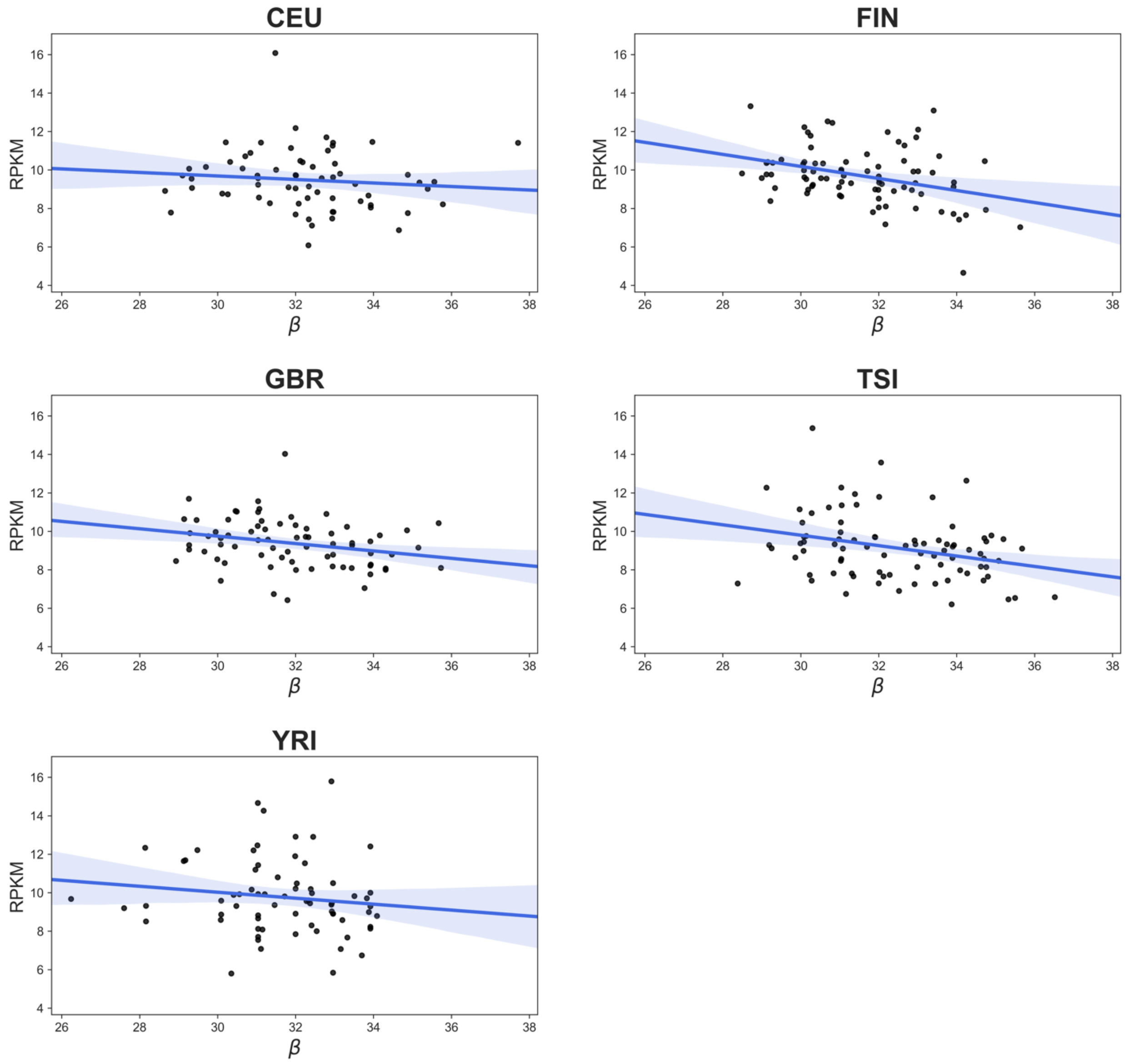

(F) FGA locus vs *PLRG1* gene

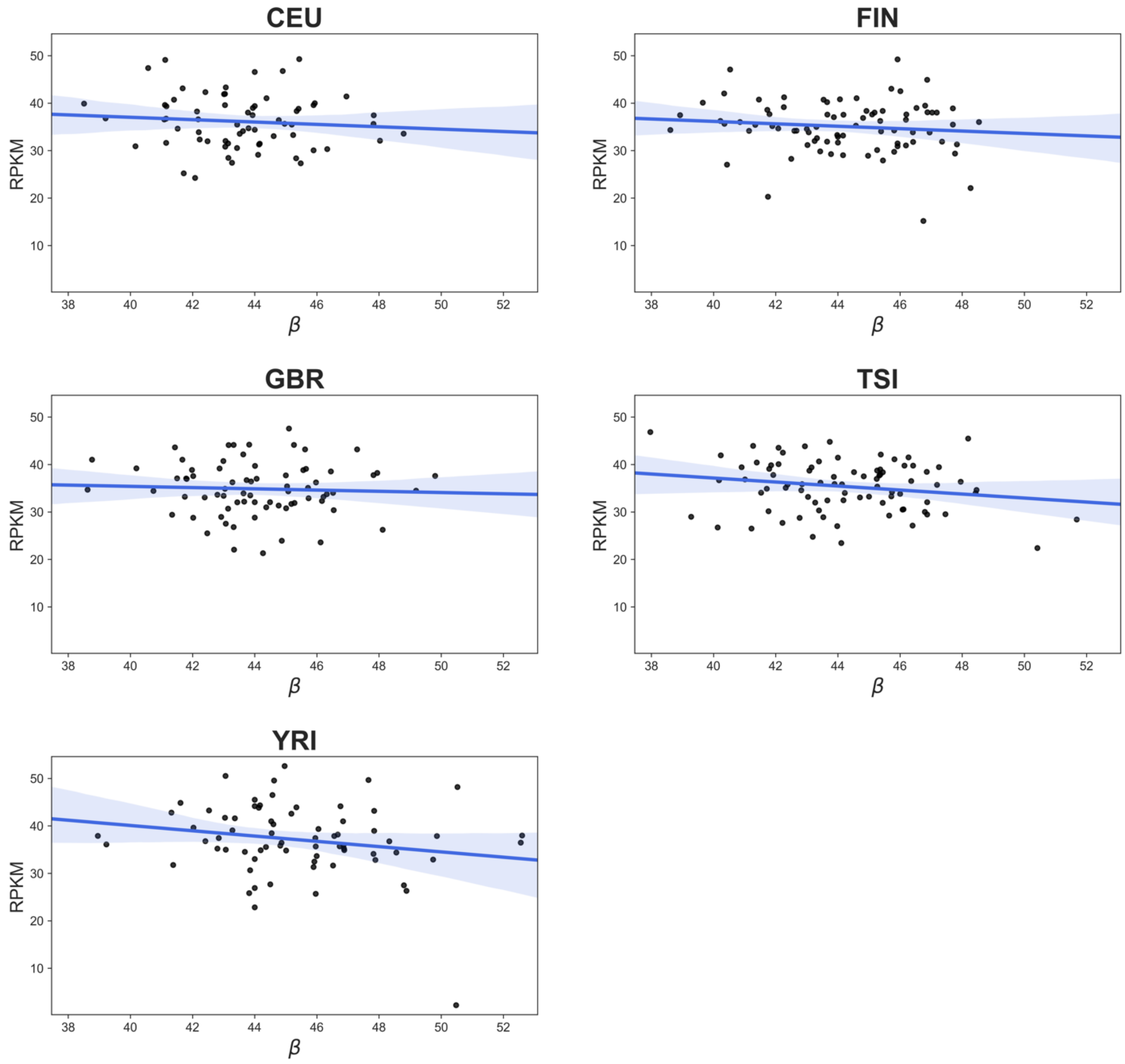

**Supplemental Table 3: Subpopulation-based correlations between CODISeSTRs and associated genes in the 1000 Genomes data.**

| CODIS STR | Gene | Population | $r^2$ | *p* value | |
|---|---|---|---|---|---|
| **CSF1PO** | CSF1R | CEU | -0.0119 | 0.622 | |
| | | FIN | 0.0385 | 0.044 | * |
| | | GBR | 0.0439 | 0.042 | * |
| | | TSI | 0.0001 | 0.318 | |
| | | YRI | -0.0139 | 0.813 | |
| **CSF1PO** | TIGD6 | CEU | -0.0145 | 0.775 | |
| | | FIN | -0.0069 | 0.504 | |
| | | GBR | 0.0167 | 0.140 | |
| | | TSI | -0.0068 | 0.505 | |
| | | YRI | -0.0091 | 0.542 | |
| **D2S441** | C1D | CEU | -0.0018 | 0.350 | |
| | | FIN | -0.0121 | 0.830 | |
| | | GBR | 0.0222 | 0.109 | |
| | | TSI | 0.0048 | 0.241 | |
| | | YRI | 0.0197 | 0.127 | |
| **D3S1358** | LARS2 | CEU | -0.0042 | 0.396 | |
| | | FIN | 0.1118 | 0.001 | * |
| | | GBR | 0.0560 | 0.025 | * |
| | | TSI | 0.0809 | 0.005 | * |
| | | YRI | 0.0037 | 0.266 | |
| **D18S51** | KDSR | CEU | 0.0103 | 0.202 | |
| | | FIN | -0.0108 | 0.703 | |
| | | GBR | -0.0136 | 0.848 | |
| | | TSI | -0.0009 | 0.338 | |
| | | YRI | 0.1112 | 0.003 | * |
| **FGA** | PLRG1 | CEU | -0.0071 | 0.461 | |
| | | FIN | -0.0001 | 0.323 | |
| | | GBR | -0.0115 | 0.672 | |
| | | TSI | 0.0267 | 0.075 | |
| | | YRI | 0.0258 | 0.097 | |

*significant

**Supplemental Table 4: Two-tailed Kolmogorov-Smirnov test CODIS STR b values by subpopulation and gene expression.**

D2S441 β subpopulation pairwise comparison using two-tailed Kolmogorov-Smirnov test.

| | FIN | GBR | CEU | TSI |
|---|---|---|---|---|
| **YRI** | 0.338 | 0.468 | 0.487 | 0.531 |
| **FIN** | | 0.040* | 0.371 | 0.299 |
| **GBR** | | | 0.456 | 0.457 |
| **CEU** | | | | 0.907 |

*C1D* gene expression subpopulation pairwise comparison using two-tailed Kolmogorov-Smirnov test.

| | FIN | GBR | CEU | TSI |
|---|---|---|---|---|
| **YRI** | 0.544 | 0.500 | 0.566 | 0.139 |
| **FIN** | | 0.686 | 0.602 | 0.560 |
| **GBR** | | | 0.708 | 0.844 |
| **CEU** | | | | 0.898 |

D3S1358 β subpopulation pairwise comparison using two-tailed Kolmogorov-Smirnov test for all subpopulations.

| | FIN | GBR | CEU | TSI |
|---|---|---|---|---|
| **YRI** | 0.237 | 0.655 | 0.393 | 0.023* |
| **FIN** | | 0.641 | 0.107 | 0.019* |
| **GBR** | | | 0.204 | 0.264 |
| **CEU** | | | | 0.178 |

*LARS2* gene expression subpopulation pairwise comparison using two-tailed Kolmogorov-Smirnov test for all subpopulations.

| | FIN | GBR | CEU | TSI |
|---|---|---|---|---|
| **YRI** | 0.569 | 0.136 | 0.356 | 0.036* |
| **FIN** | | 0.318 | 0.475 | 0.009** |
| **GBR** | | | 0.842 | 0.056 |
| **CEU** | | | | 0.118 |

D18S51 β subpopulation pairwise comparison using two-tailed Kolmogorov-Smirnov test for all subpopulations.

| | FIN | GBR | CEU | TSI |
|---|---|---|---|---|
| **YRI** | 4e-7** | 3e-7** | 1e-6** | 2e-10** |
| **FIN** | | 0.784 | 0.945 | 0.054 |
| **GBR** | | | 0.972 | 0.344 |
| **CEU** | | | | 0.063 |

*KDSR* gene expression subpopulation pairwise comparison using two-tailed Kolmogorov-Smirnov test for all subpopulations.

| | FIN | GBR | CEU | TSI |
|---|---|---|---|---|
| **YRI** | 0.103 | 0.492 | 0.004** | 0.111 |
| **FIN** | | 0.167 | 0.021 | 1e-5** |
| **GBR** | | | 3e-5** | 0.008** |
| **CEU** | | | | 1e-5** |

CSF1PO $\beta$ pairwise subpopulation comparison using two-tailed Kolmogorov-Smirnov test for all subpopulations.

| | FIN | GBR | CEU | TSI |
|---|---|---|---|---|
| **YRI** | 9e-5** | 0.002** | 0.015 | 0.002** |
| **FIN** | | 0.485 | 0.066 | 0.15 |
| **GBR** | | | 0.561 | 0.323 |
| **CEU** | | | | 0.586 |

*TIGD6* gene expression subpopulation pairwise comparison using two-tailed Kolmogorov-Smirnov test for all subpopulations.

| | FIN | GBR | CEU | TSI |
|---|---|---|---|---|
| **YRI** | 0.188 | 0.657 | 0.222 | 0.256 |
| **FIN** | | 0.550 | 0.572 | 0.753 |
| **GBR** | | | 0.393 | 0.401 |
| **CEU** | | | | 0.823 |

*CSF1R* gene expression subpopulation pairwise comparison using two-tailed Kolmogorov-Smirnov test for all subpopulations.

| | FIN | GBR | CEU | TSI |
|---|---|---|---|---|
| **YRI** | 0.010* | 0.247 | 2e-5** | 0.054 |
| **FIN** | | 0.310 | 0.028* | 0.107 |
| **GBR** | | | 2e-4** | 0.835 |
| **CEU** | | | | 5e-5** |

FGA β pairwise subpopulation comparison using two-tailed Kolmogorov-Smirnov test for all subpopulations.

| | FIN | GBR | CEU | TSI |
|---|---|---|---|---|
| **YRI** | 0.121 | 0.022* | 0.001** | 0.043* |
| **FIN** | | 0.543 | 0.099 | 0.996 |
| **GBR** | | | 0.669 | 0.610 |
| **CEU** | | | | 0.128 |

*PLRG1* gene expression subpopulation pairwise comparison using two-tailed Kolmogorov-Smirnov test for all subpopulations.

| | FIN | GBR | CEU | TSI |
|---|---|---|---|---|
| **YRI** | 0.080 | 0.031* | 0.309 | 0.189 |
| **FIN** | | 0.737 | 0.472 | 0.594 |
| **GBR** | | | 0.551 | 0.809 |
| **CEU** | | | | 0.891 |

**Supplemental Figure 3: Histograms of subpopulation CODISeSTR β values and associated gene expression levels.**

The empirical distributions of CODIS loci β values and associated genes for which a significant or marginally significant association was observed in the within population analysis are plotted. Results shown for (A) D2S441, (B) *C1D*, (C) D3S1358, (D) *LARS2*, (E) D18S51, (F) *KDSR*, (G) CSF1PO, (H) *CSF1R*, (I) *TIGD6*, (J) FGA and (K) *PLRG1*.

(A)

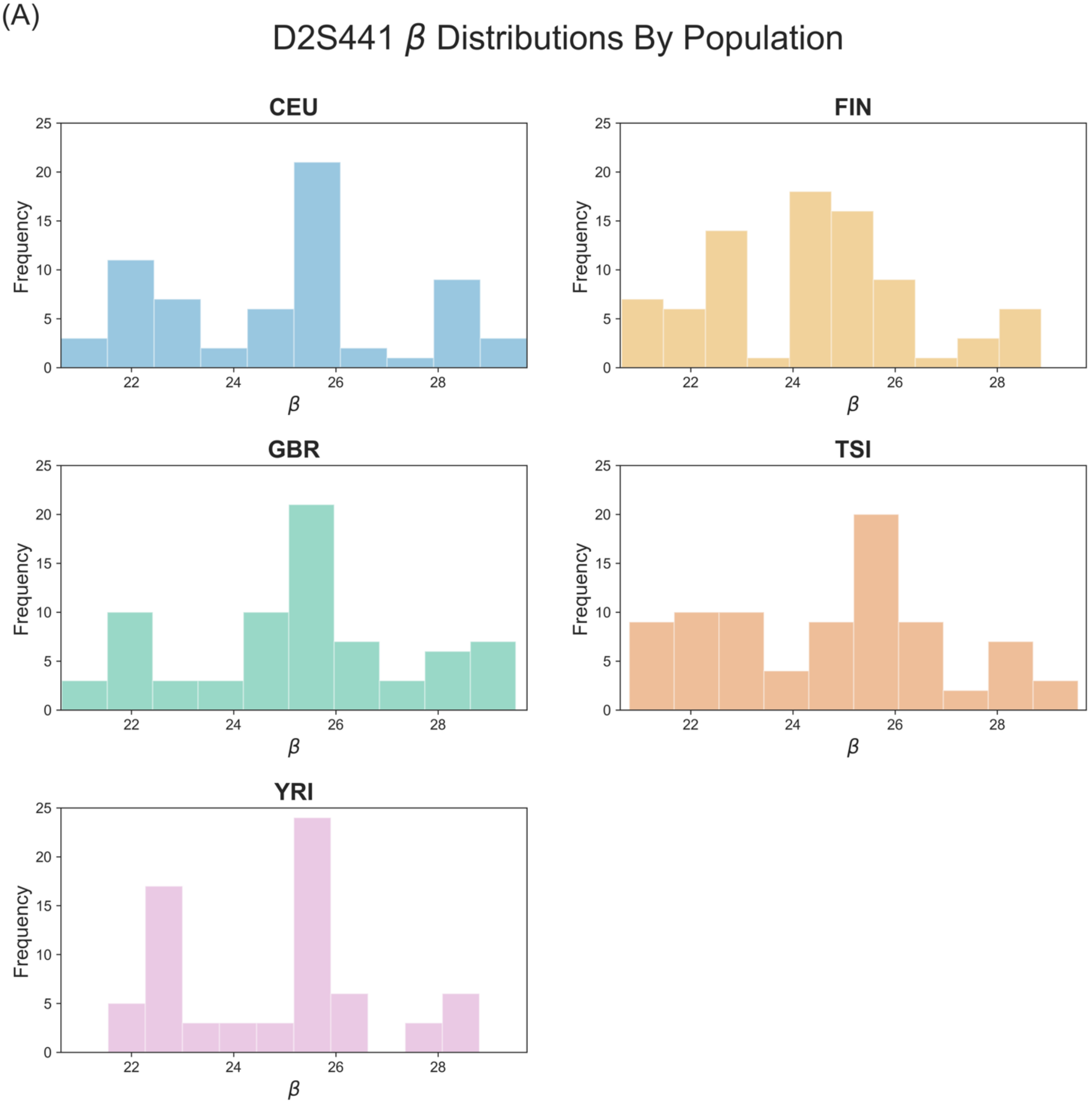

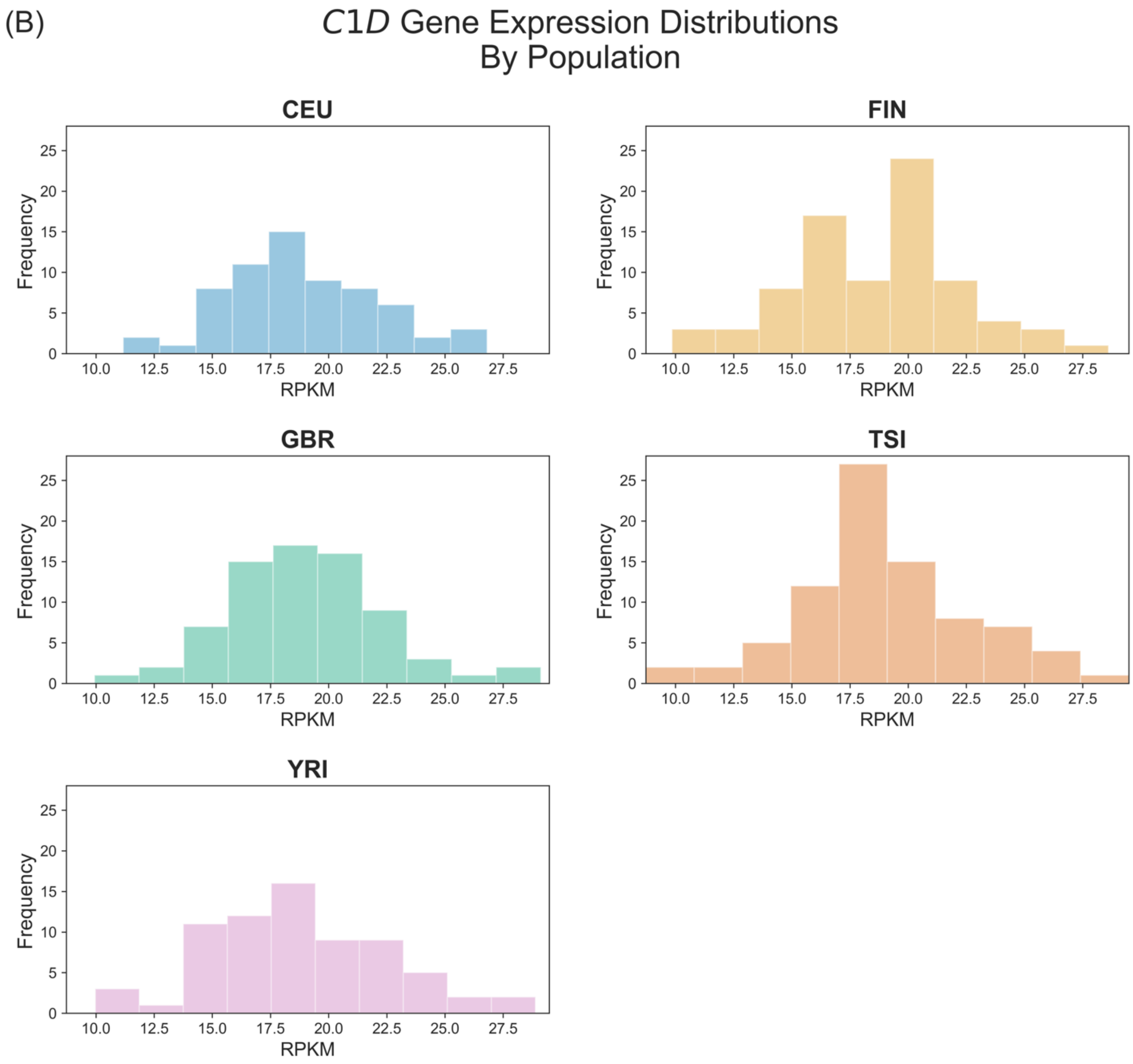

(B)
C1D Gene Expression Distributions
By Population
CEU
FIN
GBR
TSI
YRI
Frequency
RPKM
0
5
10
15
20
25
10.0
12.5
15.0
17.5
20.0
22.5
25.0
27.5

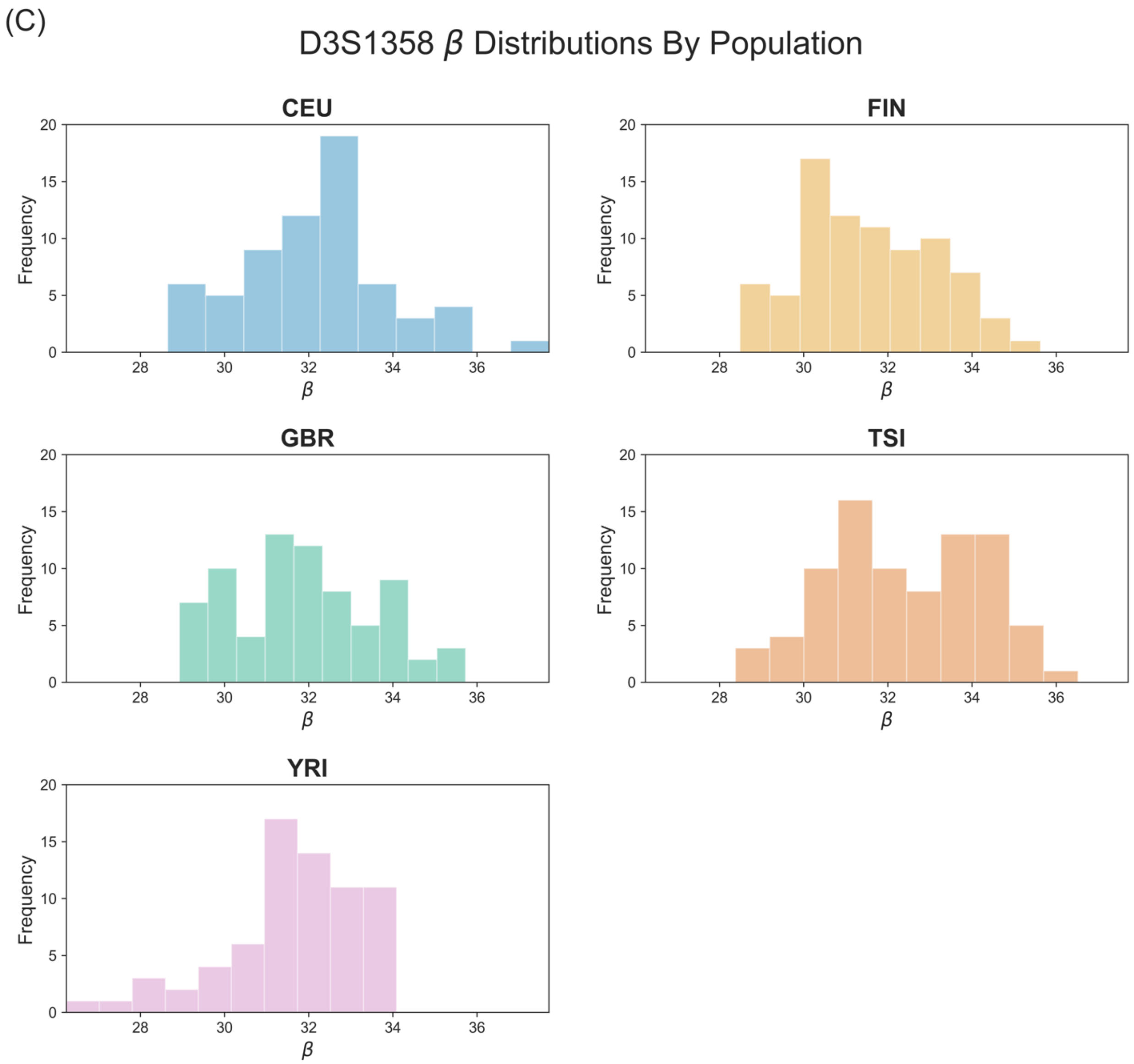
(C)
D3S1358 β Distributions By Population
CEU
FIN
GBR
TSI
YRI
Frequency
β

(D) *LARS2* Gene Expression Distributions By Population

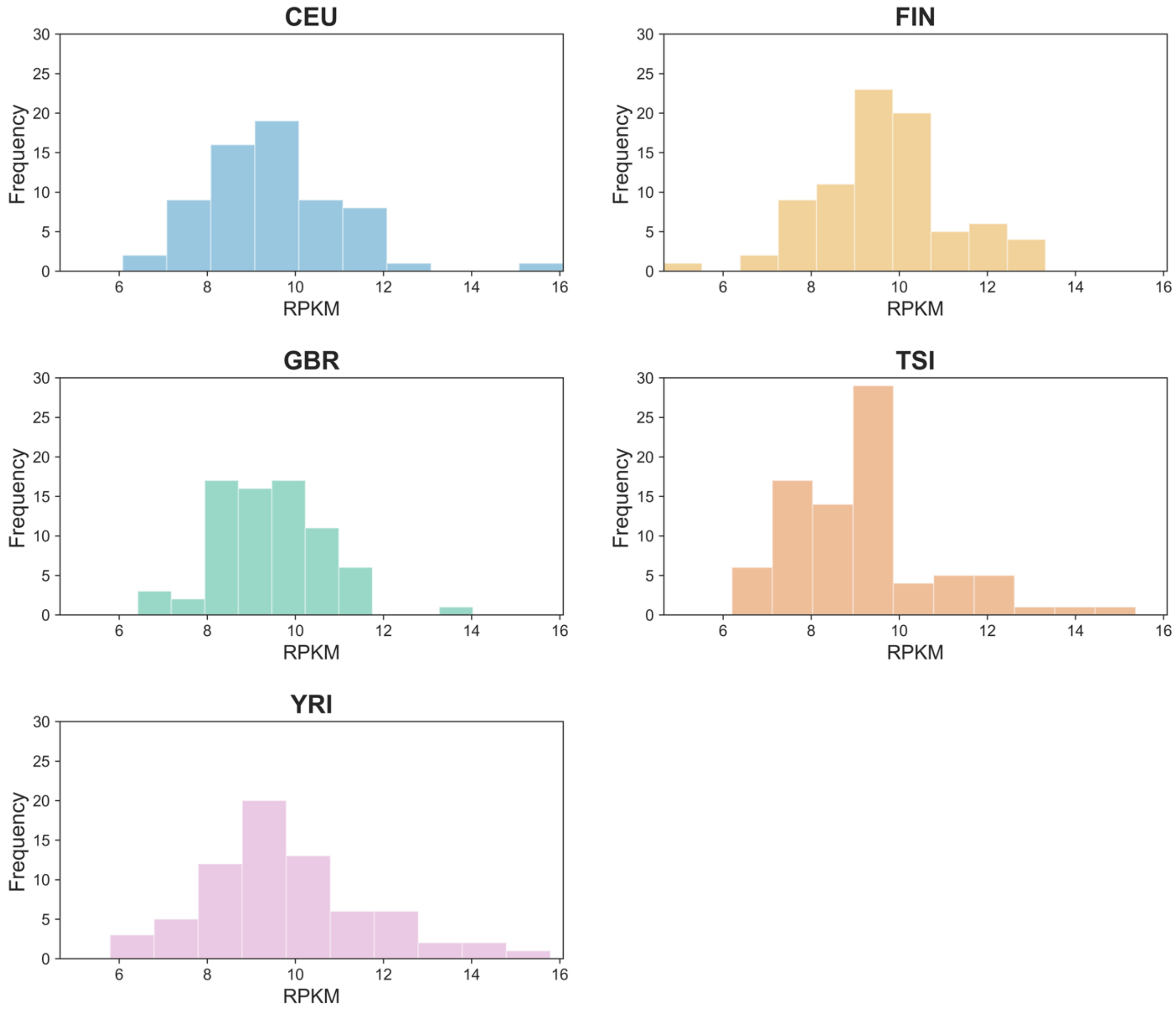

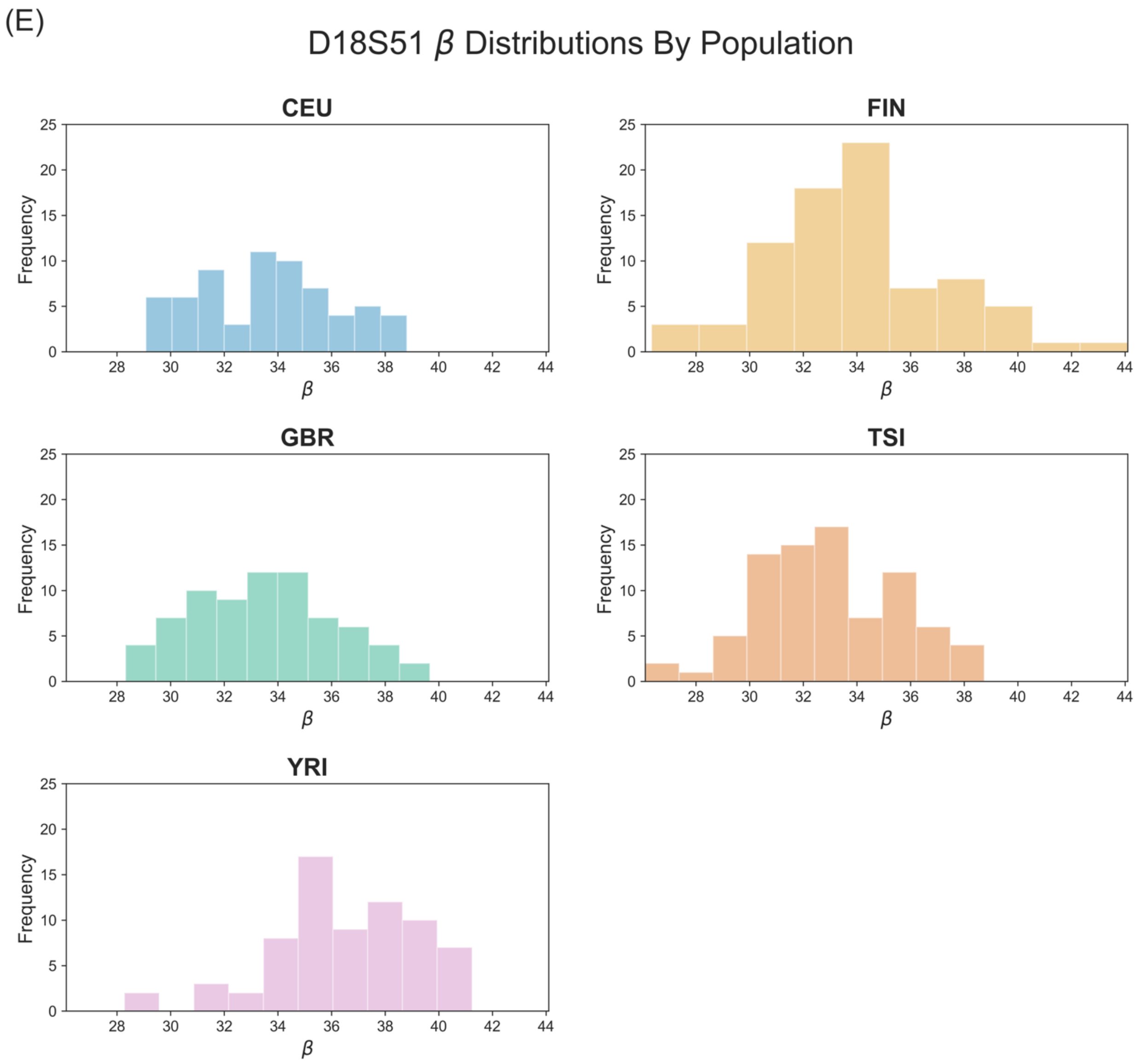

(E)
D18S51 β Distributions By Population
CEU
FIN
GBR
TSI
YRI
Frequency
β
0
5
10
15
20
25
28
30
32
34
36
38
40
42
44

(F) *KDSR* Gene Expression Distributions By Population

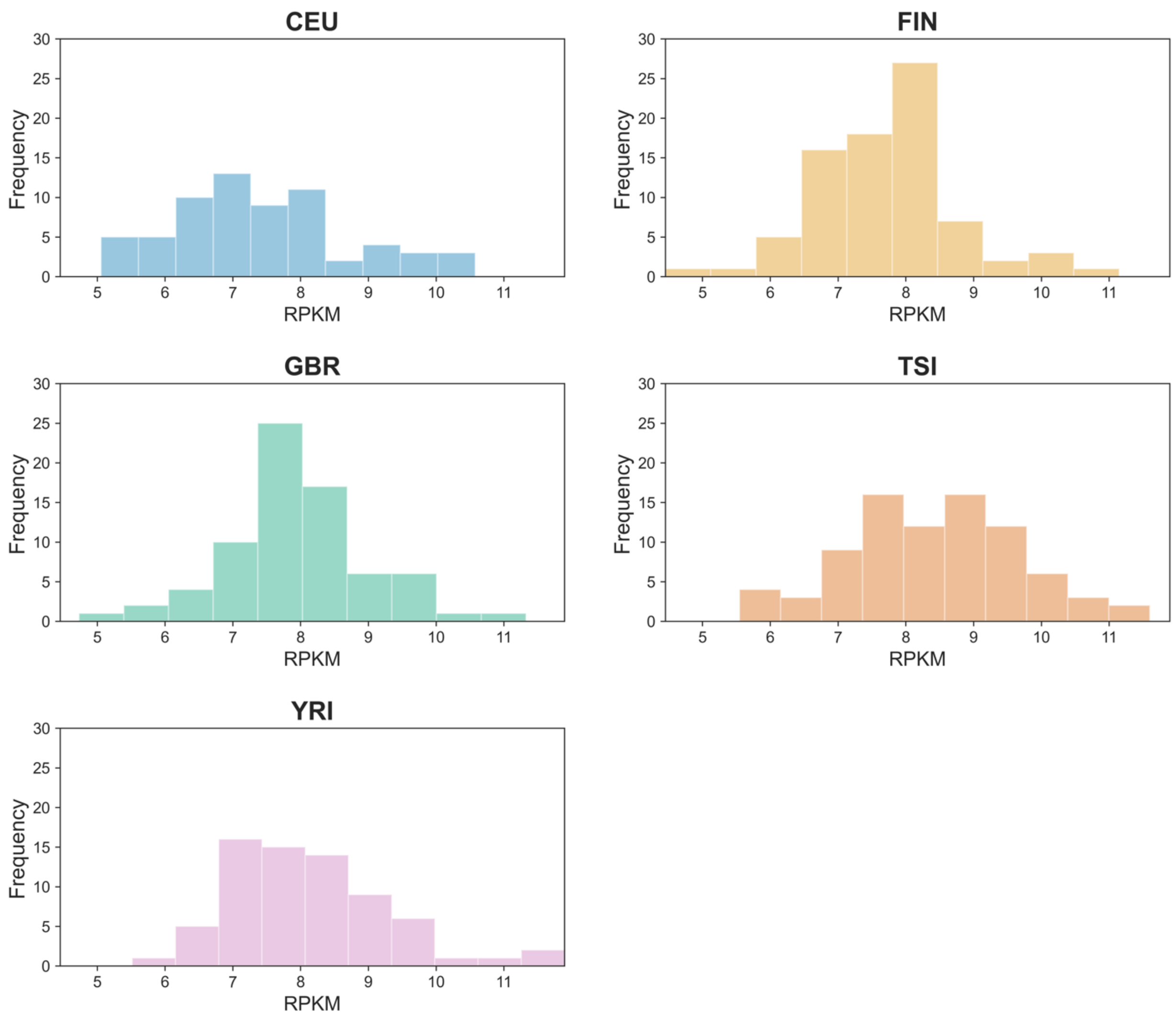

(G)

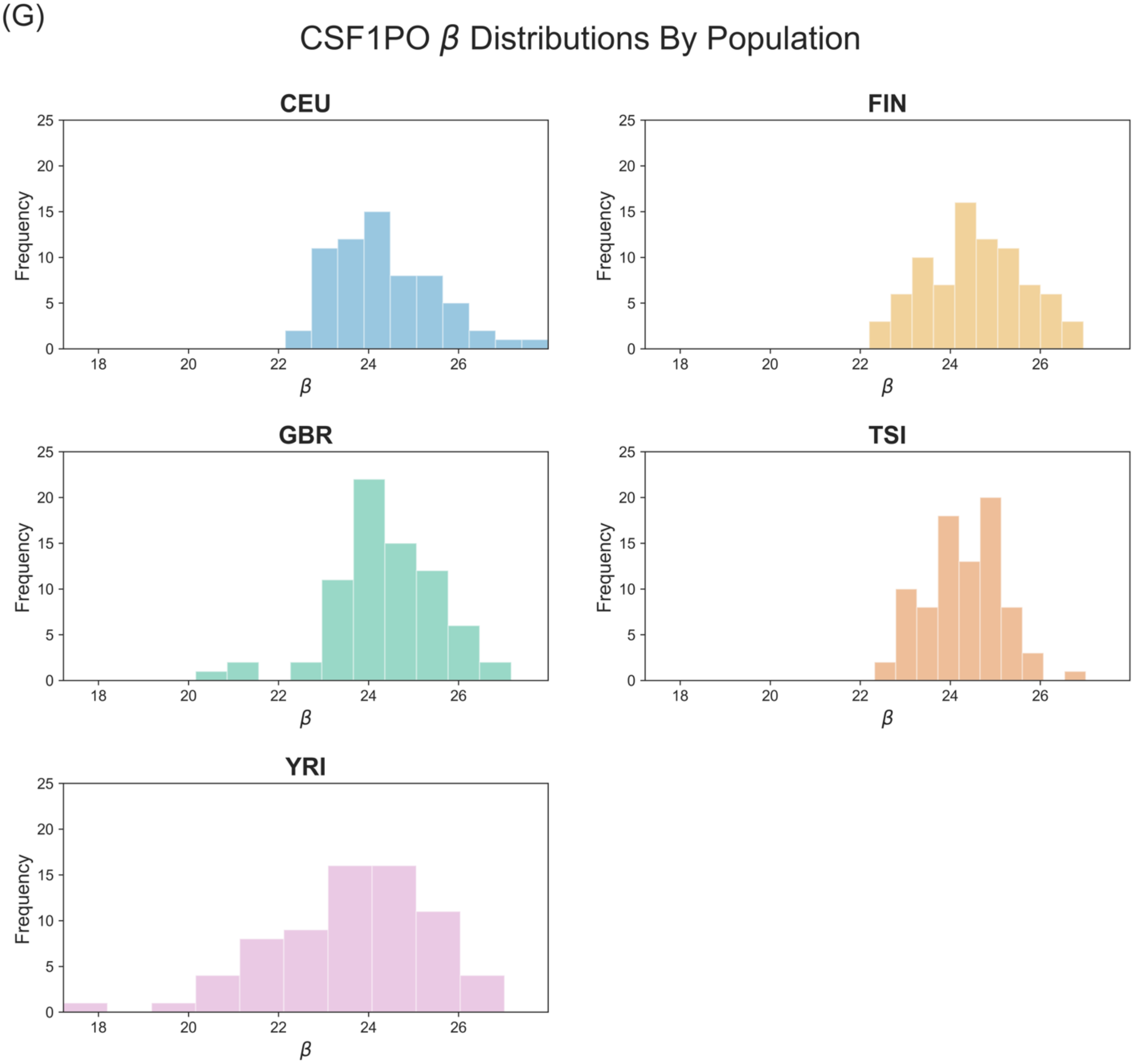

CSF1PO β Distributions By Population
CEU
FIN
GBR
TSI
YRI
Frequency
β

(H) *CSF1R* Gene Expression Distributions By Population

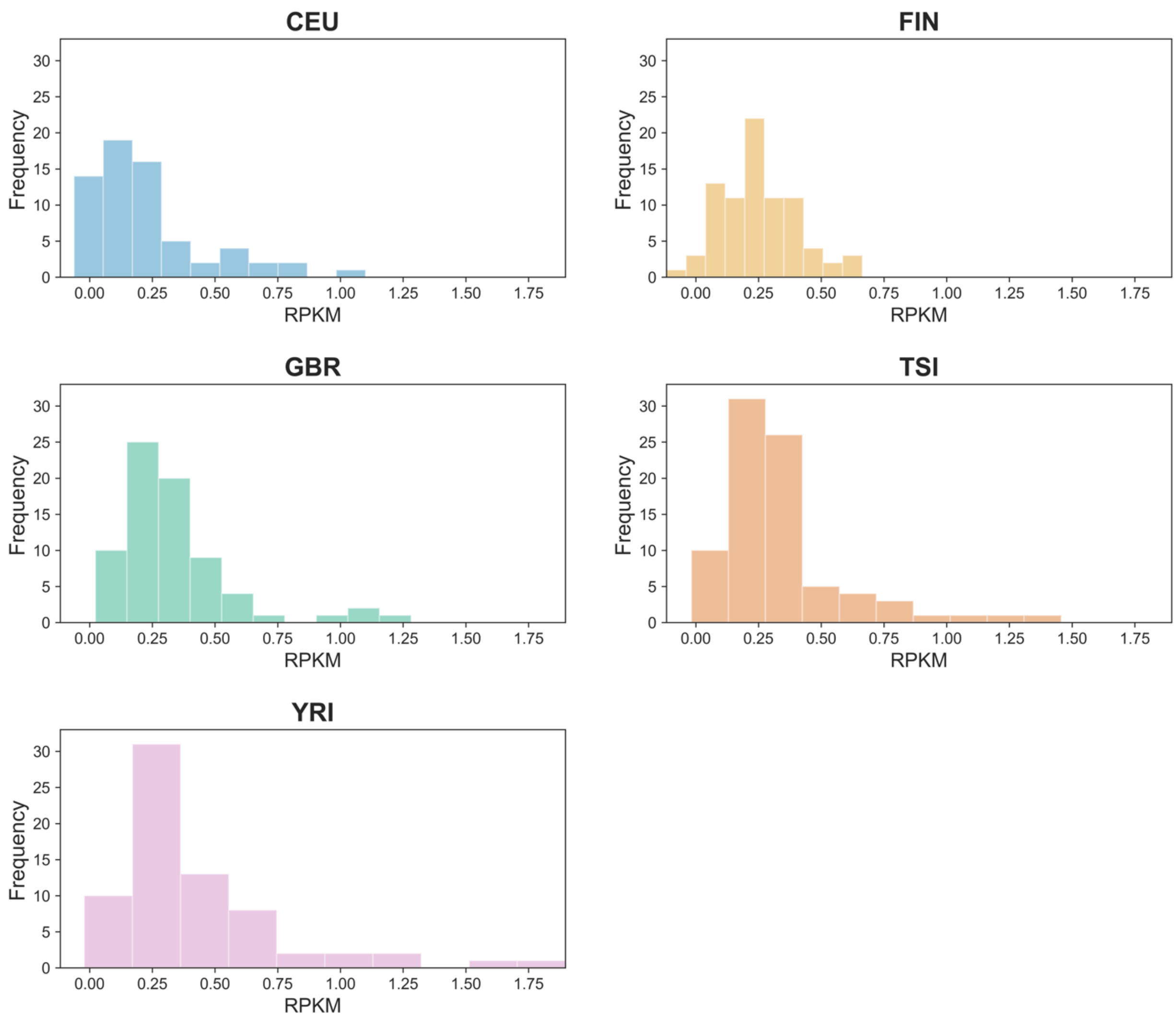

(I) *TIGD6* Gene Expression Distributions By Population

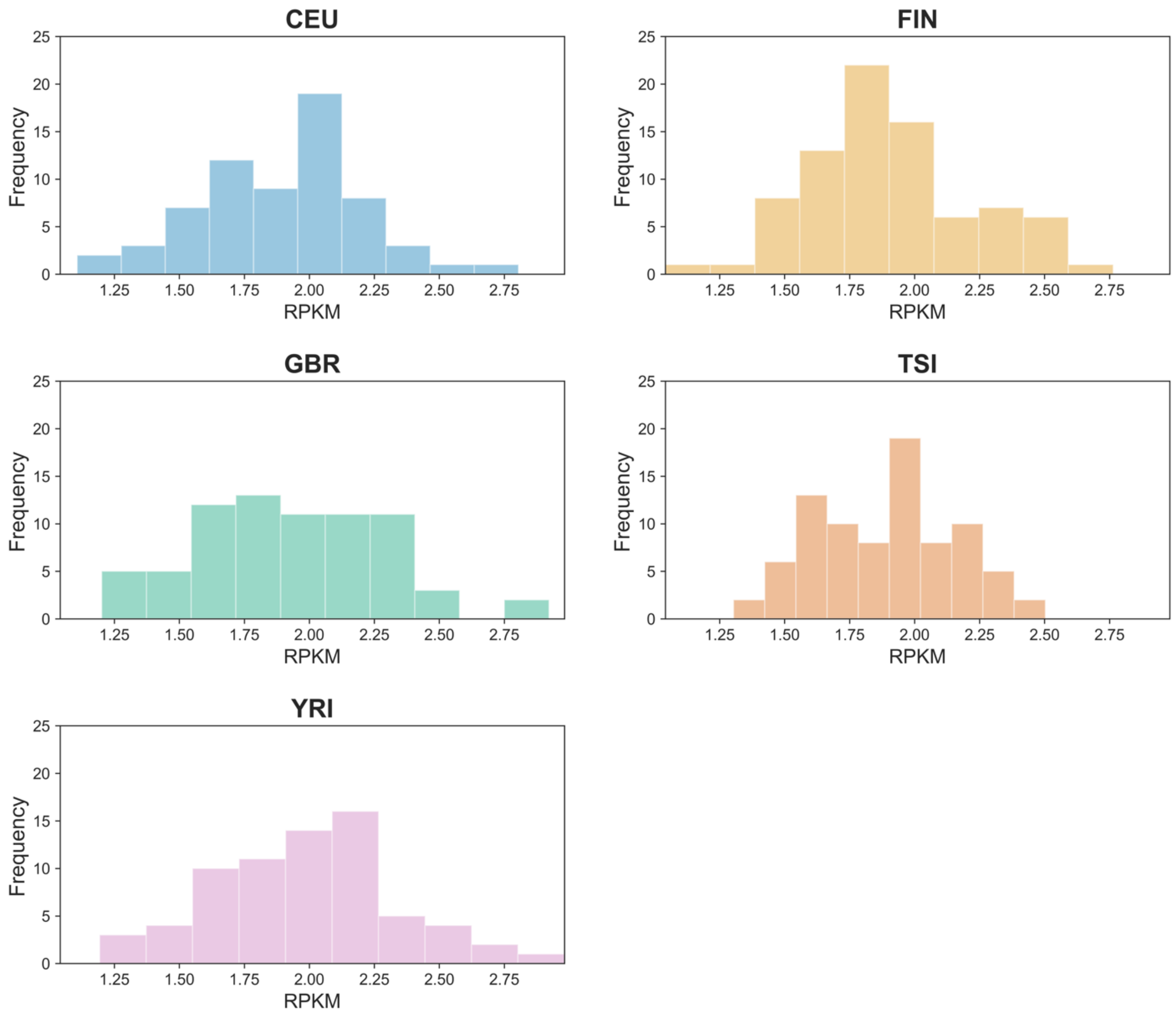

(J)

FGA $\beta$ Distributions By Population

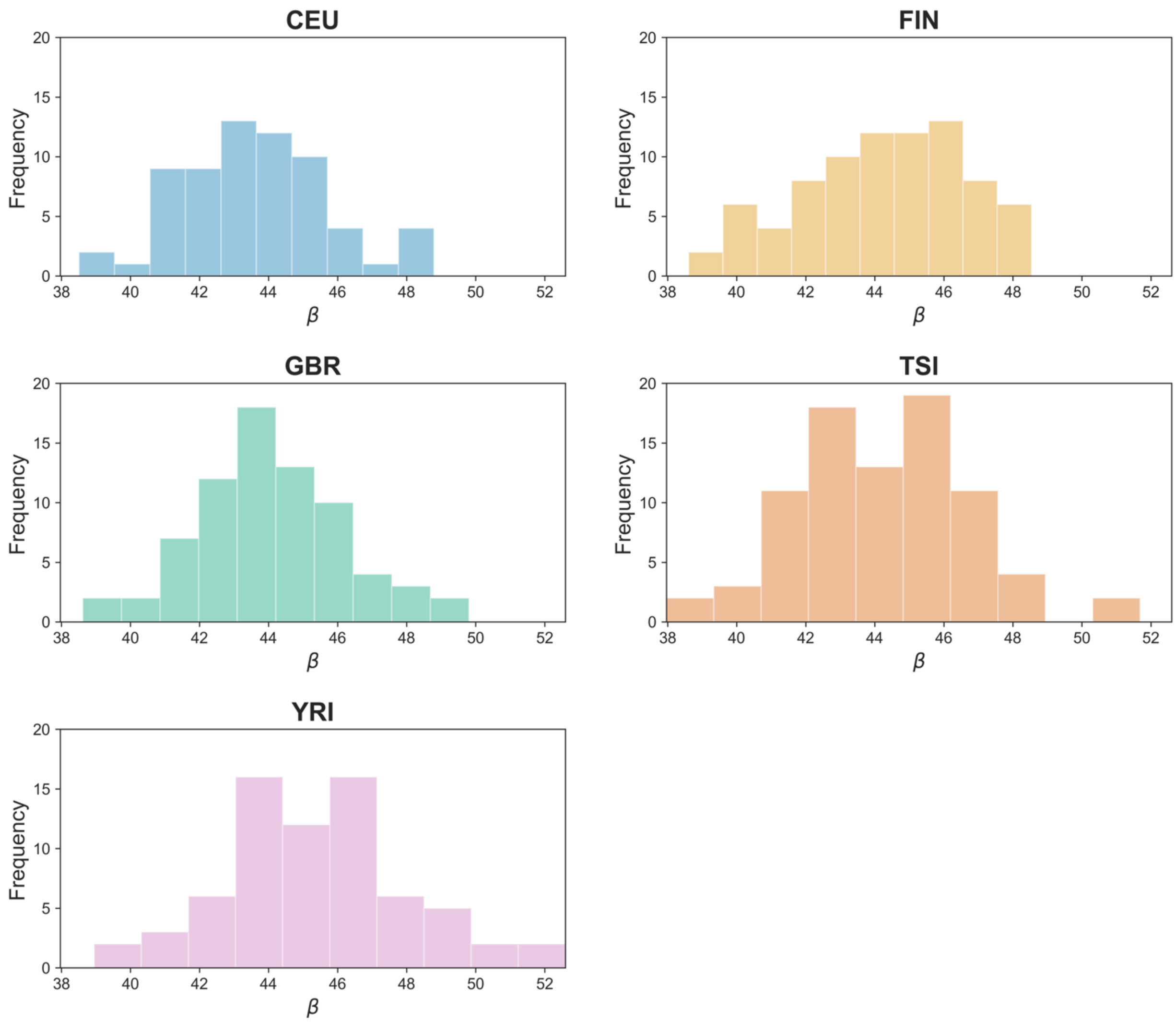

(K) *PLRG1* Gene Expression Distributions By Population

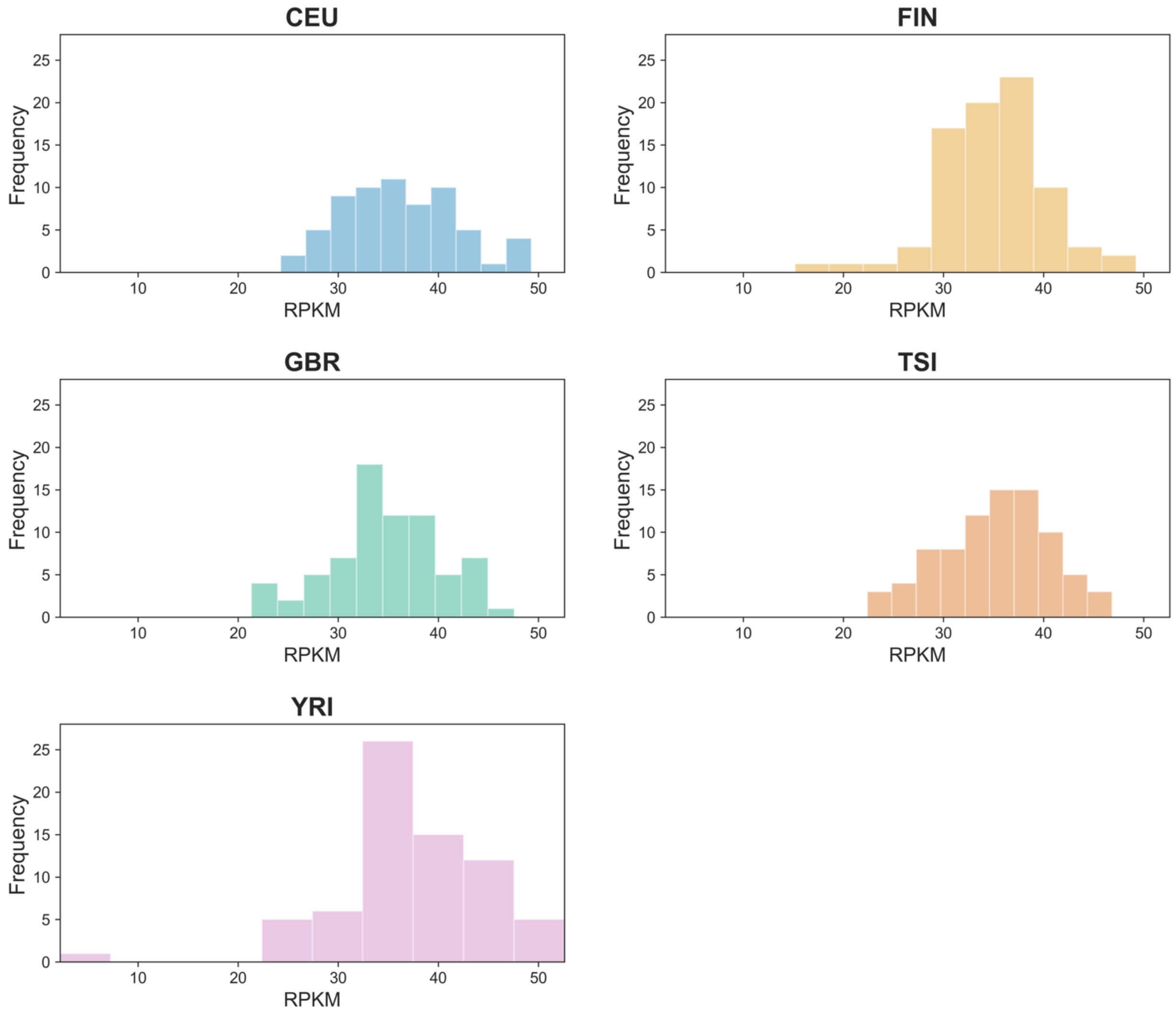

**Supplemental Figure 4: Genome-wide STR lengths.**
The histogram shows the lengths of genome wide STRs. The lengths of the CODISeSTRs are indicated with arrows as CSF1PO in orange, D18S51 in light blue, D2S441 in green, D3S1358 in yellow, and FGA in dark blue. Note that for purposes of the display, this plot excludes the 9 longest STRs from the genomic data, which have lengths above 15,000 base pairs.

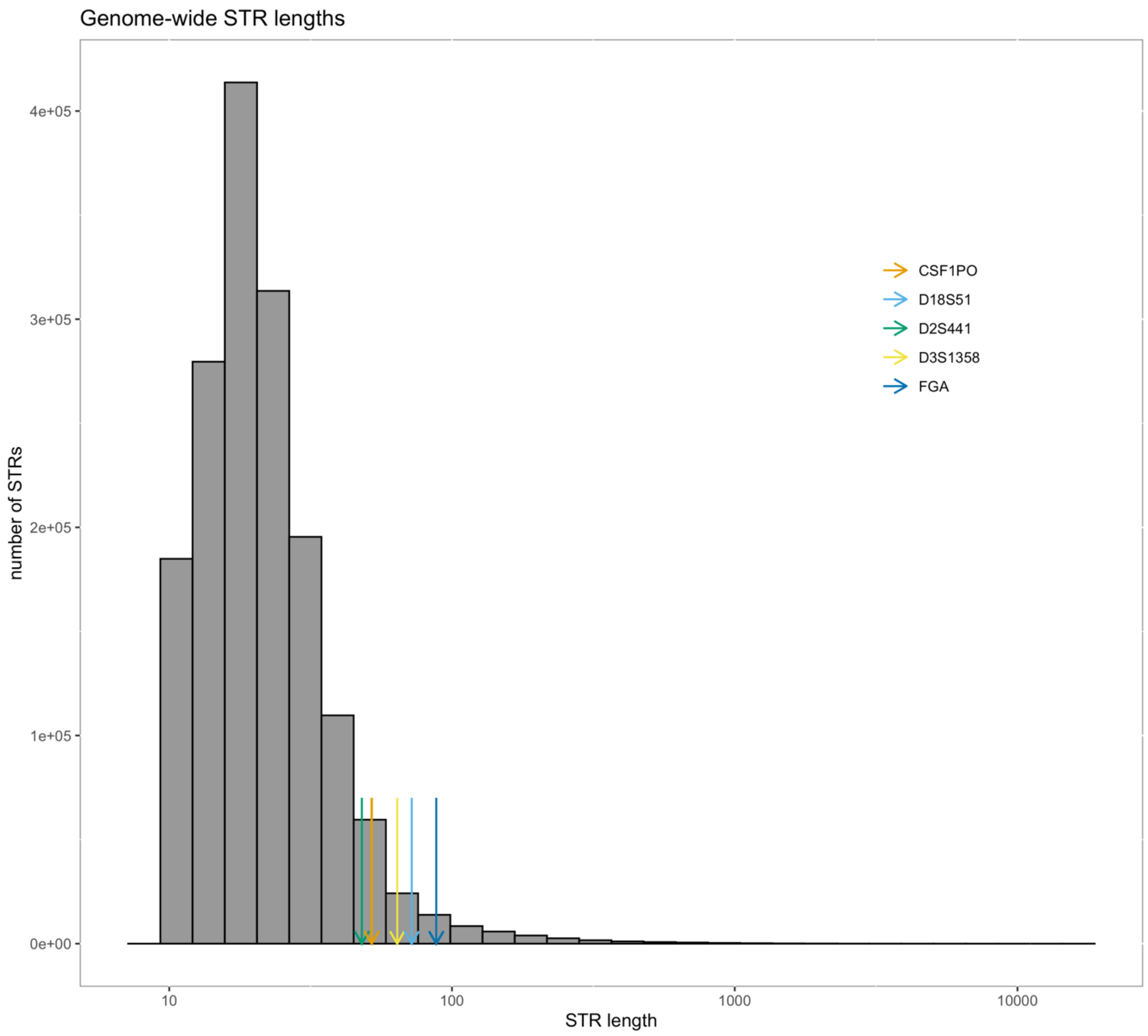

**Supplemental Figure 5: Genome-wide STR distances to nearest TSS**
The histogram shows distance between genome wide STRs and the nearest TSS. The respective distances of the CODISeSTRs are indicated with arrows: CSF1PO in orange, D18S51 in light blue, D2S441 in green, D3S1358 in yellow, and FGA in dark blue.

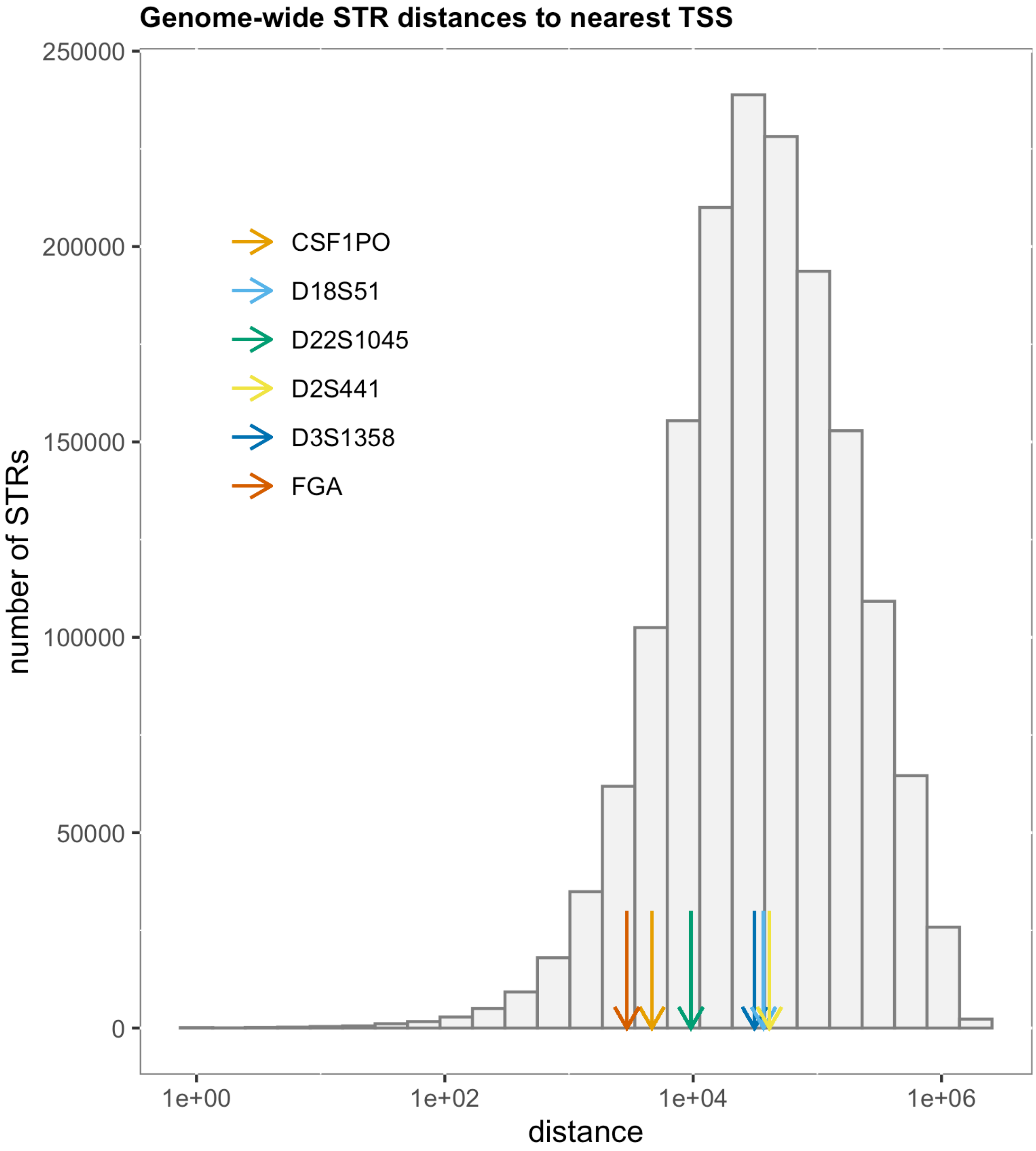

**Supplemental Figure 6: Genome-wide distances between STRs and DNAseIHS sites.**
The histogram shows the distance between each STR in the human genome and the nearest DNAseIHS site. The distances for the CODISeSTRs are indicated with arrows as CSF1PO in orange, D18S51 in light blue, D2S441 in green, D3S1358 in yellow, and FGA in dark blue.

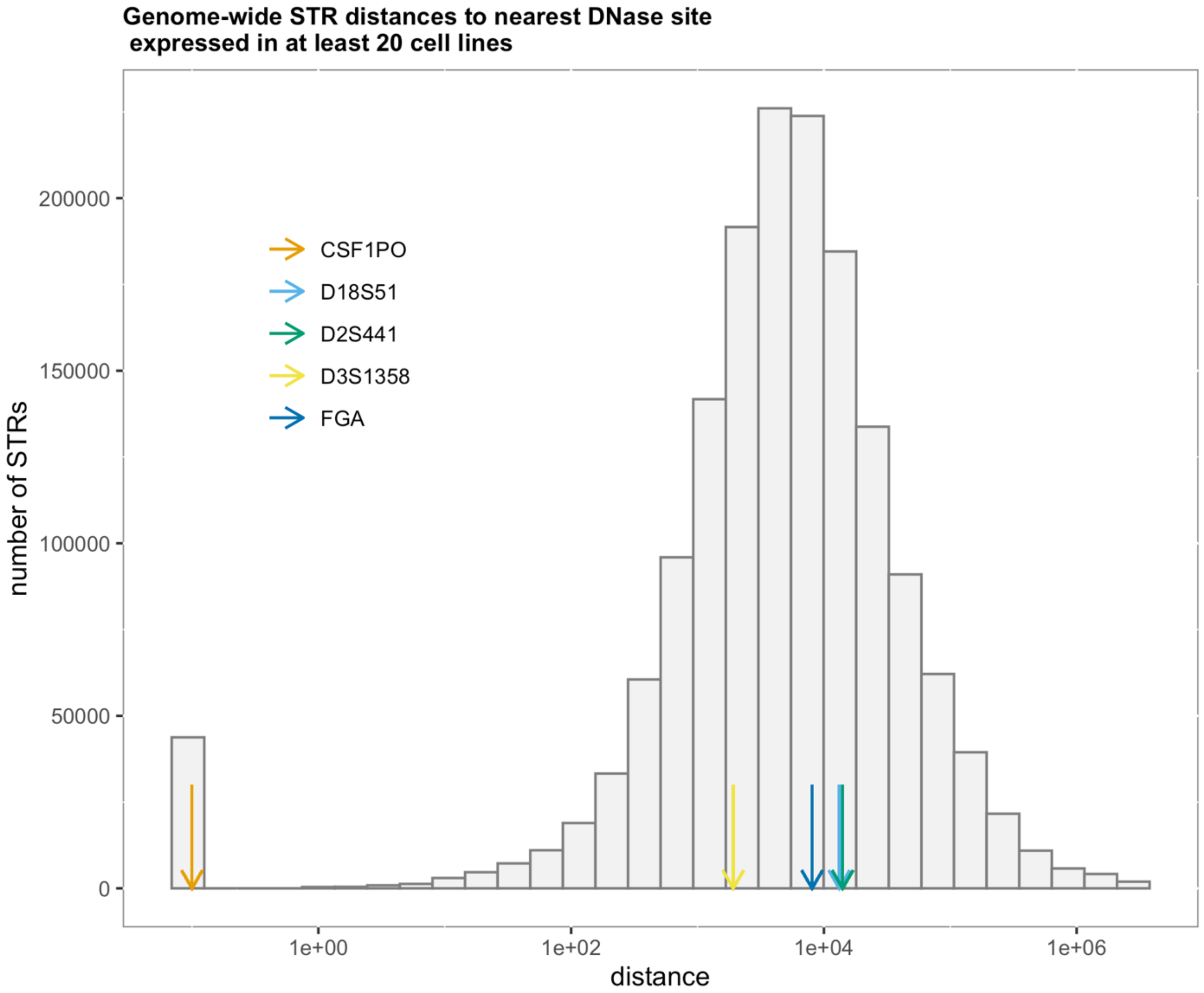

**Supplemental Figure 7: Genome-wide distances between STRs and DNAseIHS sites found in lymphoblastoid cell lines.**

The histogram shows the distance between each STR in the human genome and the nearest lymphoblasts or lymphoblast derivative DNAseIHS site. The distances for the CODISeSTRs are indicated with arrows as CSF1PO in orange, D18S51 in light blue, D2S441 in green, D3S1358 in yellow, and FGA in dark blue.

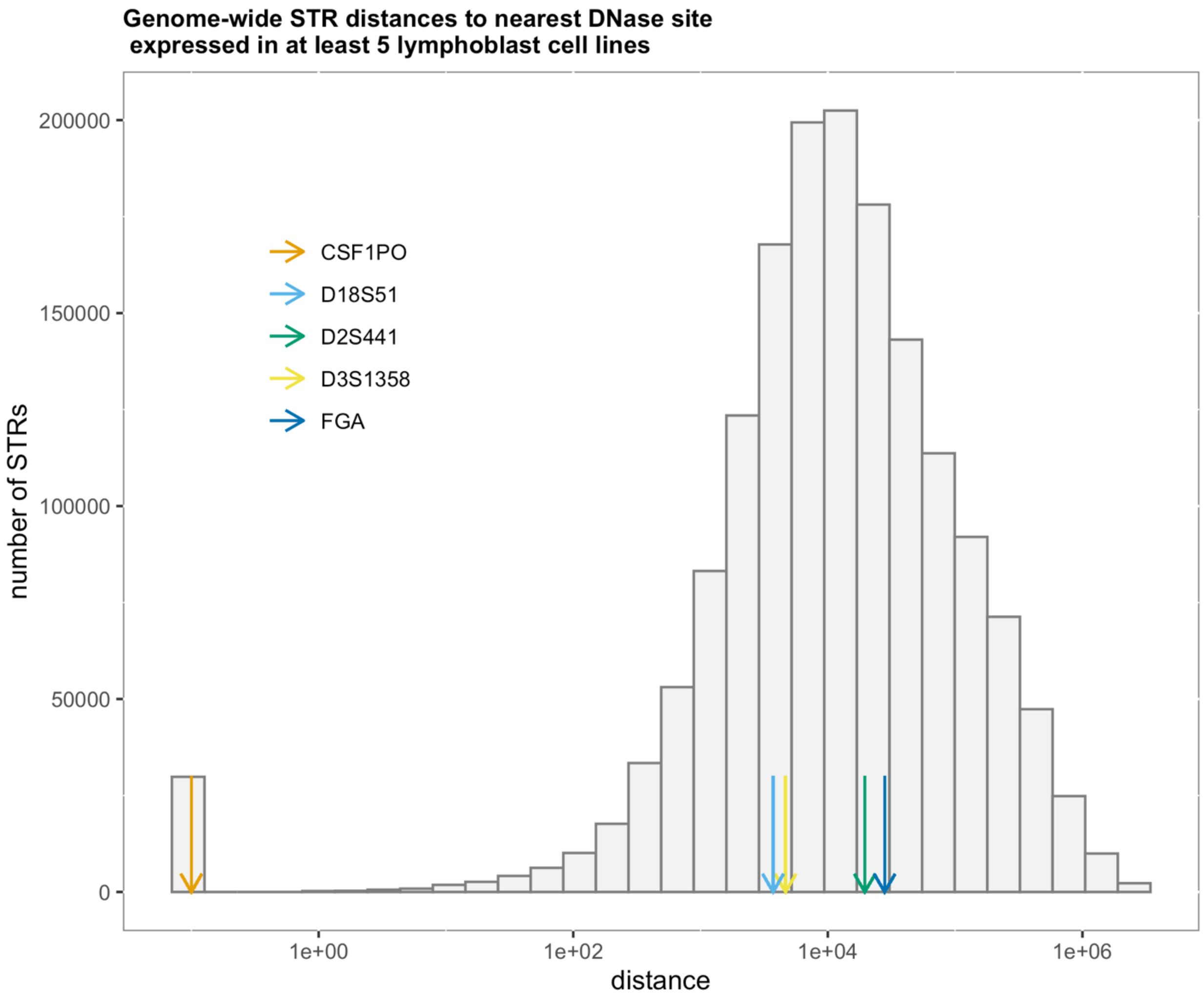

**Supplemental Table 5: CAVIAR Score Summaries.** Table containing CAVIAR results for putative causal variants within populations. We consider variants within a 100kb window centering on each gene of interest. Results are shown for *CSF1R* in the FIN and GBR populations, *KDSR* in the YRI population, and *LARS2* for FIN, GBR and TSI populations.

**Supplemental Figure 7: Local landscapes of LD and CAVIAR for KDSR and CSF1R .**

Each plot shows a 100kb window centered on the gene of interest. The top panels show LD between the CODISeSTR versus each variant in the ρ causal set. Bottom panels show CAVIAR scores for variants in the ρ causal set. Dark green circles enclose putative causal variants in both CAVIAR and LD panels. Vertical line indicates the location of the CODISeSTR. a) *KDSR* gene for YRI subpopulation, b) *CSF1R* gene in the FIN subpopulation, and c) *CSF1R* gene in the GBR population.

a)

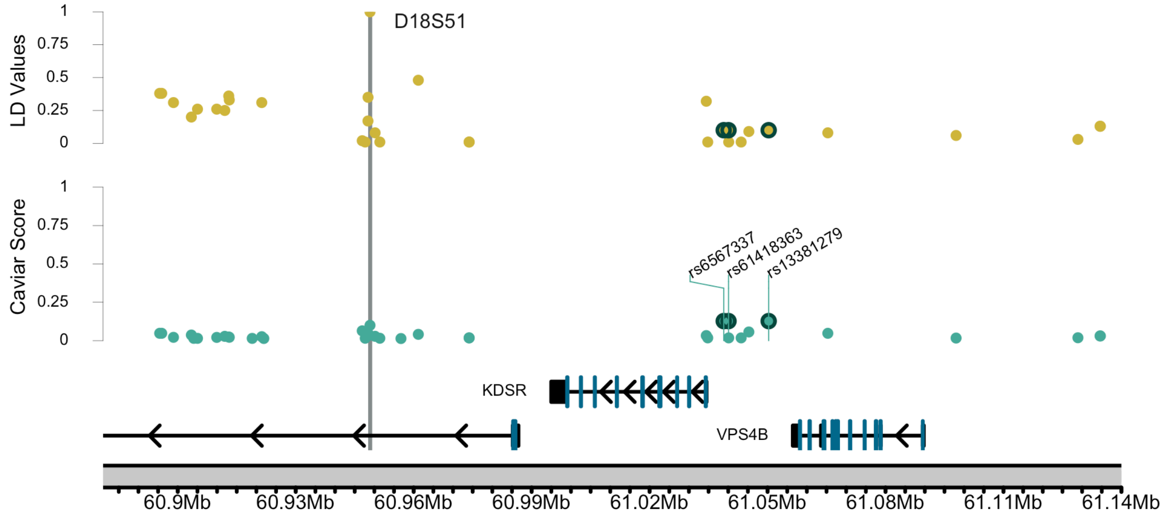

b)

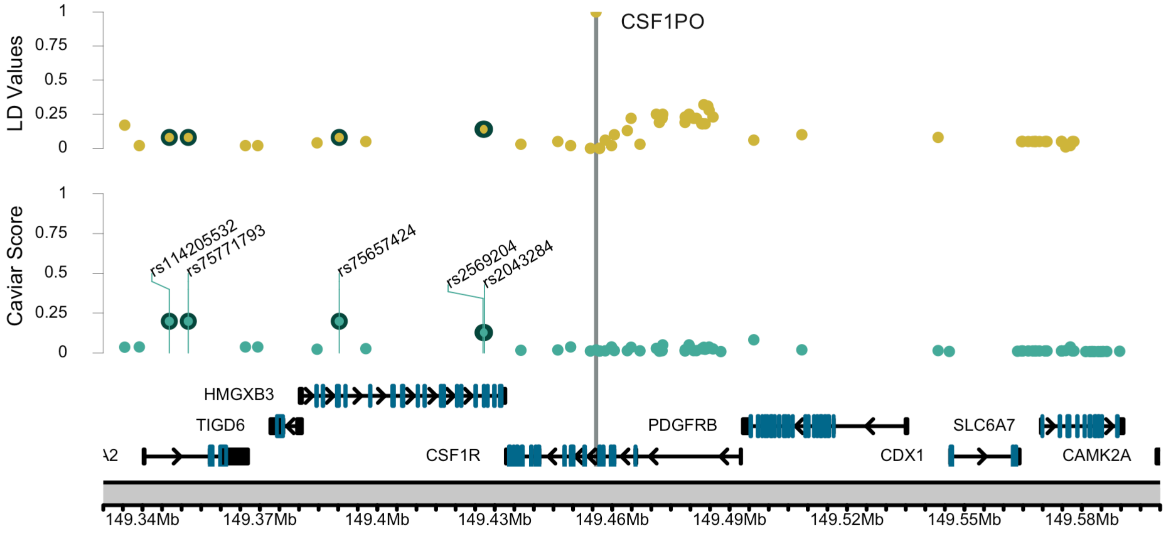

c)

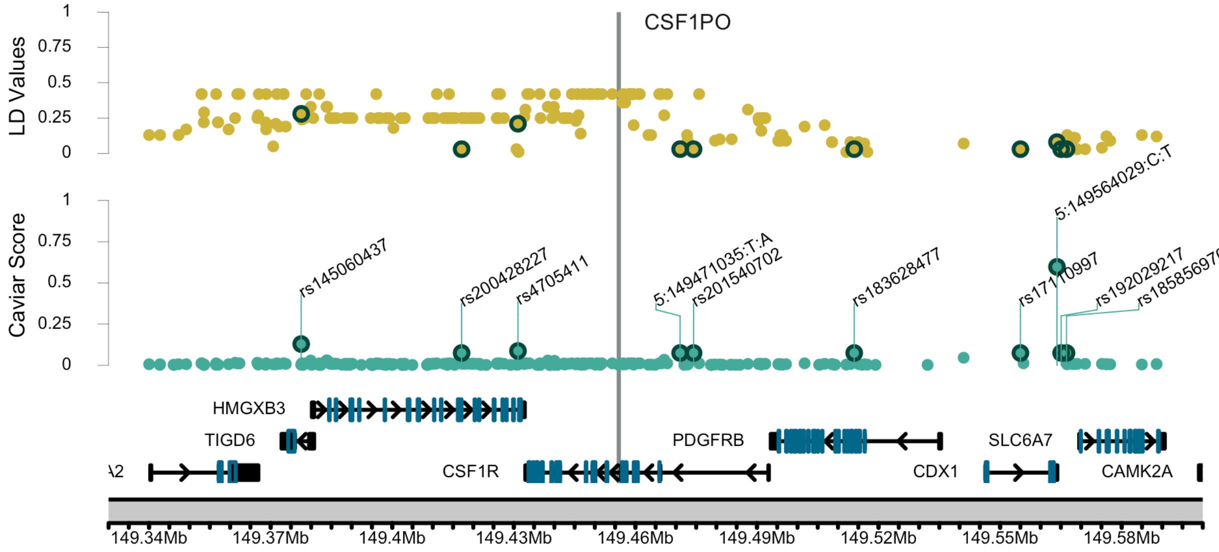

LD Values
CSF1PO
Caviar Score
rs145060437
rs200428227
rs4705411
5:149471035:T:A
rs201540702
rs183628477
5:149564029:C:T
rs17110997
rs192029217
rs185856970
HMGXB3
TIGD6
PDGFRB
SLC6A7
CSF1R
CDX1
CAMK2A
149.34Mb
149.37Mb
149.4Mb
149.43Mb
149.46Mb
149.49Mb
149.52Mb
149.55Mb
149.58Mb

**Supplemental Figure 8: Local landscapes of LD and DNAseIHSs for CODIS STRs**

Each plot shows a 100kb window centered on CODIS loci. The top panel shows the distributions of the LD between CODIS loci and surrounding SNPs. The middle panel maps the location of each DNAseIHS along the genome in green. The bottom panel represents local genes and their corresponding introns (black), coding exons (blue), non-coding exons (black), and strand marks (red). The highest LDs between CODIS loci and a local DNAseIHS are highlighted in green while all other SNPs are represented in yellow. (A) represents DNAseIHSs found within at least five lymphoblast cell lines. (B) represents DNAseIHSs found within at least 20 cell line sources. The location of CODIS loci is represented by a solid vertical line in the center.

**A** Multi-track LD Plot with DNAseIHSs found within >= 5 lymphoblast cell lines

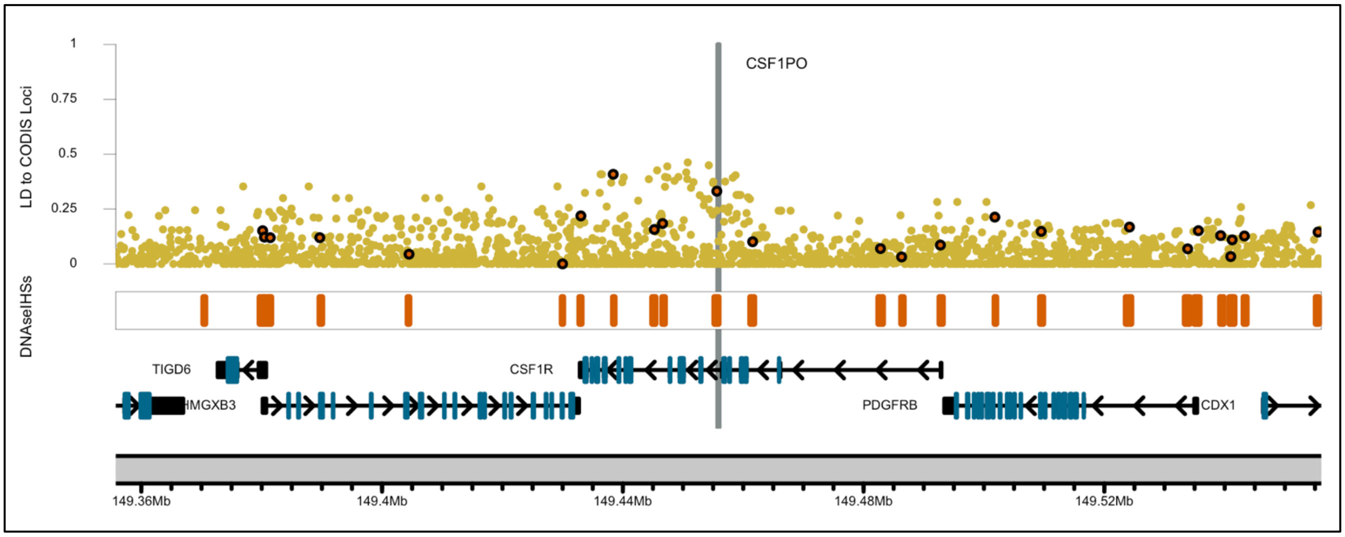

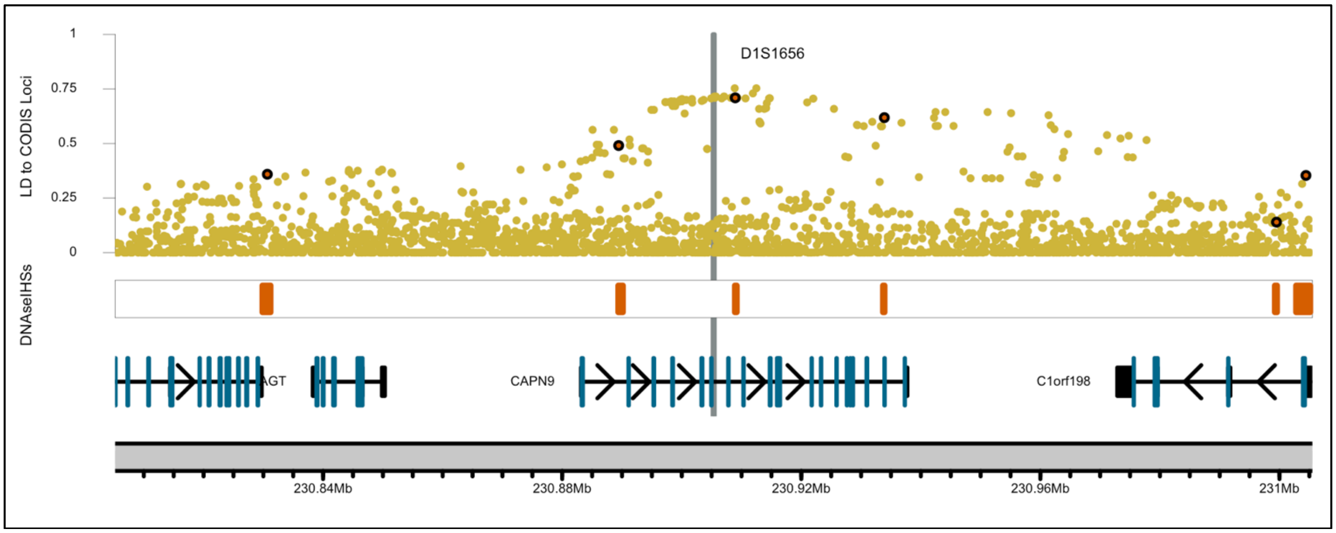

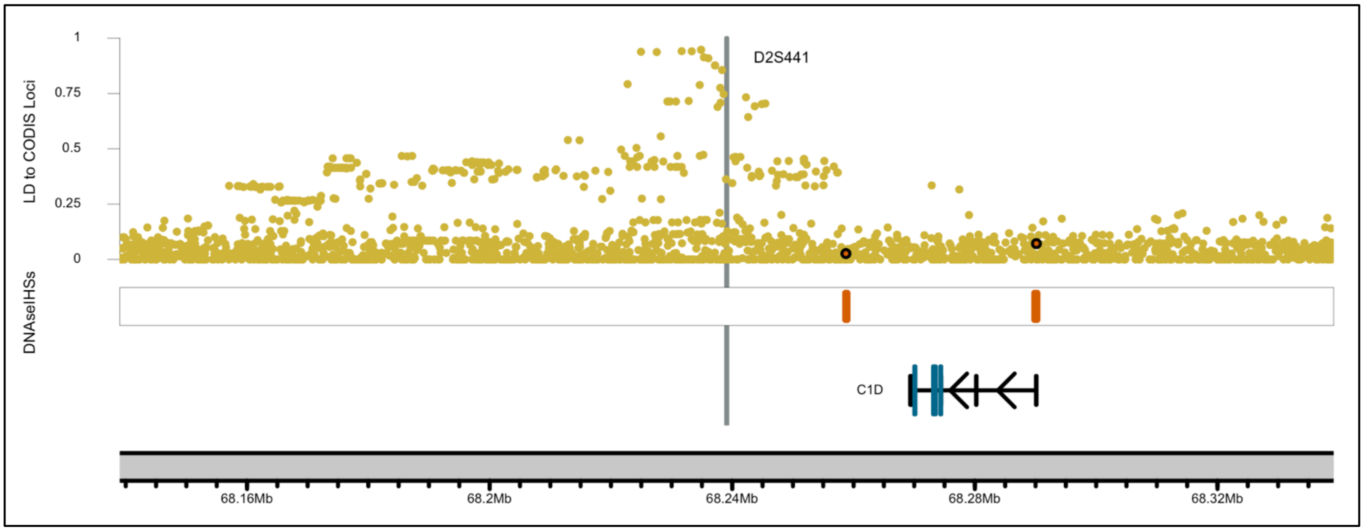
D2S441
LD to CODIS Loci
1
0.75
0.5
0.25
0
DNaseIHSs
C1D
68.16Mb
68.2Mb
68.24Mb
68.28Mb
68.32Mb

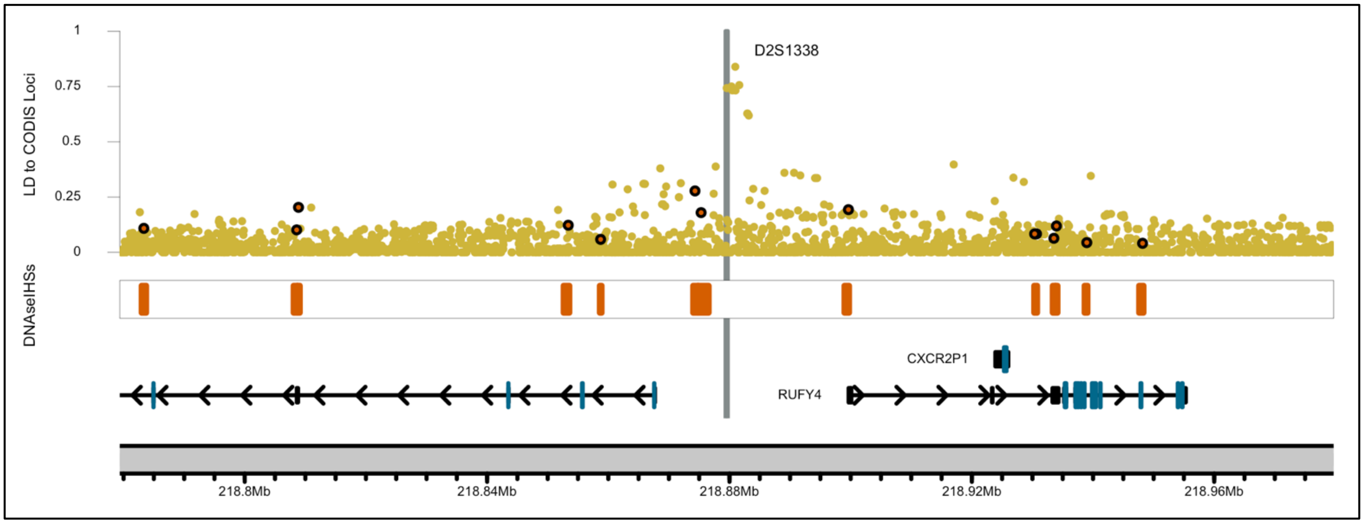
D2S1338
LD to CODIS Loci
1
0.75
0.5
0.25
0
DNaseIHSs
CXCR2P1
RUFY4
218.8Mb
218.84Mb
218.88Mb
218.92Mb
218.96Mb

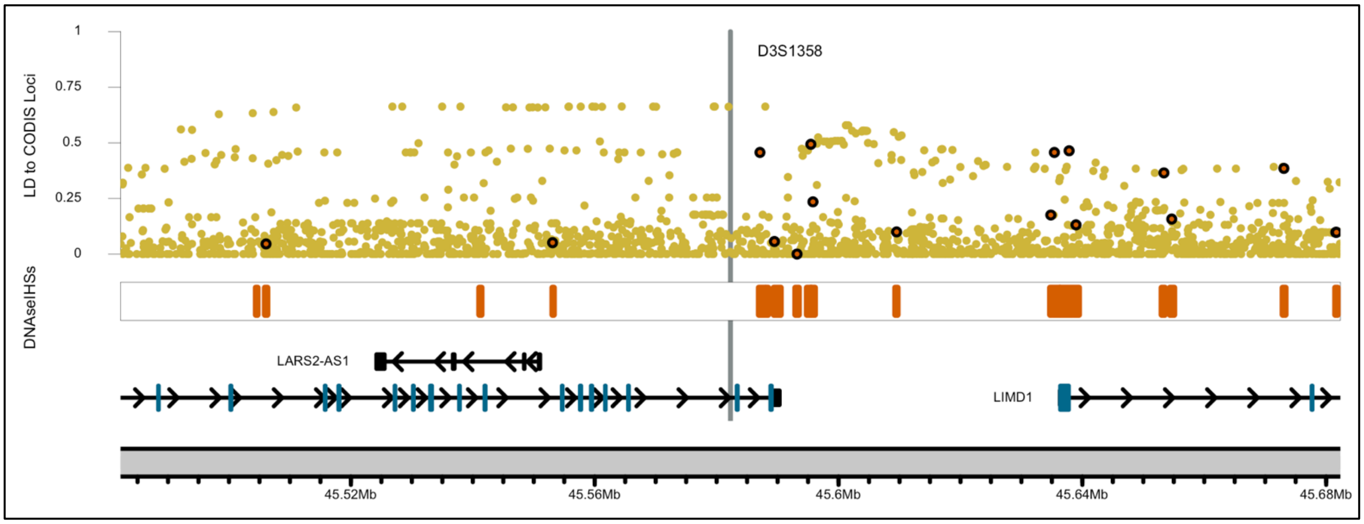
D3S1358
LD to CODIS Loci
1
0.75
0.5
0.25
0
DNaseIHSs
LARS2-AS1
LIMD1
45.52Mb
45.56Mb
45.6Mb
45.64Mb
45.68Mb

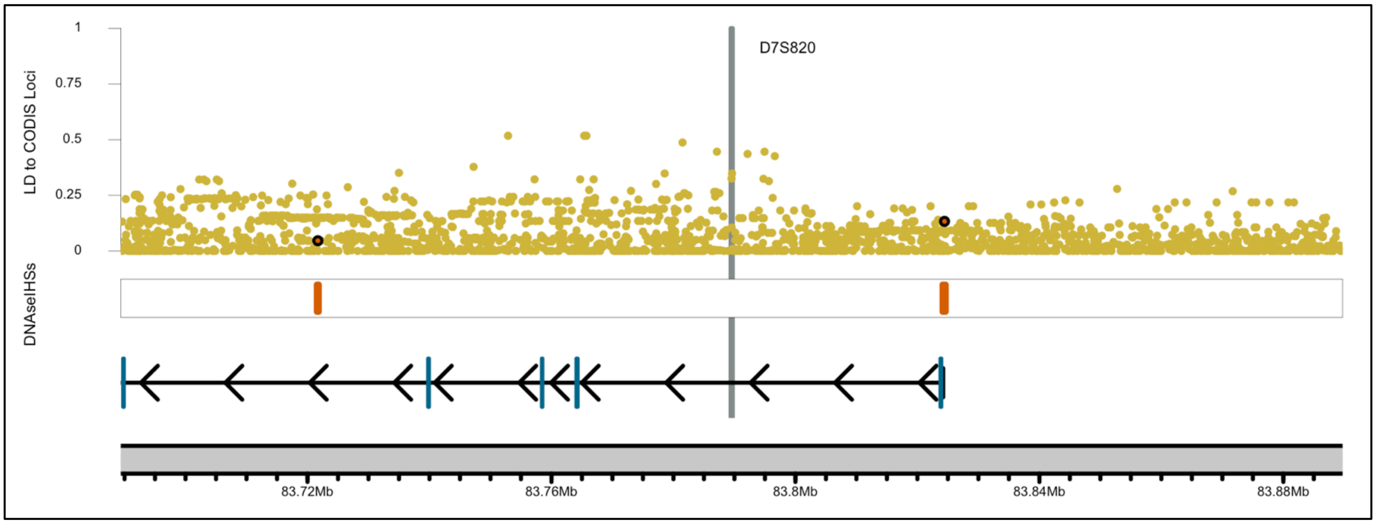
D7S820
LD to CODIS Loci
1
0.75
0.5
0.25
0
DNAseIHSs
83.72Mb
83.76Mb
83.8Mb
83.84Mb
83.88Mb

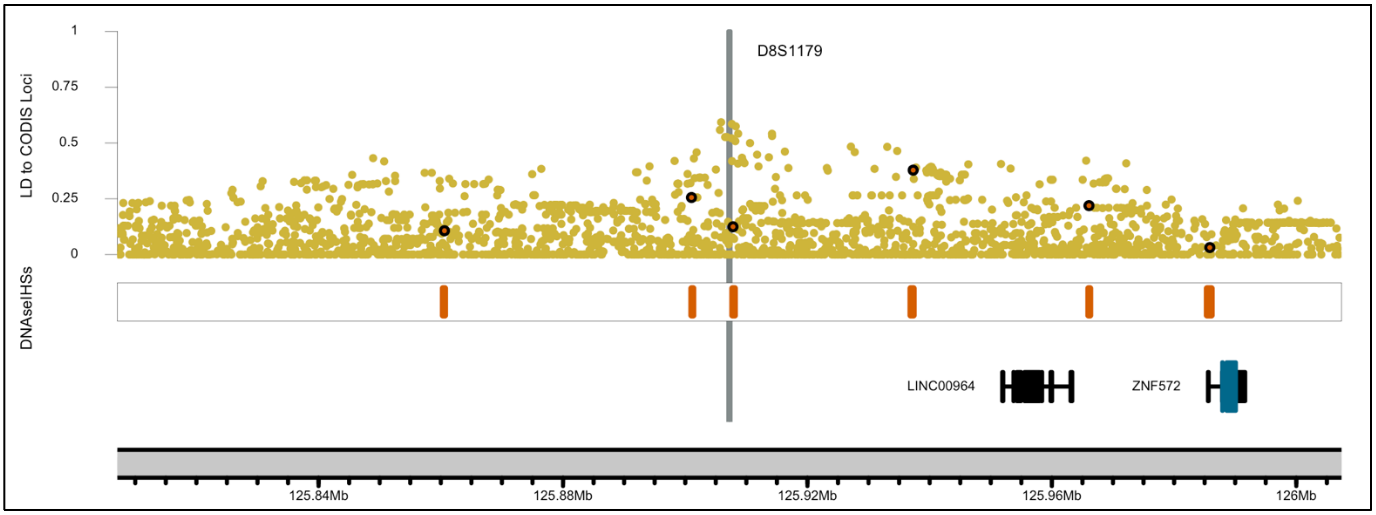
D8S1179
LD to CODIS Loci
1
0.75
0.5
0.25
0
DNAseIHSs
LINC00964
ZNF572
125.84Mb
125.88Mb
125.92Mb
125.96Mb
126Mb

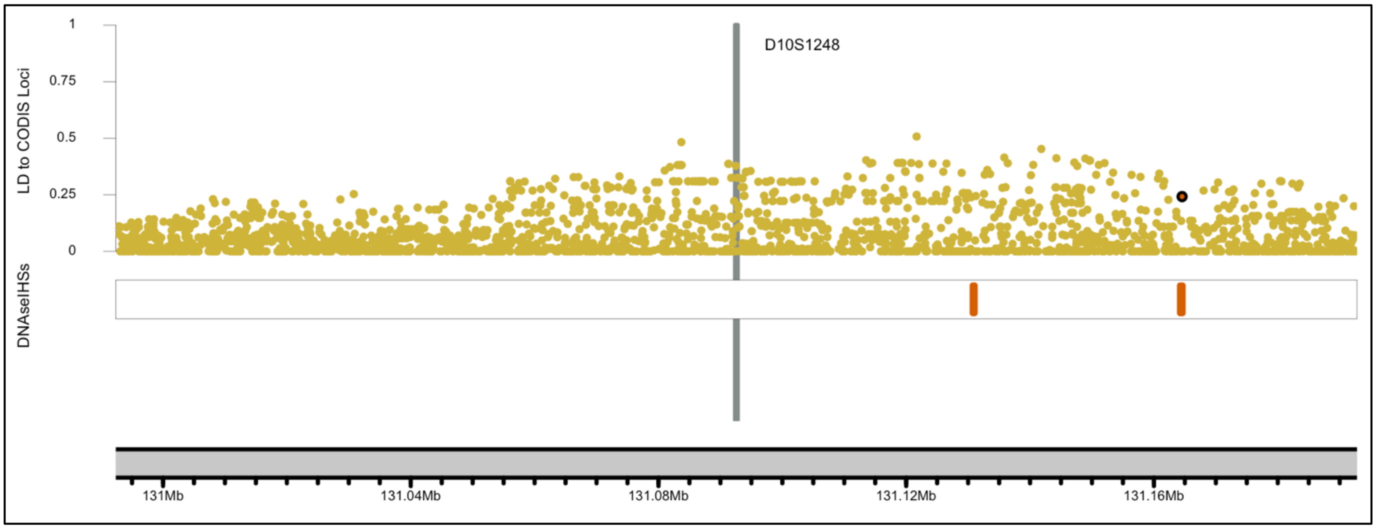
D10S1248
LD to CODIS Loci
1
0.75
0.5
0.25
0
DNAseIHSs
131Mb
131.04Mb
131.08Mb
131.12Mb
131.16Mb

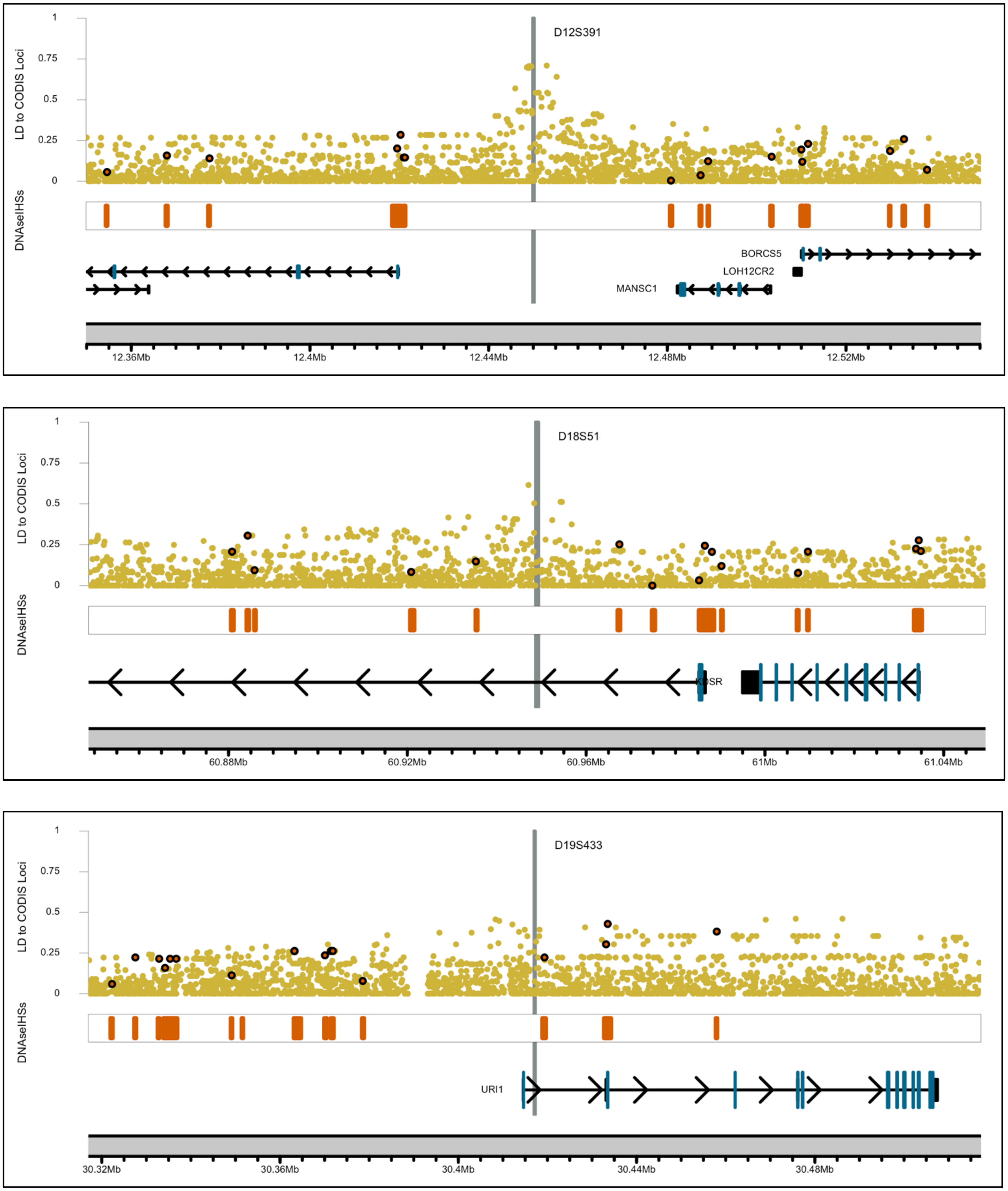
D12S391
LD to CODIS Loci
DNAseIHSs
1
0.75
0.5
0.25
0
BORCS5
LOH12CR2
MANSC1
12.36Mb
12.4Mb
12.44Mb
12.48Mb
12.52Mb
D18S51
60.88Mb
60.92Mb
60.96Mb
61Mb
61.04Mb
D19S433
URI1
30.32Mb
30.36Mb
30.4Mb
30.44Mb
30.48Mb

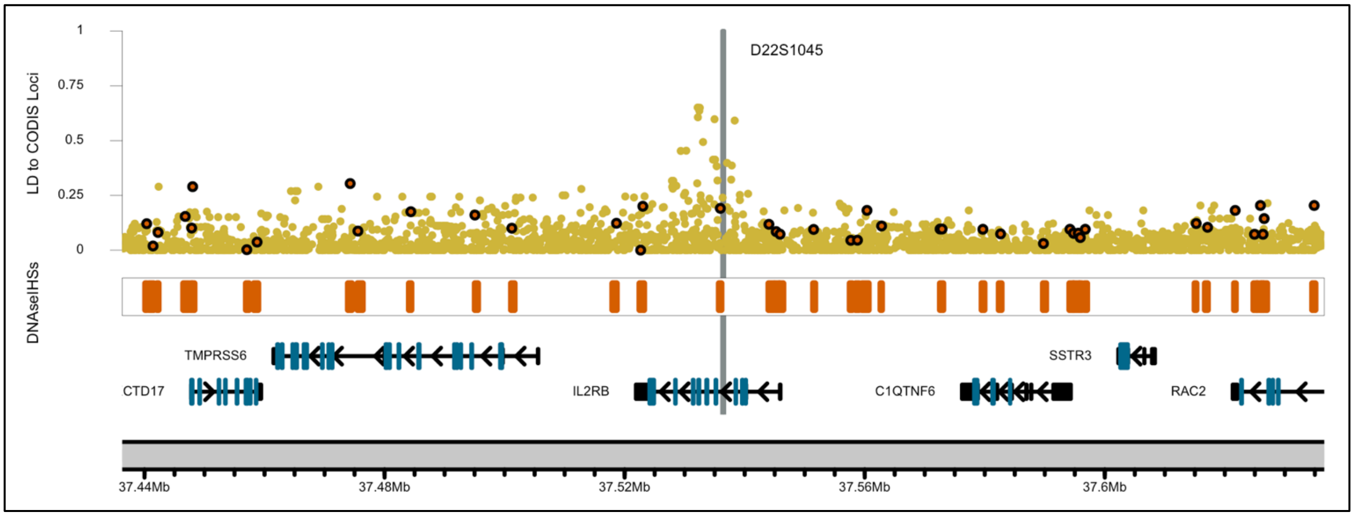
D22S1045
LD to CODIS Loci
DNAseIHSs
TMPRSS6
CTD17
IL2RB
C1QTNF6
SSTR3
RAC2
37.44Mb
37.48Mb
37.52Mb
37.56Mb
37.6Mb

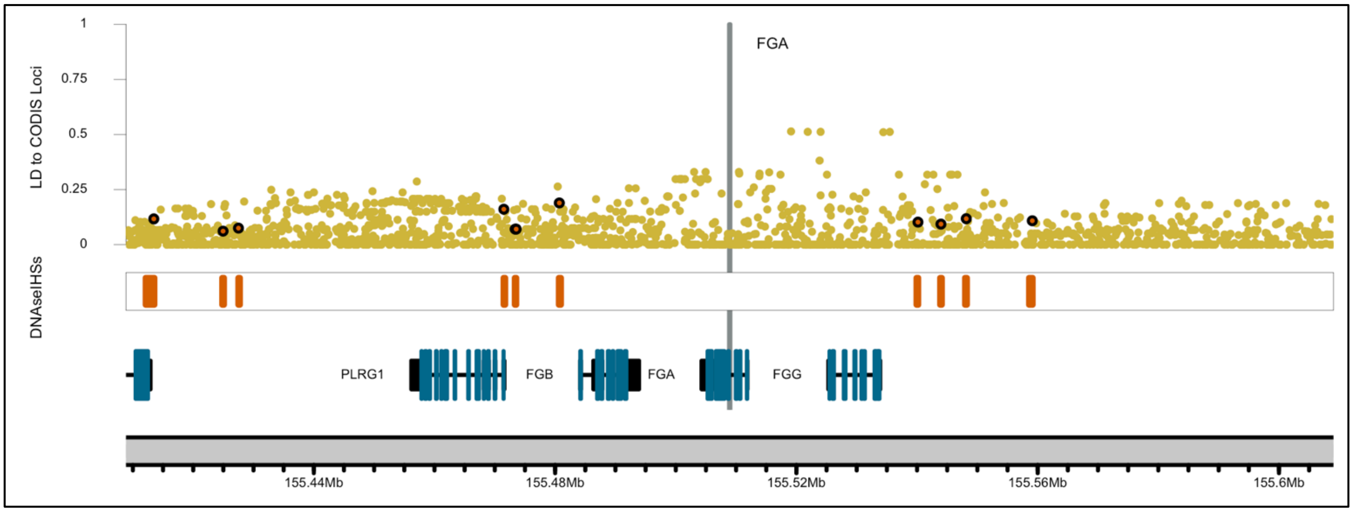
FGA
LD to CODIS Loci
DNAseIHSs
PLRG1
FGB
FGA
FGG
155.44Mb
155.48Mb
155.52Mb
155.56Mb
155.6Mb

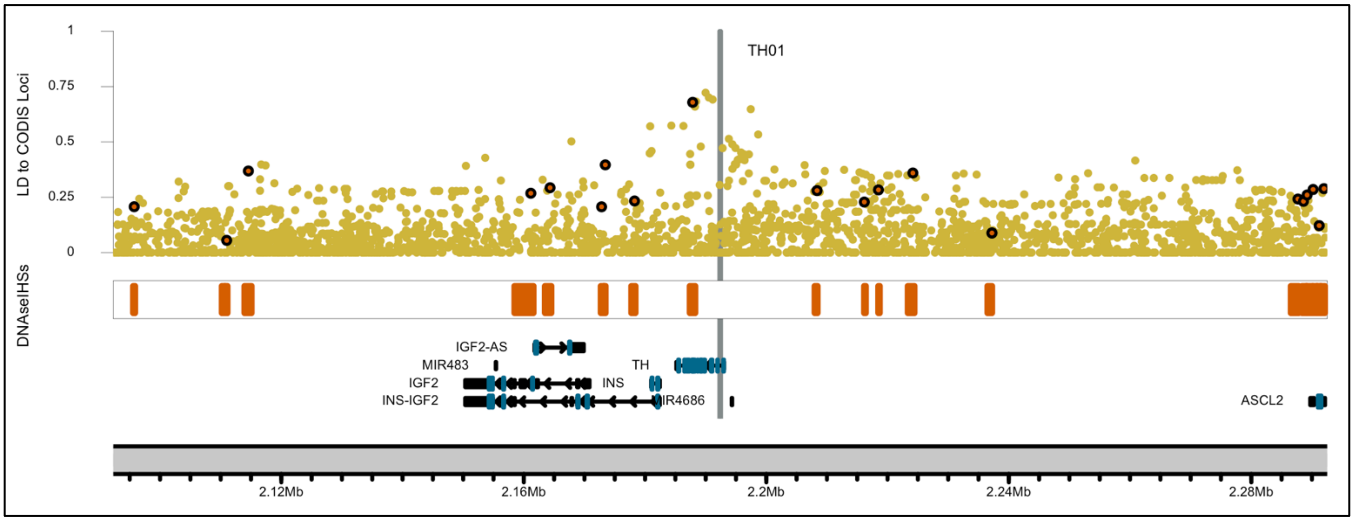
TH01
LD to CODIS Loci
DNAseIHSs
IGF2-AS
MIR483
IGF2
INS-IGF2
TH
INS
MIR4686
ASCL2
2.12Mb
2.16Mb
2.2Mb
2.24Mb
2.28Mb

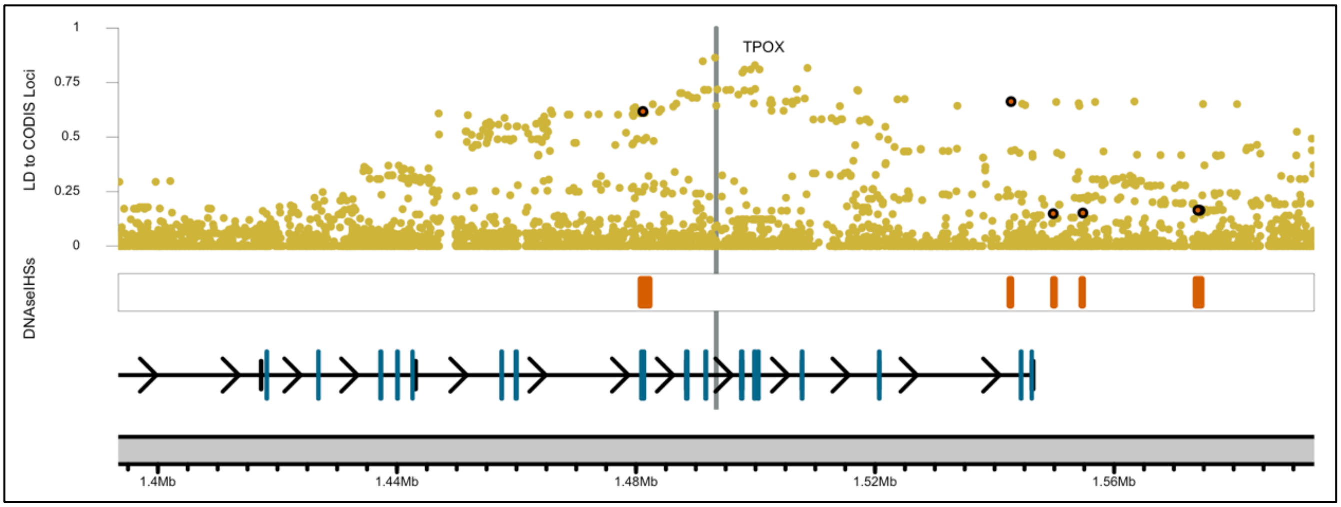

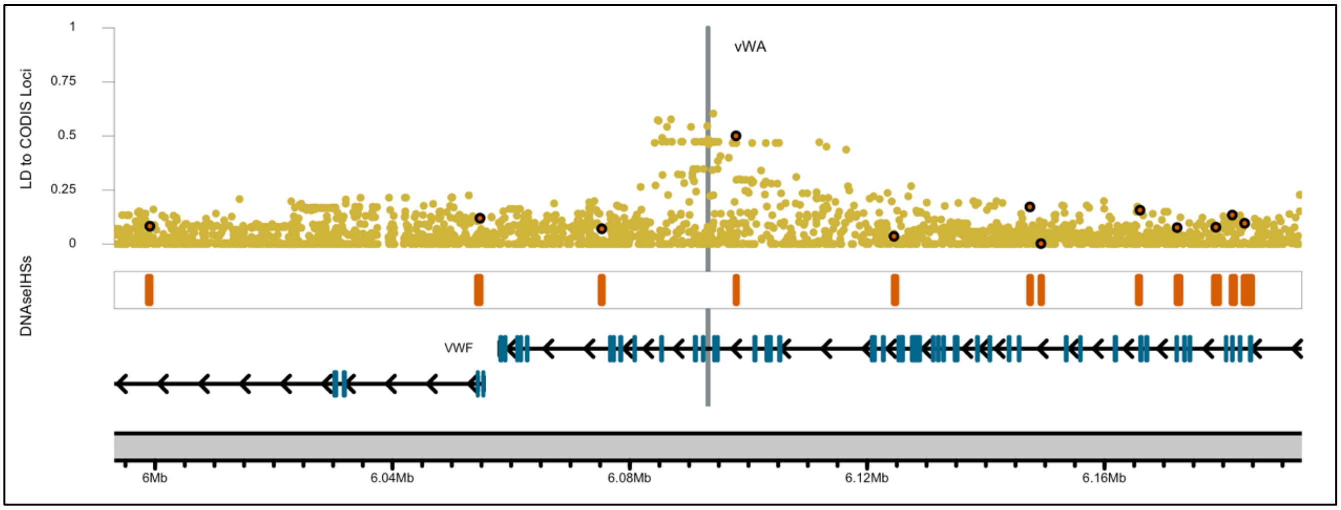

**B** Multi-track LD Plot with DNAseIHSs found within >= 20 cell line sources

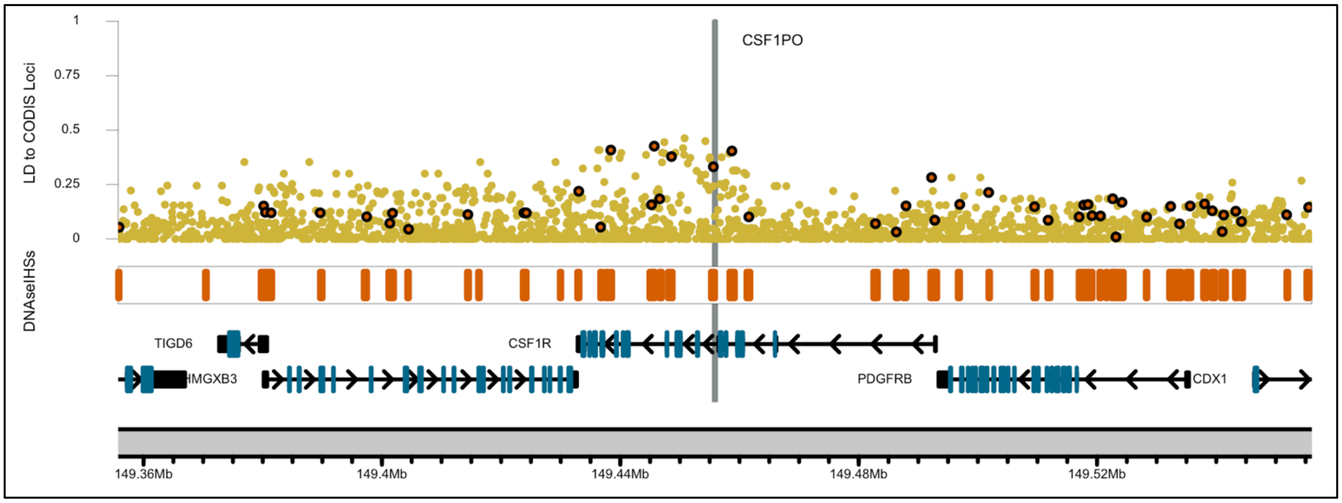

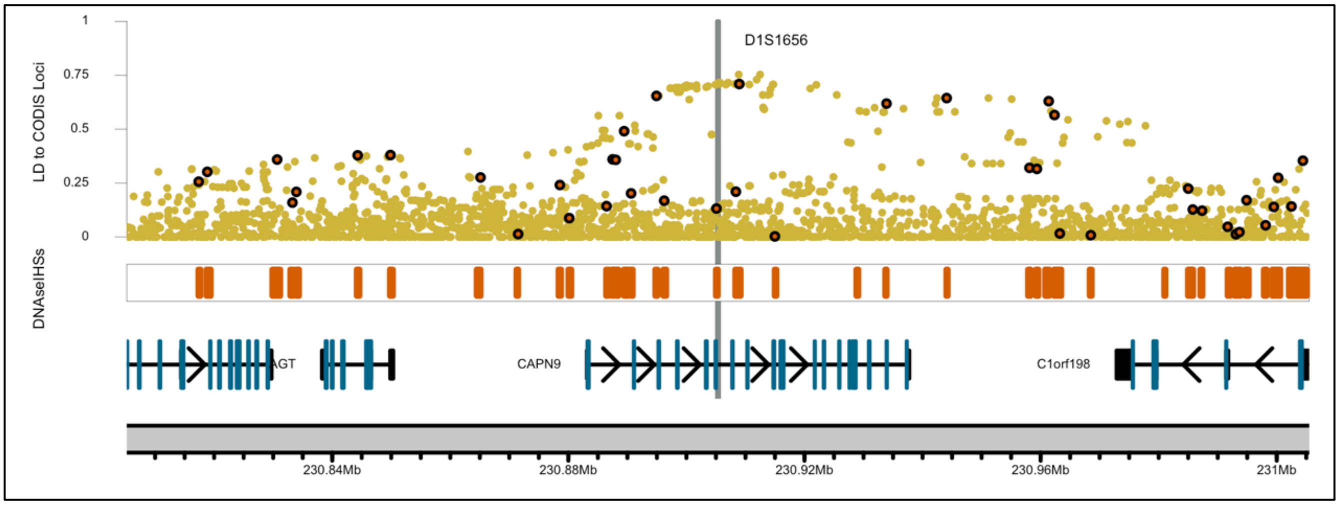
D1S1656
LD to CODIS Loci
1
0.75
0.5
0.25
0
DNAseIHSs
AGT
CAPN9
C1orf198
230.84Mb
230.88Mb
230.92Mb
230.96Mb
231Mb

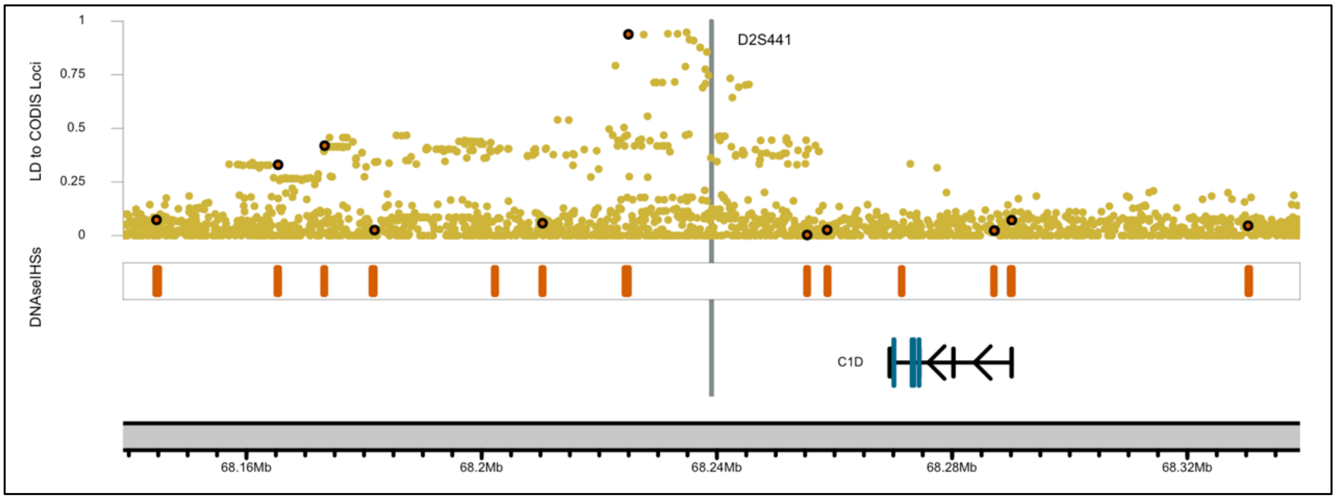
D2S441
LD to CODIS Loci
1
0.75
0.5
0.25
0
DNAseIHSs
C1D
68.16Mb
68.2Mb
68.24Mb
68.28Mb
68.32Mb

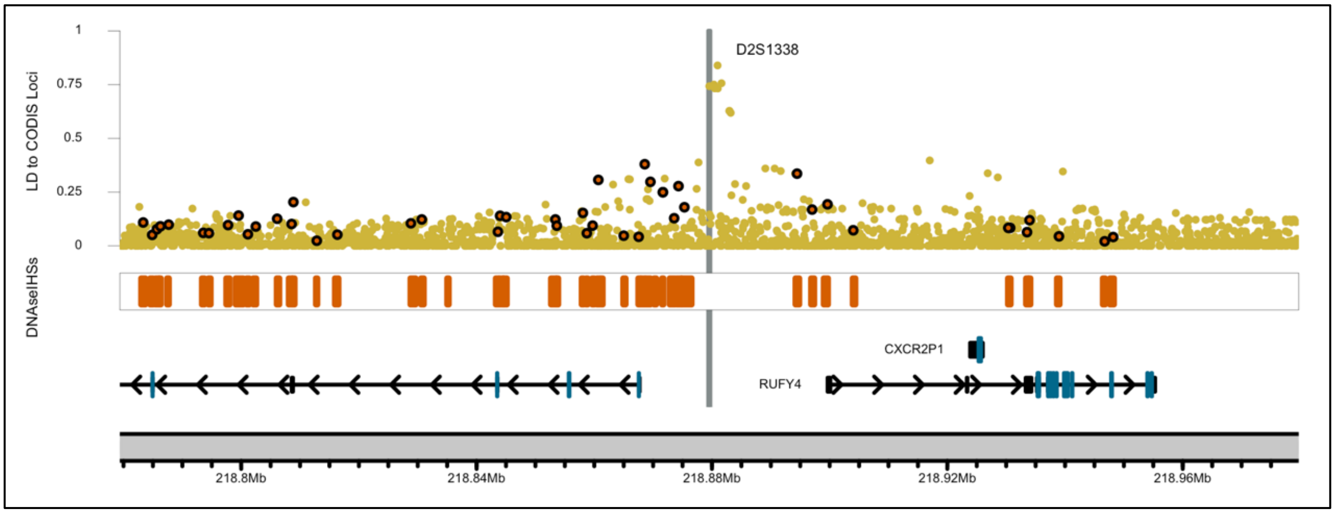
D2S1338
LD to CODIS Loci
1
0.75
0.5
0.25
0
DNAseIHSs
CXCR2P1
RUFY4
218.8Mb
218.84Mb
218.88Mb
218.92Mb
218.96Mb

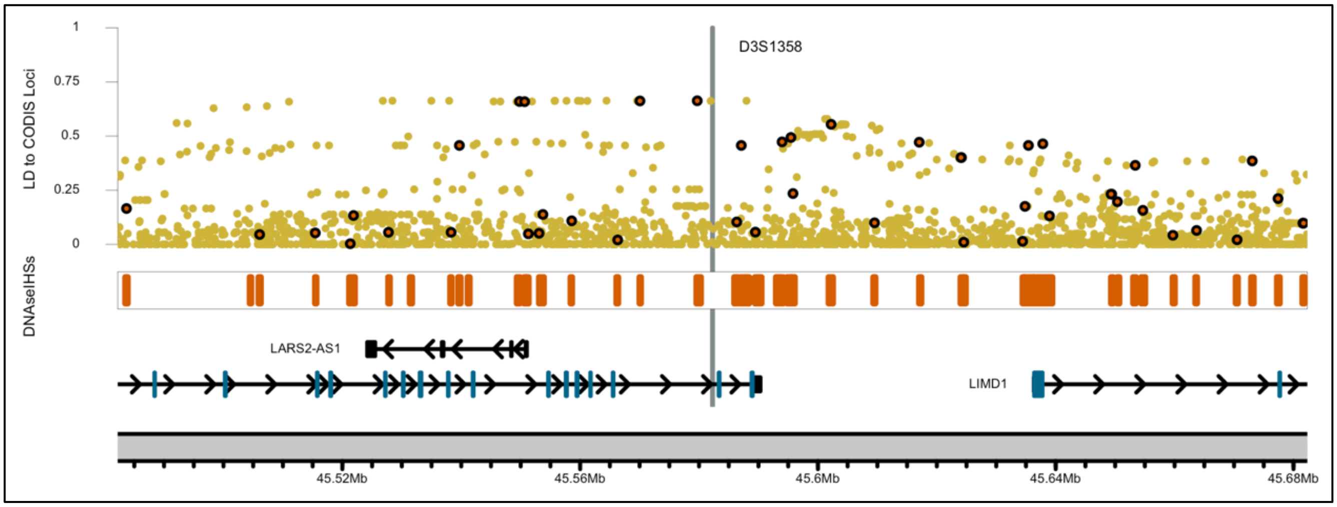
D3S1358
LD to CODIS Loci
1
0.75
0.5
0.25
0
DNAseIHSs
LARS2-AS1
LIMD1
45.52Mb
45.56Mb
45.6Mb
45.64Mb
45.68Mb

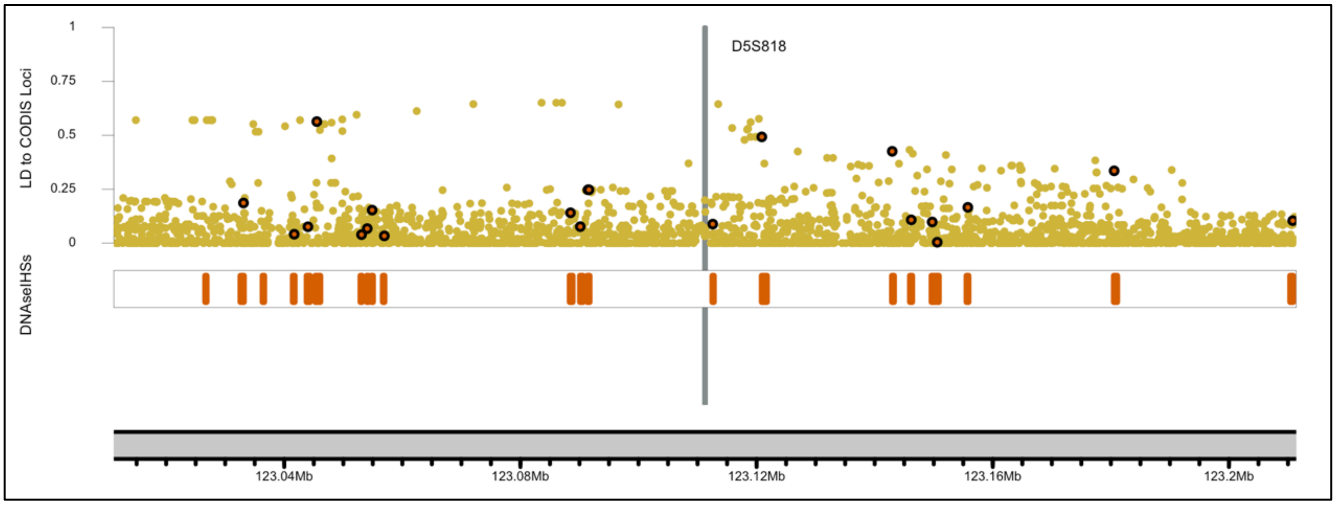
D5S818
LD to CODIS Loci
1
0.75
0.5
0.25
0
DNAseIHSs
123.04Mb
123.08Mb
123.12Mb
123.16Mb
123.2Mb

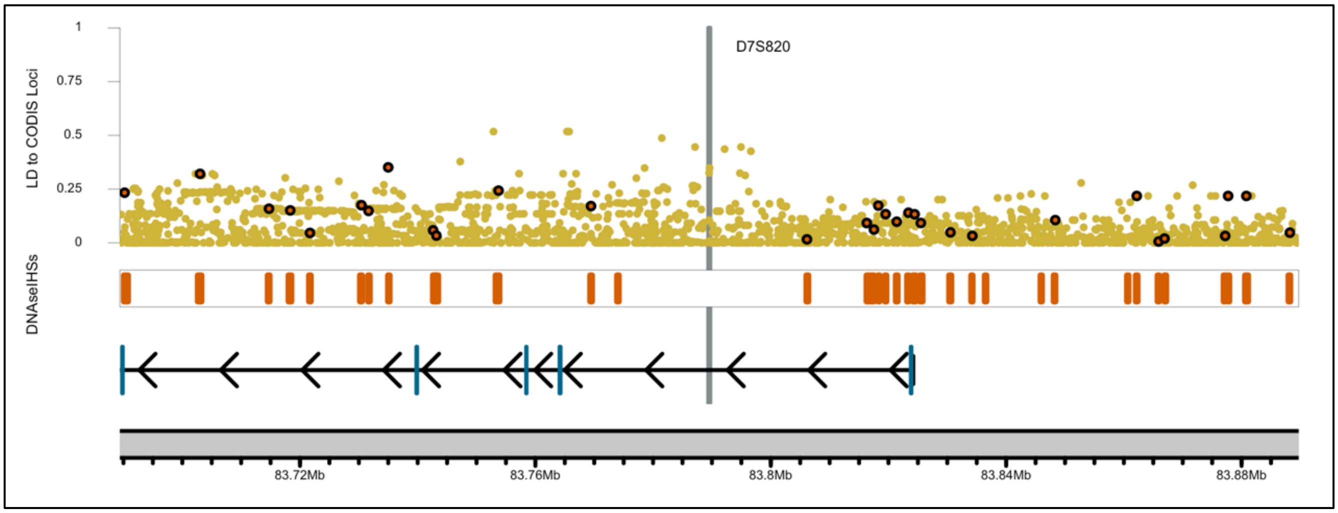
D7S820
LD to CODIS Loci
1
0.75
0.5
0.25
0
DNAseIHSs
83.72Mb
83.76Mb
83.8Mb
83.84Mb
83.88Mb

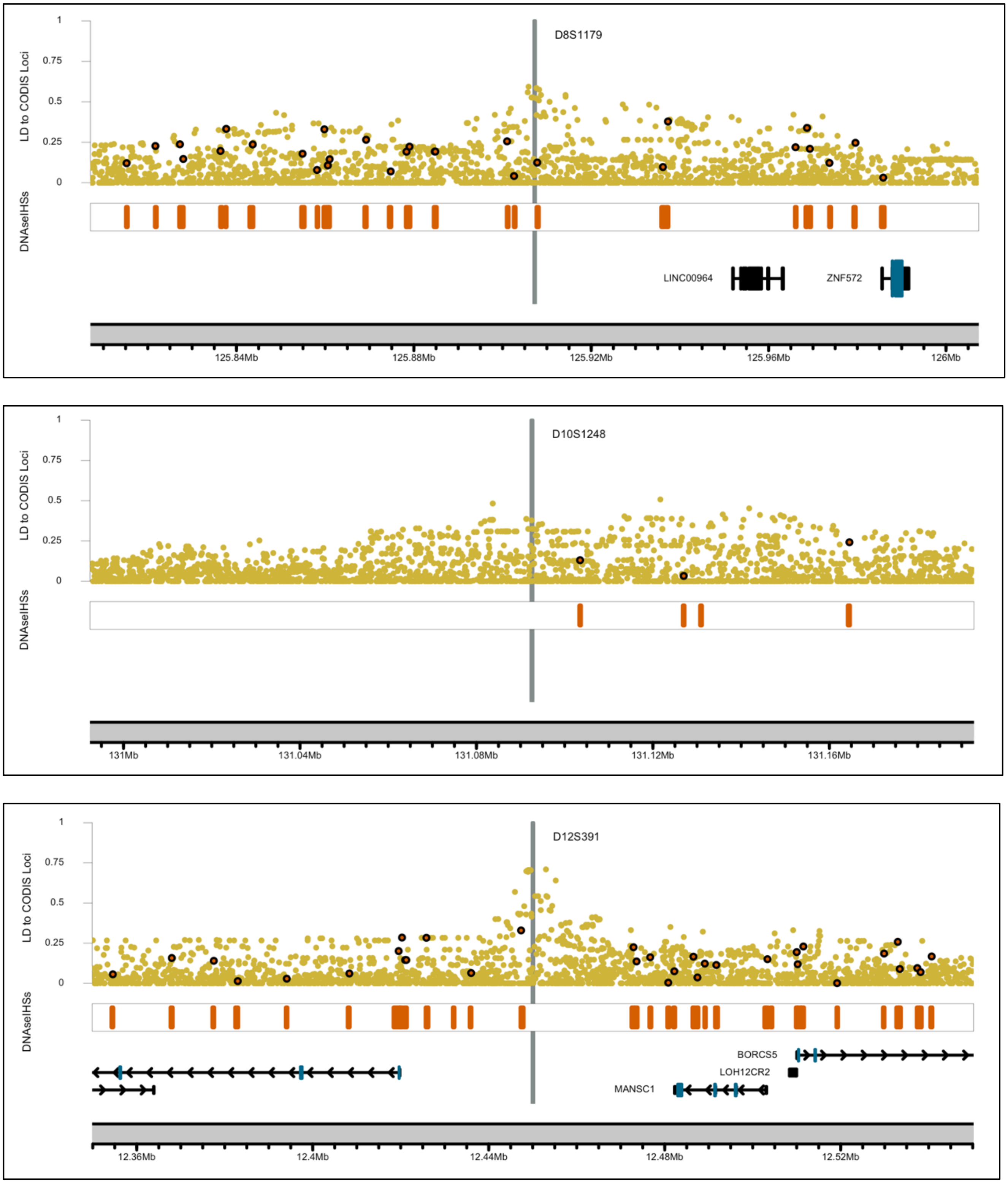

D8S1179
LD to CODIS Loci
DNAseIHSs
LINC00964
ZNF572
125.84Mb
125.88Mb
125.92Mb
125.96Mb
126Mb
D10S1248
131Mb
131.04Mb
131.08Mb
131.12Mb
131.16Mb
D12S391
BORCS5
LOH12CR2
MANSC1
12.36Mb
12.4Mb
12.44Mb
12.48Mb
12.52Mb

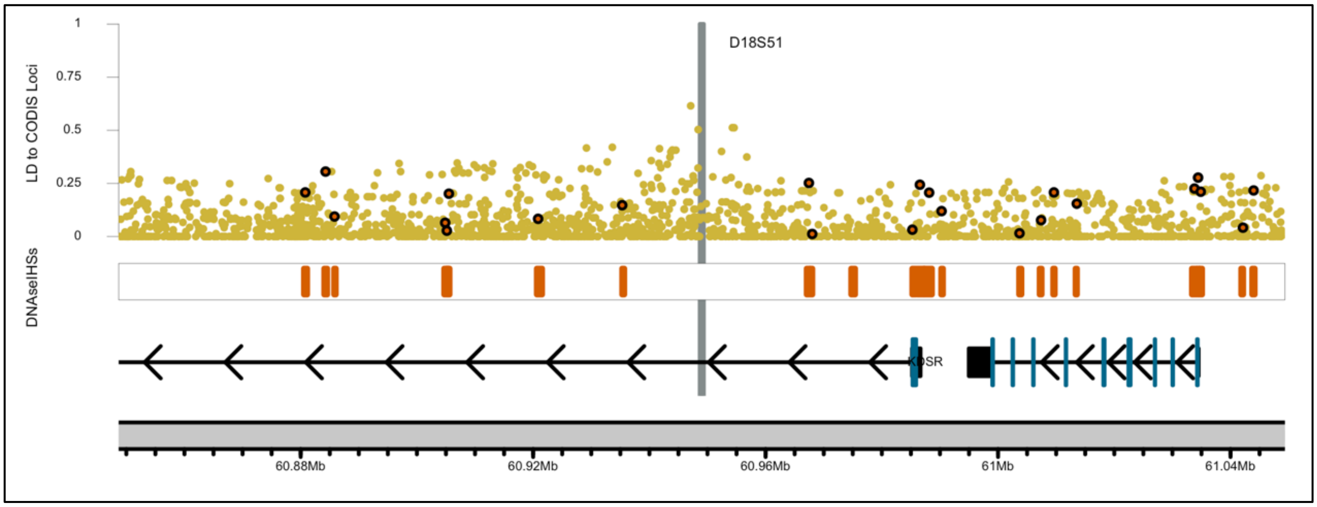
D18S51
LD to CODIS Loci
1
0.75
0.5
0.25
0
DNaseIHSs
DSR
60.88Mb
60.92Mb
60.96Mb
61Mb
61.04Mb

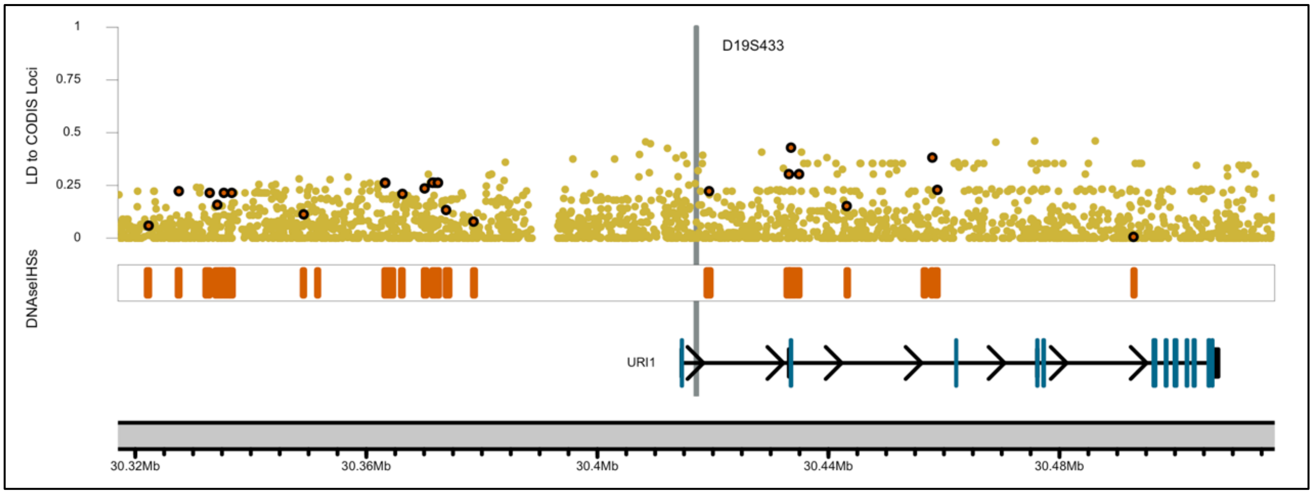
D19S433
LD to CODIS Loci
1
0.75
0.5
0.25
0
DNaseIHSs
URI1
30.32Mb
30.36Mb
30.4Mb
30.44Mb
30.48Mb

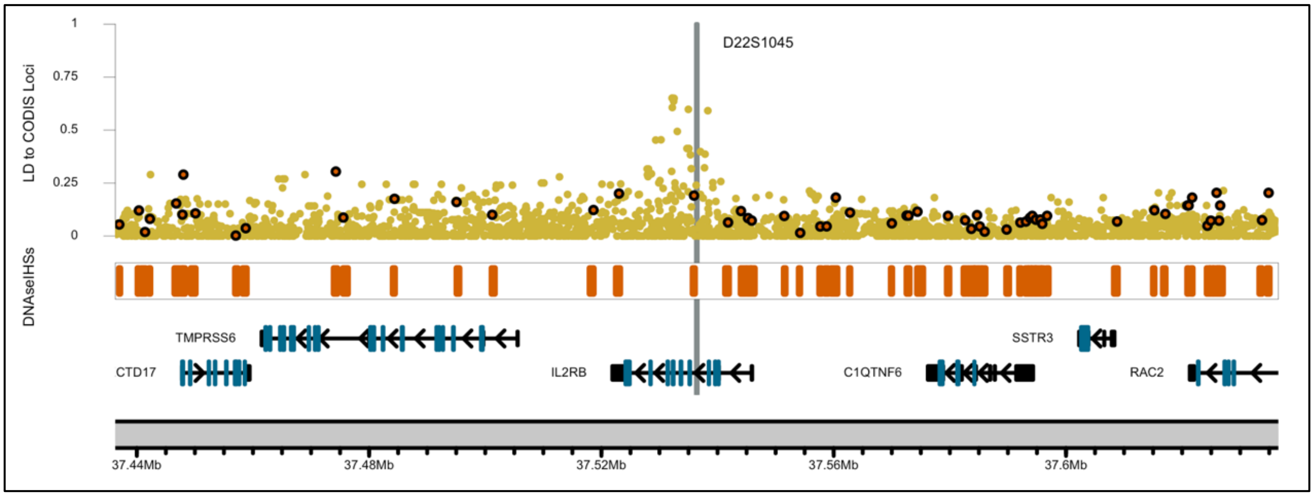
D22S1045
LD to CODIS Loci
1
0.75
0.5
0.25
0
DNaseIHSs
TMPRSS6
SSTR3
CTD17
IL2RB
C1QTNF6
RAC2
37.44Mb
37.48Mb
37.52Mb
37.56Mb
37.6Mb

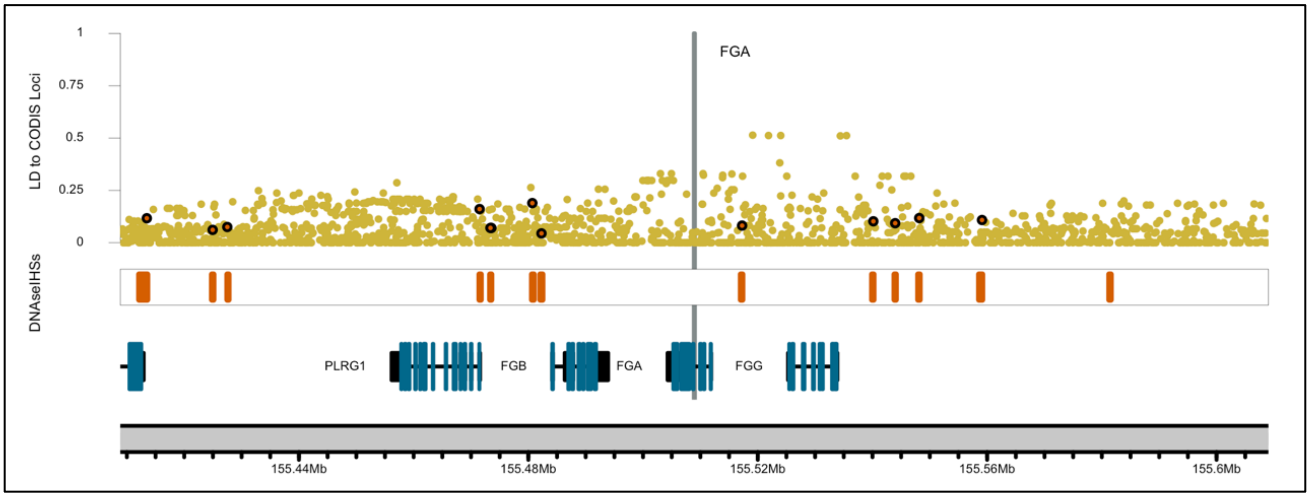
FGA
LD to CODIS Loci
1
0.75
0.5
0.25
0
DNAseIHSs
PLRG1
FGB
FGA
FGG
155.44Mb
155.48Mb
155.52Mb
155.56Mb
155.6Mb

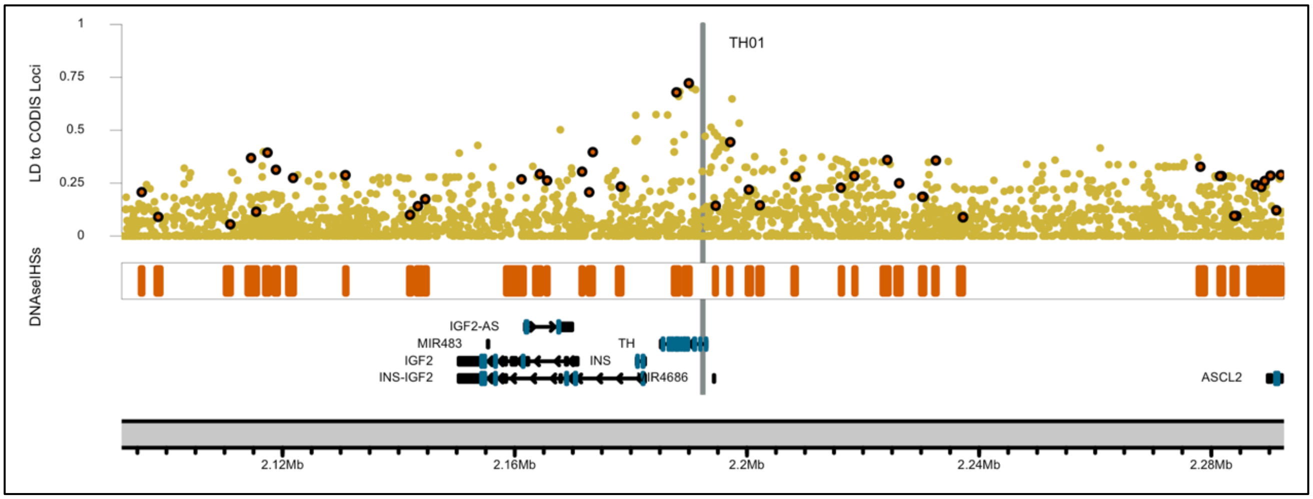
TH01
LD to CODIS Loci
1
0.75
0.5
0.25
0
DNAseIHSs
IGF2-AS
MIR483
IGF2
INS-IGF2
TH
INS
MIR4686
ASCL2
2.12Mb
2.16Mb
2.2Mb
2.24Mb
2.28Mb

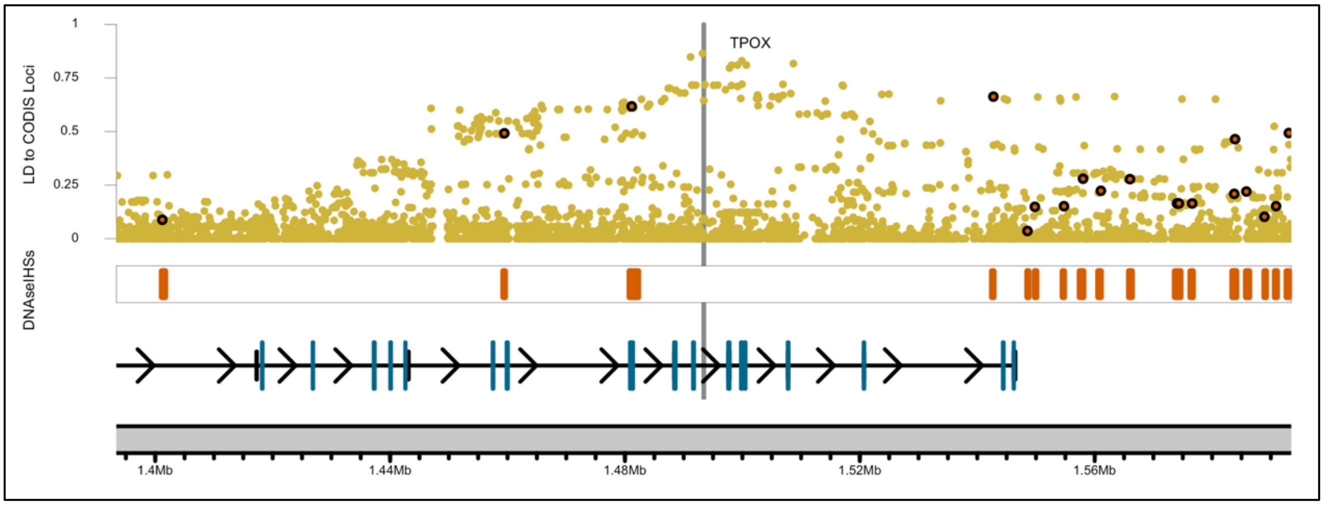
TPOX
LD to CODIS Loci
1
0.75
0.5
0.25
0
DNAseIHSs
1.4Mb
1.44Mb
1.48Mb
1.52Mb
1.56Mb

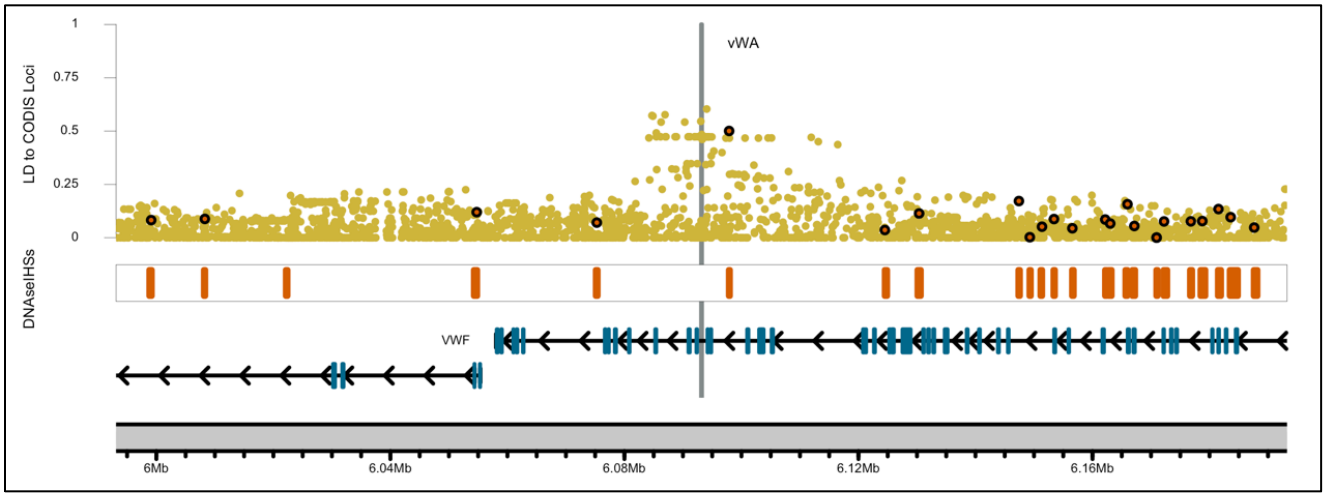
LD to CODIS Loci
1
0.75
0.5
0.25
0
vWA
DNAseIHSs
VWF
6Mb
6.04Mb
6.08Mb
6.12Mb
6.16Mb

**Supplemental Figure 9: LD between CODIS STRs and DNAseIHSs**

The following plots show the distributions of the LD between CODIS loci and DNAseIHSs within 100kb. The five CODISeSTRs (red) are positioned on the left-hand side of the plot. Other CODIS STRs (blue) are positioned on the right-hand side of the plot. Results shown for (A) LD between CODIS loci and DNAseIHSs found within lymphoblast cell lines and (B) LD between CODIS loci and DNAseIHSs found within 20 or more cell line sources.

**A**

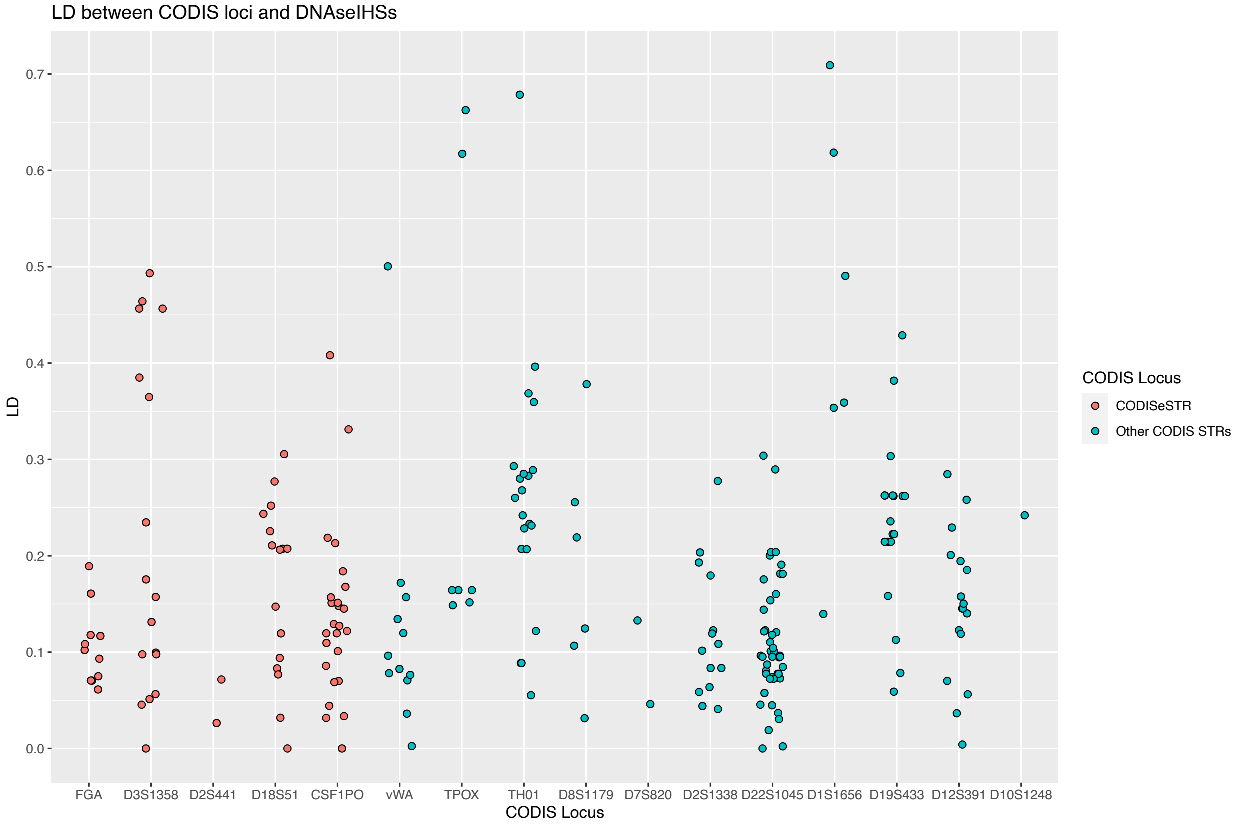

**B**

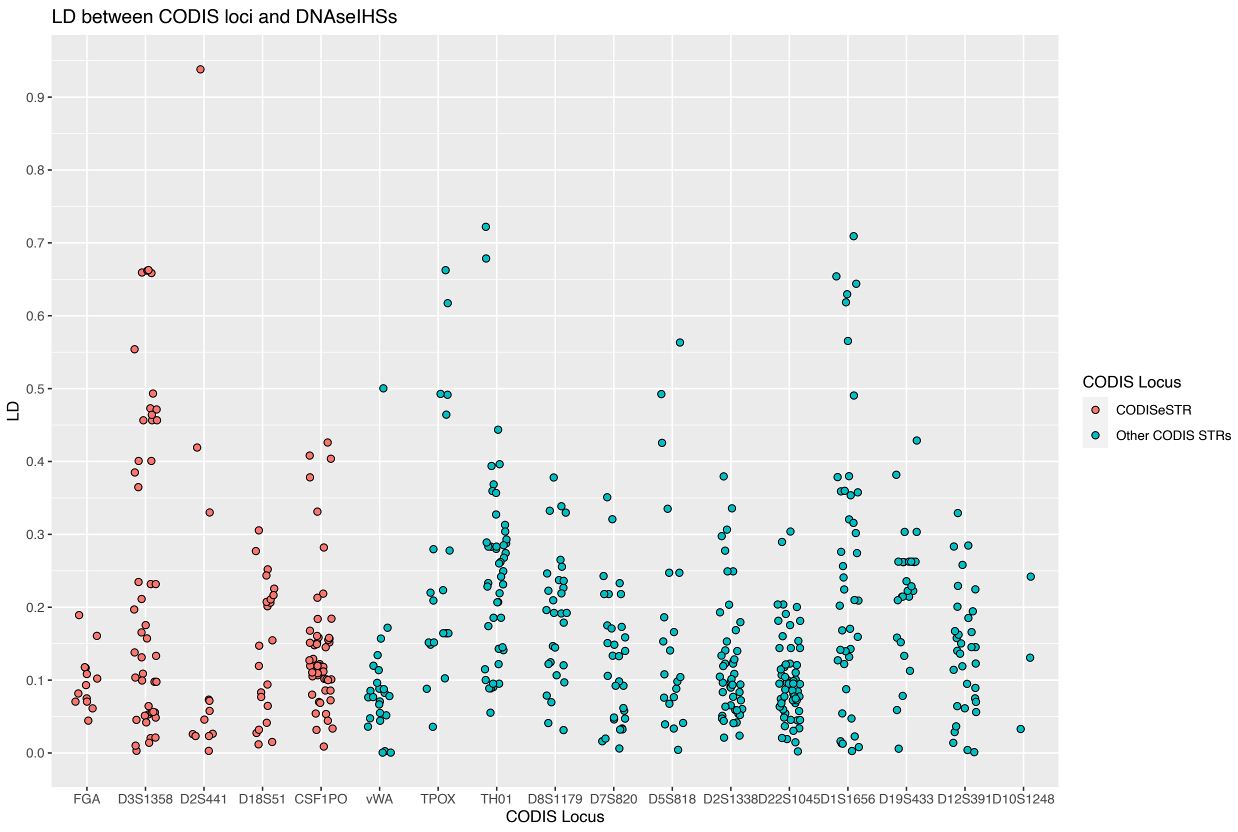

**Supplemental Table 6: Highest LD between CODIS STRs DNAseIHSs**

Table of highest LD between CODIS STRs and SNPs within a DNAseIHS and their corresponding ID and genomic position. Each row represents an individual SNP within a DNAseIHS and its LD with a CODIS STR. DNAseIHSs were within 100kb of the CODIS loci.

**Supplemental Table 7: Table of CODISeSTR-gene pair with associated medical condition**

| CODISeSTR | Gene | Medical Condition |
|---|---|---|
| CSF1PO | CSF1R | Pediatric-onset leukoencephalopathy (Oosterhof et al., 2019) |
| | | Hereditary diffuse leukoencephalopathy with spheroids (HDLS) and other brain malformations (Eichler et al., 2016; Guo et al., 2019; Konno et al., 2014; Nicholson et al., 2013; Rademakers et al., 2012) |
| | | Epilepsy treatment (Srivastava et al., 2018) |
| | | Alzheimer's treatment (Mancuso et al., 2019; Olmos-Alonso et al., 2016; Sosna et al., 2018) |
| | | Spinal cord recovery treatment (Bellver-Landete et al., 2019) |
| | | Major depressive disorder and schizophrenia (Gandal et al., 2018; Shimamoto-Mitsuyama et al., 2021; Zhang et al., 2020) |
| | | Depression and anxiety-like behavior – in mouse models (Chitu et al., 2015; Guo et al., 2019) |
| CSF1PO | TIGD6 | Obesity (Kaewsutthi et al. 2016) |
| D2S441 | C1D | NA |
| D3S1358 | LARS2 | Perrault syndrome (Pierce et al. 2013; Willems et al. 2014; Soldà et al. 2016; Demain et al. 2017) |
| | | MELAS syndrome (R. Li et al., 2010) |
| | | Type II Diabetes (Yao et al., 2003) |
| | | Severe multisystem metabolic disorder ('T Hart et al. 2005; Riley et al. 2016) |
| | | Nasopharyngeal carcinoma (Zhou et al. 2009) |
| | | Bipolar disorder and schizophrenia (Munakata et al. 2005) |
| D18S51 | KDSR | Progressive symmetric erythrokeratoderma (Boyden et al., 2017) |
| | | Low platelet levels and anemia (Bariana et al., 2019; Takeichi et al., 2017) |
| FGA | PLRG1 | NA |

**Supplemental Text 1: Medical impact of LARS2 expression variation**

LARS2 encodes a mitochondrial tRNA synthetase (Soldà et al., 2016). Inherited mutations in LARS2 that reduce or knock out its function have been associated with Perrault syndrome (Pierce et al. 2013; Willems et al. 2014; Soldà et al. 2016; Demain et al. 2017)**,** MELAS syndrome (R. Li et al., 2010; ), as well as having some evidence for association with type II diabetes (Yao et al., 2003), and severe multisystem metabolic disorder ('T Hart et al., 2005; Riley et al., 2016). Somatic mutations that reduce or eliminate LARS2 function have been associated with nasopharyngeal carcinoma (Zhou et al., 2009)**.** Increased expression of LARS2 in brain tissue has been observed in individuals with bipolar disorder and schizophrenia, and associated with a mutation in the mitochondrial leucine tRNA, although it is not clear if the up-regulation of LARS2 is causative or compensatory (Munakata et al., 2005).

**Supplemental Text 2: Medical relevance of C1D expression variation**

C1D encodes a DNA-binding protein involved in DNA repair and preservation following UV exposure, induced by the expression of the XPB gene (G. Li et al., 2010). Although high expression levels of C1D are associated with apoptosis (G. Li et al., 2010; Rothbarth et al., 1999; Tomita et al., 2018), that expression variance appears to be environmentally regulated, so it is unlikely that constitutive C1D expression variation inferred through D2S441 genotypes would reveal medical information.

**Supplemental Text 3: Medical relevance of CSF1R expression variation**

CSF1R encodes a receptor for a cytokine that controls production, differentiation, and function of macrophages (Stanley & Chitu, 2014). Mutations in the coding sequence have been causally associated with a number of brain conditions. Some splice and missense mutations can cause pediatric-onset leukoencephalopathy (Oosterhof et al., 2019), while nonsense mutations have been associated with hereditary diffuse leukoencephalopathy with spheroids (HDLS) and other brain malformations (Eichler et al., 2016; Guo et al., 2019; Konno et al., 2014; Nicholson et al., 2013; Rademakers et al., 2012). Further, the expression of CSF1R regulates microglia, in fact, inhibiting CSf1R protein function has been suggested as therapeutic treatment for neural conditions like epilepsy (Srivastava et al., 2018), Alzheimer's disease (Mancuso et al., 2019; Olmos-Alonso et al., 2016; Sosna et al., 2018), and spinal cord injury recovery (Bellver-Landete et al., 2019) (although pleiotropic impacts may complicate the therapy (Lei et al., n.d.)). Expression and splicing variation of CSF1R have also been associated with major psychiatric disorders including major depressive disorder and schizophrenia in humans (Gandal et al., 2018; Shimamoto-Mitsuyama et al., 2021; Zhang et al., 2020), while heterozygous CSF1R +/- mouse models have displayed depression with anxiety-like behavior (Chitu et al., 2015; Guo et al., 2019).

**Supplemental Text 4: Medical relevance of TIGD6 expression variation**

TIGD6 is a protein coding gene and a member of the pogo family of DNA-mediated transposons (Simpson et al., 2000). While its expression has been detected in many tissues, very little evidence has been found to support its impact to medical traits. However, a case study of 3 individuals suggested that TIGD6 expression variation may differ between obese and non-obese individuals (Kaewsutthi et al., 2016).

**Supplemental Text 5: Medical relevance of KDSR expression variation**

KDSR is a gene that encodes an enzyme utilized in the catalytic reduction of 3-ketohydrosphingosine to dihydrosphingosine (Linn et al., 2001), an important reaction in sphingolipid metabolism to synthesize the lipid ceramide – a biochemical molecule found in many fundamental cellular processes (Pralhada Rao et al., 2013). A number of mutations, including an inversion, deletions, and a silent third base change that caused exon skipping, have been found to reduce or eliminate enzyme function (Boyden et al., 2017). Those mutations are associated with progressive symmetric erythrokeratoderma, which causes patches of thick, scaly, red skin (Boyden et al., 2017). Some individuals with decreased or eliminated KDSR function additionally suffer from low platelet levels and anemia (Bariana et al., 2019; Takeichi et al., 2017).

**Supplemental Text 6: Medical relevance of PLRG1 expression variation**

PLRG1 encodes pleiotropic regulator 1, a spliceosomal protein that plays an important role in the cell division cycle 5-like (CDC5L) complex (Ajuh et al., 2000). The CDC5L complex is a major component of the spliceosome required for pre-mRNA splicing, such that that PLRG1 plays a critical role in alternative splice site selection (Ajuh & Lamond, 2003). Total inactivation of PLRG1 is lethal in mice, while tissuespecific inactivation leads to apoptosis (Kleinridders et al., 2009). While this suggests that PLRG1 is an essential gene, no strong evidence has been found associating PLRG1 variation with medical traits (but see (Lovely et al., 2011)).

### References

'T Hart, L. M., Hansen, T., Rietveld, I., Dekker, J. M., Nijpels, G., Janssen, G. M. C., Arp, P. A., Uitterlinden, A. G., Jørgensen, T., Borch-Johnsen, K., Pols, H. A. P., Pedersen, O., Van Duijn, C. M., Heine, R. J., & Maassen, J. A. (2005). Evidence that the mitochondrial leucyl tRNA synthetase (LARS2) gene represents a novel type 2 diabetes susceptibility gene. *Diabetes*, *54*(6), 1892–1895. https://doi.org/10.2337/diabetes.54.6.1892

Ajuh, P., Kuster, B., Panov, K., Zomerdijk, J. C. B. M., Mann, M., & Lamond, A. I. (2000). Functional analysis of the human CDC5L complex and identification of its components by mass spectrometry. *EMBO Journal*, *19*(23), 6569–6581. https://doi.org/10.1093/emboj/19.23.6569

Ajuh, P., & Lamond, A. I. (2003). Identification of peptide inhibitors of pre-mRNA splicing derived from the essential interaction domains of CDC5L and PLRG1. *Nucleic Acids Research*, *31*(21), 6104–6116. https://doi.org/10.1093/nar/gkg817

Bariana, T. K., Labarque, V., Heremans, J., Thys, C., De Reys, M., Greene, D., Jenkins, B., Grassi, L., Seyres, D., Burden, F., Whitehorn, D., Shamardina, O., Papadia, S., Gomez, K., Van Geet, C., Koulman, A., Ouwehand, W. H., Ghevaert, C., Frontini, M., … Freson, K. (2019). Sphingolipid dysregulation due to lack of functional KDSR impairs proplatelet formation causing thrombocytopenia. *Haematologica*, *104*(5), 1036–1045. https://doi.org/10.3324/haematol.2018.204784

Bellver-Landete, V., Bretheau, F., Mailhot, B., Vallières, N., Lessard, M., Janelle, M. E., Vernoux, N., Tremblay, M. È., Fuehrmann, T., Shoichet, M. S., & Lacroix, S. (2019). Microglia are an essential component of the neuroprotective scar that forms after spinal cord injury. *Nature Communications*, *10*(1), 1–18. https://doi.org/10.1038/s41467-019-08446-0

Boyden, L. M., Vincent, N. G., Zhou, J., Hu, R., Craiglow, B. G., Bayliss, S. J., Rosman, I. S., Lucky, A. W., Diaz, L. A., Goldsmith, L. A., Paller, A. S., Lifton, R. P., Baserga, S. J., & Choate, K. A. (2017). Mutations in KDSR Cause Recessive Progressive Symmetric Erythrokeratoderma. *American Journal of Human Genetics*, *100*(6), 978–984. https://doi.org/10.1016/j.ajhg.2017.05.003

Chitu, V., Gokhan, S., Gulinello, M., Branch, C. A., Patil, M., Basu, R., Stoddart, C., Mehler, M. F., & Richard Stanley, E. (2015). Phenotypic characterization of a Csf1r haploinsufficient mouse model of adult-onset leukodystrophy with axonal spheroids and pigmented glia (ALSP). *Neurobiology of Disease*, *74*, 219–228. https://doi.org/10.1016/j.nbd.2014.12.001

Demain, L. A. M., Urquhart, J. E., O'Sullivan, J., Williams, S. G., Bhaskar, S. S., Jenkinson, E. M., Lourenco, C. M., Heiberg, A., Pearce, S. H., Shalev, S. A., Yue, W. W., Mackinnon, S., Munro, K. J., Newbury-Ecob, R., Becker, K., Kim, M. J., O' Keefe, R. T., & Newman, W. G. (2017). Expanding the genotypic spectrum of Perrault syndrome. *Clinical Genetics*, *91*(2), 302–312. https://doi.org/10.1111/cge.12776

Eichler, F. S., Li, J., Guo, Y., Caruso, P. A., Bjonnes, A. C., Pan, J., Booker, J. K., Lane, J. M., Tare, A., Vlasac, I., Hakonarson, H., Gusella, J. F., Zhang, J., Keating, B. J., & Saxena, R. (2016). *CSF1R* mosaicism in a family with hereditary diffuse leukoencephalopathy with spheroids. *Brain*, *139*(6), 1666–1672. https://doi.org/10.1093/brain/aww066

Gandal, M. J., Zhang, P., Hadjimichael, E., Walker, R. L., Chen, C., Liu, S., Won, H., Van Bakel, H., Varghese, M., Wang, Y., Shieh, A. W., Haney, J., Parhami, S., Belmont, J., Kim, M., Losada, P. M., Khan, Z., Mleczko, J., Xia, Y., … Geschwind, D. H. (2018). Transcriptome-wide isoform-level dysregulation in ASD, schizophrenia, and bipolar disorder. *Science*, *362*(6420). https://doi.org/10.1126/science.aat8127

Guo, L., Bertola, D. R., Takanohashi, A., Saito, A., Segawa, Y., Yokota, T., Ishibashi, S., Nishida, Y., Yamamoto, G. L., Franco, J. F. da S., Honjo, R. S., Kim, C. A., Musso, C. M., Timmons, M., Pizzino, A., Taft, R. J., Lajoie, B., Knight, M. A., Fischbeck, K. H., … Ikegawa, S. (2019). Bi-allelic CSF1R Mutations Cause Skeletal Dysplasia of Dysosteosclerosis-Pyle Disease Spectrum and Degenerative Encephalopathy with Brain Malformation. *American Journal of Human Genetics*, *104*(5), 925–935. https://doi.org/10.1016/j.ajhg.2019.03.004

Kaewsutthi, S., Santiprabhob, J., Phonrat, B., Tungtrongchitr, A., Lertrit, P., & Tungtrongchitr, R. (2016). Exome sequencing in Thai patients with familial obesity. *Genetics and Molecular Research*, *15*(2). https://doi.org/10.4238/gmr.15028311

Kleinridders, A., Pogoda, H.-M., Irlenbusch, S., Smyth, N., Koncz, C., Hammerschmidt, M., & Brüning, J. C. (2009). PLRG1 Is an Essential Regulator of Cell Proliferation and Apoptosis during Vertebrate Development and Tissue Homeostasis. *Molecular and Cellular Biology*, *29*(11), 3173–3185. https://doi.org/10.1128/mcb.01807-08

Konno, T., Tada, M., Tada, M., Koyama, A., Nozaki, H., Harigaya, Y., Nishimiya, J., Matsunaga, A., Yoshikura, N., Ishihara, K., Arakawa, M., Isami, A., Okazaki, K., Yokoo, H., Itoh, K., Yoneda, M., Kawamura, M., Inuzuka, T., Takahashi, H., … Ikeuchi, T. (2014). Haploinsufficiency of CSF-1R and clinicopathologic characterization in patients with HDLS. *Neurology*, *82*(2), 139–148. https://doi.org/10.1212/WNL.0000000000000046

Lei, F., Cui, N., Zhou, C., Chodosh, J., Vavvas, D. G., Βάββας, Δ. Γ., Paschalis, E. I., Πασχάλης Ήλιος, Ε., & Paulson, J. A. (n.d.). *CSF1R inhibition by a small-molecule inhibitor is not microglia specific; affecting hematopoiesis and the function of macrophages*. https://doi.org/10.1073/pnas.1922788117

Li, G., Liu, J., Abu-Asab, M., Masabumi, S., & Maru, Y. (2010). XPB induces C1D expression to counteract UV-induced apoptosis. *Molecular Cancer Research : MCR*, *8*(6), 885–895. https://doi.org/10.1158/1541-7786.MCR-09-0467

Li, R., Chomyn, A., & Guan, M.-X. (2010). Human mitochondrial leucyl-tRNA synthetase corrected mitochondrial dysfunctions due to the MELAS and diabetes associated tRNA Leu(UUR) A3243G mutation Running title: tRNA synthetase corrects mitochondrial dysfunction Downloaded from. *Mol. Cell. Biol*. https://doi.org/10.1128/MCB.01614-09

Linn, S. C., Kim, H. S., Keane, E. M., Andras, L. M., Wang, E., & Merrill, J. (2001). Regulation of de novo sphingolipid biosynthesis and the toxic consequences of its disruption. *Biochemical Society Transactions*, *29*(6), 831–835. https://doi.org/10.1042/bst0290831

Lovely, R. S., Yang, Q., Massaro, J. M., Wang, J., D'Agostino, R. B., O'Donnell, C. J., Shannon, J., & Farrell, D. H. (2011). Assessment of genetic determinants of the association of γ′ fibrinogen in relation to cardiovascular disease. *Arteriosclerosis, Thrombosis, and Vascular Biology*, *31*(10), 2345–2352. https://doi.org/10.1161/ATVBAHA.111.232710

Mancuso, R., Fryatt, G., Cleal, M., Obst, J., Pipi, E., Monzón-Sandoval, J., Ribe, E., Winchester, L., Webber, C., Nevado, A., Jacobs, T., Austin, N., Theunis, C., Grauwen, K., Daniela Ruiz, E., Mudher, A., Vicente-Rodriguez, M., Parker, C. A., Simmons, C., … Perry, V. H. (2019). CSF1R inhibitor JNJ-40346527 attenuates microglial proliferation and neurodegeneration in P301S mice. *Brain*, *142*(10), 3243–3264. https://doi.org/10.1093/brain/awz241

Munakata, K., Iwamoto, K., Bundo, M., & Kato, T. (2005). Mitochondrial DNA 3243A>G mutation and increased expression of LARS2 gene in the brains of patients with bipolar disorder and schizophrenia. *Biological Psychiatry*, *57*(5), 525–532. https://doi.org/10.1016/j.biopsych.2004.11.041

Nicholson, A. M., Baker, M. C., Finch, N. C. A., Rutherford, N. J., Wider, C., Graff-Radford, N. R., Nelson, P. T., Clark, H. B., Wszolek, Z. K., Dickson, D. W., Knopman, D. S., & Rademakers, R. (2013). CSF1R mutations link POLD and HDLS as a single disease entity. *Neurology*, *80*(11), 1033–1040. https://doi.org/10.1212/WNL.0b013e31828726a7

Olmos-Alonso, A., Schetters, S. T. T., Sri, S., Askew, K., Mancuso, R., Vargas-Caballero, M., Holscher, C., Perry, V. H., & Gomez-Nicola, D. (2016). Pharmacological targeting of CSF1R inhibits microglial proliferation and prevents the progression of Alzheimer's-like pathology. *Brain*, *139*(3), 891–907. https://doi.org/10.1093/brain/awv379

Oosterhof, N., Chang, I. J., Karimiani, E. G., Kuil, L. E., Jensen, D. M., Daza, R., Young, E., Astle, L., van der Linde, H. C., Shivaram, G. M., Demmers, J., Latimer, C. S., Keene, C. D., Loter, E., Maroofian, R., van Ham, T. J., Hevner, R. F., & Bennett, J. T. (2019). Homozygous Mutations in CSF1R Cause a Pediatric-Onset Leukoencephalopathy and Can Result in Congenital Absence of Microglia. *American Journal of Human Genetics*, *104*(5), 936–947. https://doi.org/10.1016/j.ajhg.2019.03.010

Pralhada Rao, R., Vaidyanathan, N., Rengasamy, M., Mammen Oommen, A., Somaiya, N., & Jagannath, M. R. (2013). Sphingolipid Metabolic Pathway: An Overview of Major Roles Played in Human Diseases. *Journal of Lipids*, *2013*, 1–12. https://doi.org/10.1155/2013/178910

Rademakers, R., Baker, M., Nicholson, A. M., Rutherford, N. J., Finch, N., Soto-Ortolaza, A., Lash, J., Wider, C., Wojtas, A., Dejesus-Hernandez, M., Adamson, J., Kouri, N., Sundal, C., Shuster, E. A., Aasly, J., MacKenzie, J., Roeber, S., Kretzschmar, H. A., Boeve, B. F., … Wszolek, Z. K. (2012). Mutations in the colony stimulating factor 1 receptor (CSF1R) gene cause hereditary diffuse leukoencephalopathy with spheroids. *Nature Genetics*, *44*(2), 200–205. https://doi.org/10.1038/ng.1027

Riley, L. G., Rudinger-Thirion, J., Schmitz-Abe, K., Thorburn, D. R., Davis, R. L., Teo, J., Arbuckle, S., Cooper, S. T., Campagna, D. R., Frugier, M., Markianos, K., Sue, C. M., Fleming, M. D., & Christodoulou, J. (2016). LARS2 variants associated with hydrops, lactic acidosis, sideroblastic anemia, and multisystem failure. In *JIMD Reports* (Vol. 28, pp. 49–57). Springer. https://doi.org/10.1007/8904_2015_515

Rothbarth, K., Spiess, E., Juodka, B., Yavuzer, U., Nehls, P., Stammer, H., & Werner, D. (1999). Induction of apoptosis by overexpression of the DNA-binding and DNA-PK-activating protein C1D. *Journal of Cell Science*, *112*(13).

Shimamoto-Mitsuyama, C., Nakaya, A., Esaki, K., Balan, S., Iwayama, Y., Ohnishi, T., Maekawa, M., Toyota, T., Dean, B., & Yoshikawa, T. (2021). Lipid Pathology of the Corpus Callosum in Schizophrenia and the Potential Role of Abnormal Gene Regulatory Networks with Reduced Microglial Marker Expression. *Cerebral Cortex*, *31*(1), 448–462. https://doi.org/10.1093/cercor/bhaa236

Simpson, J. C., Wellenreuther, R., Poustka, A., Pepperkok, R., & Wiemann, S. (2000). Systematic subcellular localization of novel proteins identified by large-scale cDNA sequencing. *EMBO Reports*, *1*(3), 287–292. https://doi.org/10.1093/embo-reports/kvd058

Soldà, G., Caccia, S., Robusto, M., Chiereghin, C., Castorina, P., Ambrosetti, U., Duga, S., & Asselta, R. (2016). First independent replication of the involvement of LARS2 in Perrault syndrome by whole-exome sequencing of an Italian family. *Journal of Human Genetics*, *61*(4), 295–300. https://doi.org/10.1038/jhg.2015.149

Sosna, J., Philipp, S., Albay, R. I., Reyes-Ruiz, J. M., Baglietto-Vargas, D., LaFerla, F. M., & Glabe, C. G. (2018). Early long-term administration of the CSF1R inhibitor PLX3397 ablates microglia and reduces accumulation of intraneuronal amyloid, neuritic plaque deposition and pre-fibrillar oligomers in 5XFAD mouse model of Alzheimer's disease. *Molecular Neurodegeneration*, *13*(1), 1–11. https://doi.org/10.1186/s13024-018-0244-x

Srivastava, P. K., Eyll, J. van, Godard, P., Mazzuferi, M., Delahaye-Duriez, A., Steenwinckel, J. Van, Gressens, P., Danis, B., Vandenplas, C., Foerch, P., Leclercq, K., Mairet-Coello, G., Cardenas, A., Vanclef, F., Laaniste, L., Niespodziany, I., Keaney, J., Gasser, J., Gillet, G., … Johnson, M. R. (2018). A systems-level framework for drug discovery identifies Csf1R as an anti-epileptic drug target. *Nature Communications*, *9*(1), 1–15. https://doi.org/10.1038/s41467-018-06008-4

Stanley, E. R., & Chitu, V. (2014). CSF-1 receptor signaling in myeloid cells. In *Cold Spring Harbor Perspectives in Biology* (Vol. 6, Issue 6). Cold Spring Harbor Laboratory Press. https://doi.org/10.1101/cshperspect.a021857

Takeichi, T., Torrelo, A., Lee, J. Y. W., Ohno, Y., Lozano, M. L., Kihara, A., Liu, L., Yasuda, Y., Ishikawa, J., Murase, T., Rodrigo, A. B., Fernández-Crehuet, P., Toi, Y., Mellerio, J., Rivera, J., Vicente, V., Kelsell, D. P., Nishimura, Y., Okuno, Y., … McGrath, J. A. (2017). Biallelic Mutations in KDSR Disrupt Ceramide Synthesis and Result in a Spectrum of Keratinization Disorders Associated with Thrombocytopenia. *Journal of Investigative Dermatology*, *137*(11), 2344–2353. https://doi.org/10.1016/j.jid.2017.06.028

Tomita, T., Ieguchi, K., Takita, M., Tsukahara, F., Yamada, M., Egly, J.-M., & Maru, Y. (2018). C1D is not directly involved in the repair of UV-damaged DNA but protects cells from oxidative stress by regulating gene expressions in human cell lines. *The Journal of Biochemistry*, *164*(6), 415–426. https://doi.org/10.1093/jb/mvy069

Willems, T., Gymrek, M., Highnam, G., 1000 Genomes Project Consortium, T. 1000 G. P., Mittelman, D., & Erlich, Y. (2014). The landscape of human STR variation. *Genome Research*, *24*(11), 1894–1904. https://doi.org/10.1101/gr.177774.114

Yao, Y.-N., Wang, L., Wu, X.-F., & Wang, E.-D. (2003). The processing of human mitochondrial leucyl-tRNA synthetase in the insect cells. *FEBS Letters*, *534*(1–3), 139–142. https://doi.org/10.1016/S0014-5793(02)03833-4

Zhang, J., Chang, L., Pu, Y., & Hashimoto, K. (2020). Abnormal expression of colony stimulating factor 1 receptor (CSF1R) and transcription factor PU.1 (SPI1) in the spleen from patients with major psychiatric disorders: A role of brain–spleen axis. *Journal of Affective Disorders*, *272*, 110–115. https://doi.org/10.1016/j.jad.2020.03.128

Zhou, W., Feng, X., Li, H., Wang, L., Zhu, B., Liu, W., Zhao, M., Yao, K., & Ren, C. (2009). Inactivation of LARS2, located at the commonly deleted region 3p21.3, by both epigenetic and genetic mechanisms in nasopharyngeal carcinoma. *Acta Biochimica et Biophysica Sinica*, *41*(1), 54–62. https://doi.org/10.1093/abbs/gmn006
